# Preoperative Neuropsychology Subtypes Predict Neuropsychological Change after Temporal Lobe resection

**DOI:** 10.64898/2026.08.17.26360571

**Authors:** Lawrence P. Binding, Sherry Liu, Aira Ansara, Anna Miserocchi, Andrew W. McEvoy, Andre Altmann, Alexandra Young, John S Duncan, Sallie Baxendale, Matthias Koepp, Fenglai Xiao

## Abstract

Cognitive outcomes following temporal lobe epilepsy surgery are highly heterogeneous and remain difficult to predict using traditional threshold-based neuropsychological classifications. Here, we implemented Subtype and Stage Inference (SuStaIn) on preoperative neuropsychology to model cognitive function as a continuous network-level process reflecting both pathological burden and compensatory reserve. We identified three distinct latent trajectories: Verbal, Naming, and Visual. The Verbal subtype reflected classic mesial temporal pathology, where postoperative decline aligned with functional adequacy of residual hippocampal tissue. The Naming subtype represented a neocortical-predominant ‘Temporal Plus’ phenotype; despite lower rates of hippocampal sclerosis, these individuals showed severe postoperative verbal memory vulnerability due to un-reorganized frontotemporal language networks. The Visual trajectory demonstrated progressive visuospatial decline with distinct sex-specific reserve profiles and poorer visual recall outcomes. These progression-based trajectories outperformed conventional static classifications in predicting 12-month postoperative outcomes on unseen test data. Operating directly on routine preoperative evaluations without requiring additional testing, this computational framework disentangles pathological burden from network reserve, providing scalable, biologically interpretable biomarkers to guide personalized risk counselling and network-informed surgical planning.

## Introduction

Cognitive impairment is among the most disabling comorbidities of temporal lobe epilepsy (TLE), affecting language, memory, executive function, and quality of life^1–3^. TLE is not simply a focal disorder but a progressive network disease^4^, in which chronic epileptic activity is associated with neurodegeneration^5,6^, accelerated cognitive decline^7–10^, and premature mortality^11^. While anterior temporal lobe resection (ATLR) remains one of the most effective treatments for drug-resistant TLE^12^, postoperative cognitive outcomes remain highly heterogenous^13^. Although many individuals remain cognitively stable or even improve following successful seizure control, others experience progressive cognitive decline, particularly in verbal and visual memory^14^. Identifying people who are most vulnerable to future cognitive deterioration therefore remains a major challenge in epilepsy surgery.

Current approaches to cognitive risk stratification rely largely on clinical characteristics and neuropsychological performance measured at a single time point.^15–19^ Although these variables provide useful prognostic information, they treat cognition as a static phenotype and do not capture the dynamic nature of disease progression. Individuals with similar overall levels of impairment may differ substantially in the cognitive domains affected, the extent of network involvement and the stage of progression along the disease course^20^. Consequently, existing approaches may overlook important biological heterogeneity that contributes to postoperative cognitive trajectories.

Data-driven studies have attempted to address this heterogeneity by identifying cognitive phenotypes within TLE using clustering approaches^21,22^. consistently describe subgroups characterised by isolated memory, combined memory-language deficits, generalised impairment or relatively preserved cognition^14,21,22^. Furthermore, their assumption of symmetrical cluster structure is poorly suited to the non-linear and classical clustering techniques such as *k*-means fail to differentiate between genuine disease subtypes and individual progression stages because it ignores the temporal dependencies inherent in progressive diseases^23^. Furthermore, their assumption of spherical cluster symmetry is ill-suited for elongated, non-linear trajectories characteristic of cognitive progression in neurological disorders^24,25^. As a result, subtype definition remains biological difficult to interpret and have shown limited availability to predict long-term outcome.

This limitation is reflected clinically. Existing cognitive subtypes demonstrated relatively weak association with postoperative seizure or cognitive outcome^14^, while established predictors, including age, side of surgery, and baseline cognitive performance scores remain the principal determinants of prognosis^15–19^. While conceptually they are clinically useful, these variables provide little information about where an individual lies along the underlying trajectory of cognitive decline. A framework capable of separating disease subtype from disease stage may therefore provide a more biologically meaningful description of cognitive vulnerability and improve personalised prognostic assessment.

To address these limitations, we leverage Subtype and Stage Inference (SuStaIn), a machine learning framework designed to disentangle phenotypic heterogeneity from temporal progression by modelling disease as a series of sequential events^23^. Unlike traditional clustering, SuStaIn recognizes that patients with similar clinical presentations may be on divergent biological trajectories or at different points along the same path. By applying this model to a comprehensive preoperative neuropsychological battery, we aim to characterise the latent temporal architecture of cognitive impairment in TLE, identifying distinct cognitive subtypes and the discrete “stages” of decline within them. This approach has successfully characterised heterogeneous neurodegenerative disorders^23,26,27^ but has not been applied to the progressive cognitive changes associated with epilepsy.

In this study, we applied SuStaIn to comprehensive preoperative neuropsychological data from individuals with drug-resistant TLE to reconstruct the latent trajectories of cognitive dysfunction before surgery. We first sought to determine whether cognitive impairment is better explained by distinct progression pathways rather than static cognitive phenotypes. We then evaluated whether an individual’s position within these trajectories predicts long-term postoperative cognitive outcome beyond conventional clinical risk factors. We hypothesise that that jointly modelling cognitive subtype and stage would provide a more biologically meaningful classification of cognitive vulnerability and improve personalised prediction of postoperative cognitive decline.

## Results

The disease progression modelling technique SuStaIn optimises the sequence of abnormality of each cognitive function. This was applied to 549 subjects diagnosed with TLE who underwent temporal lobe resection with abnormality thresholds of z=1,2,3. All subjects underwent a standard neuropsychological evaluation for TLE which probes: verbal and visual learning and memory, naming, working memory, and verbal fluency (Table 1). Reported below are truncated statistical information, full statistical information for all models are reported in Supplementary Materials 1.

**Table 1.** Neuropsychology assessments performed, the neuropsychological concepts they probe and the battery’s they are a part of.

| Assessment | Concepts | Battery |
| --- | --- | --- |
| List learning | Verbal encoding, attention | AMIPB/BMIPB |
| List recall | Verbal retrieval, consolidation |  |
| Design learning | Visual encoding, attention |  |
| Design recall | Visual retrieval, consolidation |  |
| Immediate story recall | Verbal episodic memory, attention |  |
| Delayed story recall | Verbal episodic memory, memory consolidation, executive function |  |
| Picture naming | Lexical retrieval, semantic memory, word-finding | Graded naming test |
| Digit span | Working memory, executive function | WAIS-R and WAIS-III |
| Category fluency (Animals) | Semantic memory, lexical access, executive function |  |
| Letter fluency 'S' | Executive function, lexical access, working memory | Tombaugh et al.,<sup>28</sup> |

Of 549 participants, 461 (84%) subjects exceeded the first abnormality threshold and were assigned to one of three cognitive progression subtypes, whereas the remainder were classified as stage 0 without detectable cognitive abnormality (labelled as subtype 0). SuStaIn identified three distinct trajectories (Figure 1). The largest subgroup was characterised by early impairment in verbal learning and memory (Verbal Memory subtype; n=254, 46%), followed by a Naming subtype (n=119, 22%) in which lexical retrieval abnormalities preceded visual memory deficits, and a Visual Memory subtype (n=88, 16%) characterised by early visuospatial memory impairment. Figure 1 illustrates the progressive evolution of these subtype-specific cognitive profiles across disease stages. Table 2 shows the mean adjusted demographics and clinical information of individuals assigned to each AP subtype.

**Figure 1.**
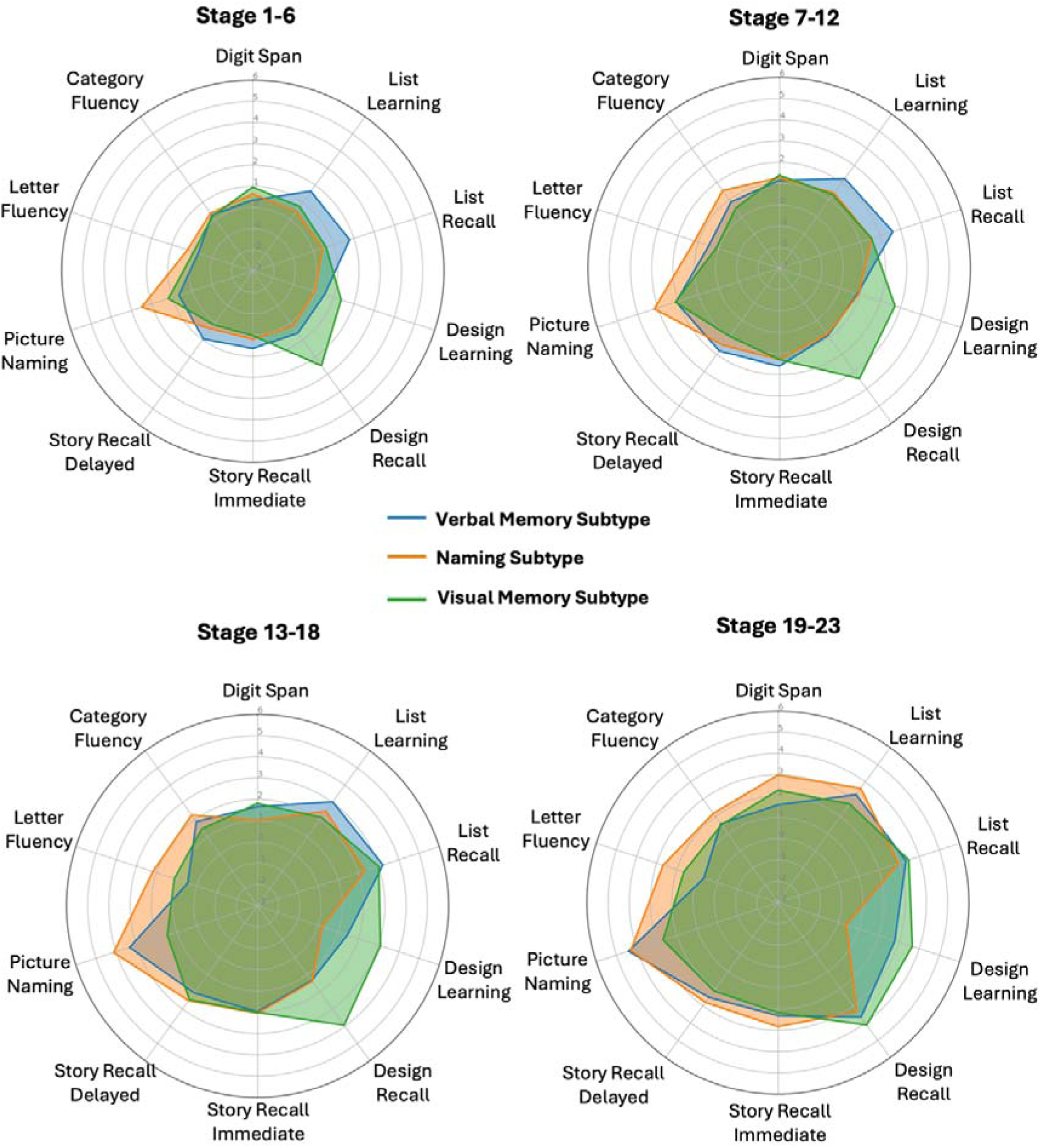
Evolution of neuropsychological profiles across subtype stage and inference (SuStaIn) inferred stages and subtype. Radar plots illustrate the mean cognitive performance of three distinct patient subtypes: Verbal Memory Subtype (blue), Naming Subtype (orange), and Visual Memory Subtype (green), across grouped stages of SuStaIn inferred disease progression. Radial axes denote mean standardised scores across ten neuropsychological assessments with greater peripheral points representing greater deficits. This covers the domains of working memory (Digit Span), verbal memory (List learning, List recall), verbal memory and attention (Story recall (immediate), Story recall (delayed)), visuospatial memory (Design learning, Design recall), and language/fluency (Picture naming, Letter fluency, Category fluency). Solid lines represent the group mean, with shaded regions denoting the encompassing profile area. The cross-sectional trajectories demonstrate that while subtype-specific cognitive patterns are initially overlapping in early disease stages (Stage 1-6), they progressively uncouple. By advanced stages (Stage 19-23), the cohorts exhibit highly distinct, domain-specific multidimensional impairment signatures.

**Table 2.**
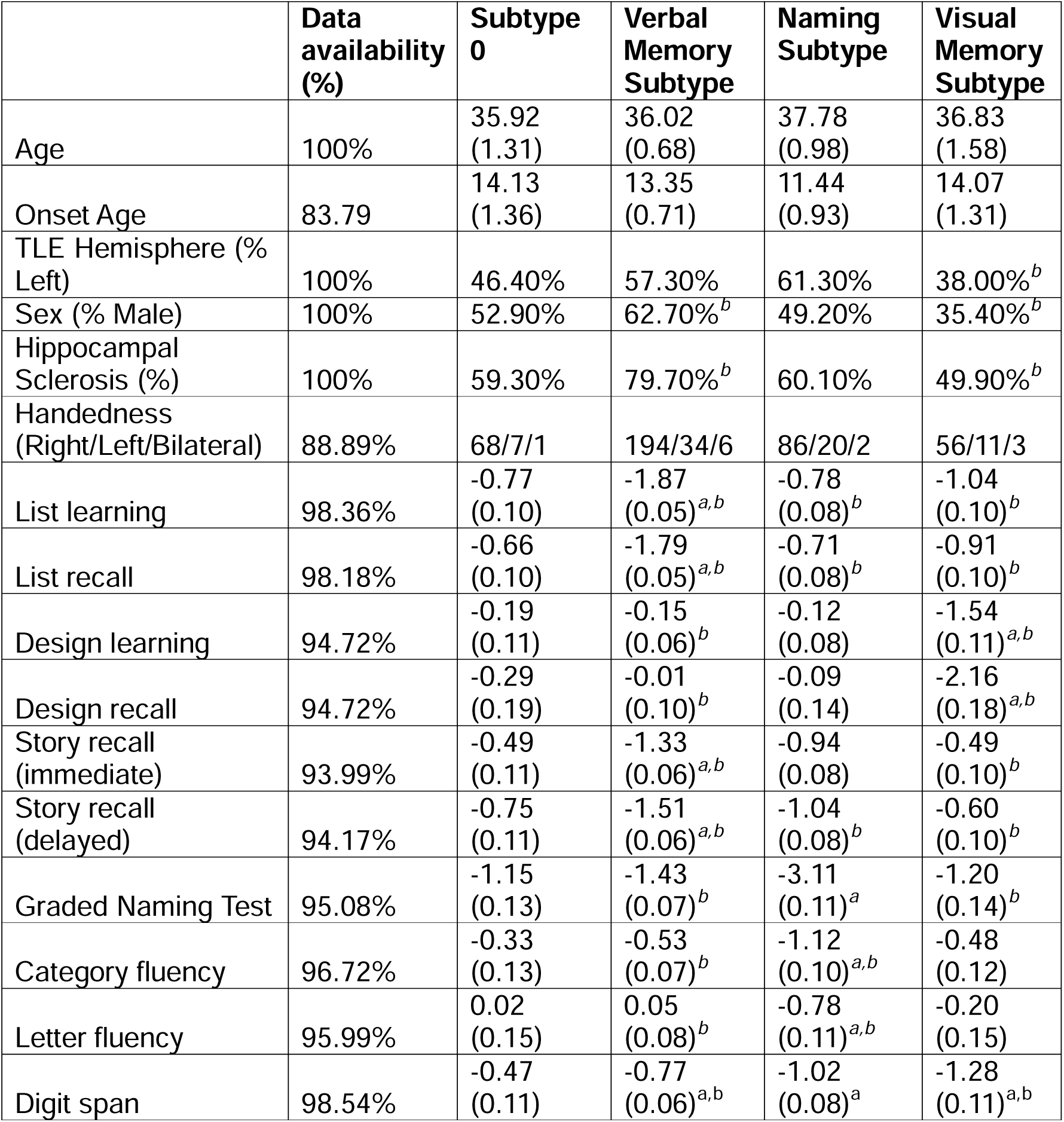
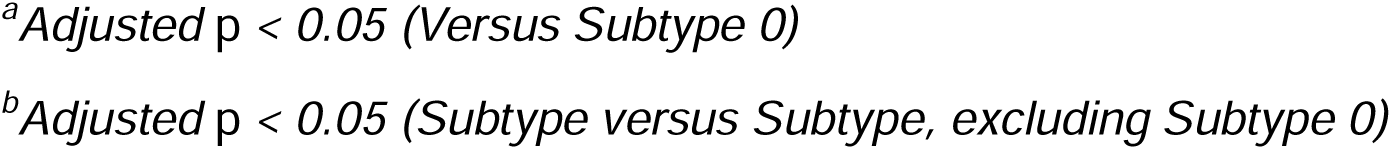
Comparison of demographic, clinical and neuropsychology means between subtypes after correcting for age, sex, TLE Hemisphere (Left, Right), Handedness (Left, Right, Bilateral), age of onset, sustain stage and subtype, unless the variable of interest. TLE hemisphere, Sex, hippocampal sclerosis, are expressed as a proportion (%). Handedness counts are reported. Standard deviations are given in parentheses where relevant. Some neuropsychology scores for subtype 0 are inflated due to adjusting for stage. We have included unadjusted means in Supplementary Materials 2.1.

|  | **Data availability (%)** | **Subtype 0** | **Verbal Memory Subtype** | **Naming Subtype** | **Visual Memory Subtype** |
| --- | --- | --- | --- | --- | --- |
| Age | 100% | 35.92 (1.31) | 36.02 (0.68) | 37.78 (0.98) | 36.83 (1.58) |
| Onset Age | 83.79 | 14.13 (1.36) | 13.35 (0.71) | 11.44 (0.93) | 14.07 (1.31) |
| TLE Hemisphere (% Left) | 100% | 46.40% | 57.30% | 61.30% | 38.00%<sup>b</sup> |
| Sex (% Male) | 100% | 52.90% | 62.70%<sup>b</sup> | 49.20% | 35.40%<sup>b</sup> |
| Hippocampal Sclerosis (%) | 100% | 59.30% | 79.70%<sup>b</sup> | 60.10% | 49.90%<sup>b</sup> |
| Handedness (Right/Left/Bilateral) | 88.89% | 68/7/1 | 194/34/6 | 86/20/2 | 56/11/3 |
| List learning | 98.36% | -0.77 (0.10) | -1.87 (0.05)<sup>a,b</sup> | -0.78 (0.08)<sup>b</sup> | -1.04 (0.10)<sup>b</sup> |
| List recall | 98.18% | -0.66 (0.10) | -1.79 (0.05)<sup>a,b</sup> | -0.71 (0.08)<sup>b</sup> | -0.91 (0.10)<sup>b</sup> |
| Design learning | 94.72% | -0.19 (0.11) | -0.15 (0.06)<sup>b</sup> | -0.12 (0.08) | -1.54 (0.11)<sup>a,b</sup> |
| Design recall | 94.72% | -0.29 (0.19) | -0.01 (0.10)<sup>b</sup> | -0.09 (0.14) | -2.16 (0.18)<sup>a,b</sup> |
| Story recall (immediate) | 93.99% | -0.49 (0.11) | -1.33 (0.06)<sup>a,b</sup> | -0.94 (0.08) | -0.49 (0.10)<sup>b</sup> |
| Story recall (delayed) | 94.17% | -0.75 (0.11) | -1.51 (0.06)<sup>a,b</sup> | -1.04 (0.08)<sup>b</sup> | -0.60 (0.10)<sup>b</sup> |
| Graded Naming Test | 95.08% | -1.15 (0.13) | -1.43 (0.07)<sup>b</sup> | -3.11 (0.11)<sup>a</sup> | -1.20 (0.14)<sup>b</sup> |
| Category fluency | 96.72% | -0.33 (0.13) | -0.53 (0.07)<sup>b</sup> | -1.12 (0.10)<sup>a,b</sup> | -0.48 (0.12) |
| Letter fluency | 95.99% | 0.02 (0.15) | 0.05 (0.08)<sup>b</sup> | -0.78 (0.11)<sup>a,b</sup> | -0.20 (0.15) |
| Digit span | 98.54% | -0.47 (0.11) | -0.77 (0.06)<sup>a,b</sup> | -1.02 (0.08)<sup>a</sup> | -1.28 (0.11)<sup>a,b</sup> |

<sup>a</sup>Adjusted $p < 0.05$ (Versus Subtype 0)
<sup>b</sup>Adjusted $p < 0.05$ (Subtype versus Subtype, excluding Subtype 0)

### Stability of Subtypes

To test whether the SuStaIn subtypes represent a distinct set of patterns, we compared the probability that each individual belonged to a particular subtype, reflecting the strength of assignment of an individual to a subtype. For individuals assigned to a subtype, most fell into stereotypical progression of each subtype. However, 9.54% of individuals did not fall cleanly into any subtype. The majority of these cases were in stage 1 (29.55%) or stage 2 (9.09%) and had a diagnosis of right TLE (56.82%).

### Subtypes characterised by distinct clinical, demographic and preoperative cognitive profiles

To test for specific clinical phenotypic profiles (Table 2), we compared subtype clinical, demographic, and cognitive variables to subtype 0 (no significant cognitive abnormality; stage = 0) and to all other subtypes grouped together (excluding subtype 0). Covariates such as sex, onset age, age, TLE side, handedness, stage, subtype were regressed out when not the variable of interest. All test were corrected for multiple comparisons using false discovery rate (FDR)

### Comparison to Subtype 0

The verbal memory subtype had significantly greater proportion of cases with hippocampal sclerosis (Odds ratio=2.704, adjusted *p*=0.012). Furthermore, they presented with significantly worse list learning (Estimate = −1.098, adjusted *p*<0.001), and list recall (Estimate = −1.138, adjusted *p*<0.001), both story recall immediate (Estimate = −0.839, adjusted *p*<0.001) and story recall delayed (Estimate = −0.760, adjusted *p*<0.001) as well as digit span (Estimate= −0.304, adjusted *p* = 0.044) cognitive functions.

The naming subtype presented with significantly worse story recall (immediate) (Estimate = −0.449, adjusted *p*=0.002), picture naming (Estimate = −1.965, adjusted *p*<0.001), category (Estimate = −0.789, adjusted *p*<0.001) and letter fluency (Estimate = −0.797, adjusted *p*<0.001) as well as digit span (Estimate = −0.550, adjusted *p* < 0.001).

The visual memory subtype patients presented with significantly worse Design learning (Estimate = −1.354, adjusted *p*<0.001), recall (Estimate = −1.872, adjusted *p*<0.001) and digit span (Estimate = −0.811, adjusted *p* < 0.001). There was no significant difference in age, TLE onset, TLE side, gender and handedness between subtype assignment.

### Between Subtype Comparison

The verbal memory subtype presented with significantly greater odds of being male (Odds Ratio = 2.170, adjusted *p* < 0.001) and having hippocampal sclerosis (Odds ratio = 3.042, adjusted *p* < 0.001). They had significantly worse list learning (Estimate = −1.001, adjusted *p* < 0.001), list recall (Estimate = −1.004, adjusted *p* < 0.001), Story recall immediate (Estimate = −0.572, adjusted *p* < 0.001), and delayed (Estimate = −0.640, adjusted *p* < 0.001) but significantly better Design learning (Estimate = 0.489, adjusted *p* < 0.001) and recall (Estimate = 0.855, adjusted *p* < 0.001), Picture naming (Estimate = 0.935, adjusted *p* < 0.001), Category (Estimate = 0.341, adjusted *p* = 0.001) and letter (Estimate = 0.612, adjusted *p* < 0.001) fluency as well as digit span (Estimate = 0.364, adjusted *p* < 0.001).

The naming subtype were significantly less likely to have hippocampal sclerosis (Odds ratio = 0.532, adjusted *p* = 0.030) and worse picture naming (Estimate = −1.728, adjusted *p* < 0.001), category fluency (Estimate = −0.613, adjusted *p* < 0.001) and letter fluency (Estimate = −0.778, adjusted *p* < 0.001). However, they did have better scores in list learning (Estimate = 0.920, adjusted *p* < 0.001) and list recall (Estimate = 0.894, adjusted *p* < 0.001), design learning (Estimate = 0.332, adjusted *p* = 0.009), and story recall delayed (Estimate = 0.263, adjusted *p* = 0.050).

The visual memory subtype contained significantly less left TLE cases (Odds Ratio = 0.440, adjusted *p* = 0.006), males (Odds Ratio = 0.392, adjusted *p* = 0.002) and cases with hippocampal sclerosis (Odds ratio = 0.372, adjusted *p* = 0.002). They had with poorer performance in design learning (Estimate = −1.409, adjusted *p* < 0.001) and design recall (Estimate = −2.145, adjusted *p* < 0.001) as well as digit span (Estimate = −0.446, adjusted *p* < 0.001). They did have better performance in list learning (Estimate = 0.483, adjusted *p* < 0.001), list recall (Estimate = 0.523, adjusted *p* < 0.001), story recall immediate (Estimate = 0.726, adjusted *p* < 0.001), story recall delayed (Estimate = 0.771, adjusted *p* < 0.001), and picture naming (Estimate = 0.719, adjusted *p* < 0.001) performance.

### Association of Stage and Performance

Increased SuStaIn assigned stage severity (i.e., more assessments exceeding the abnormality thresholds) was significantly related to a higher chance of left TLE (Odds Ratio= 1.046, adjusted *p* = 0.037), worse preoperative List learning (Estimate = −0.144, adjusted *p* < 0.001), List recall (Estimate = −0.132, adjusted *p* < 0.001), Design learning (Estimate = −0.096, adjusted *p* < 0.001), Design recall (Estimate = −0.097, adjusted *p* < 0.001), Story recall (immediate) (Estimate = −0.130, adjusted *p* < 0.001) and delayed (Estimate = −0.138, adjusted *p* < 0.001), Picture naming (Estimate = −0.168, adjusted *p* < 0.001), Category fluency (Estimate = −0.120, adjusted *p* < 0.001), letter fluency (Estimate = −0.103, adjusted *p* < 0.001) and digit span (Estimate = −0.080, adjusted *p* < 0.001) performance.

### Association with Postoperative Cognitive Outcome

We investigated whether the preoperative cognitive subtypes had any effect on the change between preoperative and 12-month postoperative temporal lobe resection neuropsychological outcome. We compared subtype clinical, demographic, and cognitive variables to subtype 0 and to all other subtypes grouped together (excluding subtype 0). Covariates such as sex, onset age, age, TLE side, handedness, stage, subtype and preoperative cognitive scores were regressed out. Model adjusted means and standard deviations for postoperative neuropsychology can be seen in Table 3.

**Table 3.** Comparison of postoperative neuropsychology means between subtypes after correcting for age, sex, TLE Hemisphere (Left, Right), Handedness (Left, Right, Bilateral), age of onset, sustain stage and subtype, and preoperative neuropsychology score. Some neuropsychology scores for subtype 0 are inflated due to adjusting for stage. We have included unadjusted means in Supplementary Materials 2.2.

|  | **Data availability (%)** | **Subtype 0** | **Verbal Memory Subtype** | **Naming Subtype** | **Visual Memory Subtype** |
| --- | --- | --- | --- | --- | --- |
| List learning 12 months change | 88.52% | 0.24<br>(0.15) | -0.28<br>(0.08)<sup>a</sup> | -0.20<br>(0.11)<sup>a</sup> | -0.11<br>(0.13) |
| List recall 12 months change | 88.52% | 0.26<br>(0.15) | -0.22<br>(0.09)<sup>b</sup> | -0.03<br>(0.12) | -0.03<br>(0.14) |
| Design learning 12 months change | 84.34% | -0.04<br>(0.14) | -0.09<br>(0.07) | -0.17<br>(0.10) | -0.34<br>(0.15) |
| Design recall 12 months change | 84.15% | -0.07<br>(0.17) | 0.07<br>(0.09) | -0.09<br>(0.13) | -0.86<br>(0.18)<sup>a,c</sup> |
| Story recall (immediate) 12 months change | 83.79% | 0.30<br>(0.15) | -0.11<br>(0.07)<sup>b</sup> | -0.20<br>(0.10)<sup>a</sup> | 0.07<br>(0.13) |
| Story recall (delayed) 12 months change | 83.79% | 0.40<br>(0.14) | -0.00<br>(0.07)<sup>b</sup> | -0.13<br>(0.10)<sup>a</sup> | 0.11<br>(0.14) |
| Picture Naming 12 months change | 83.79% | -0.03<br>(0.12) | 0.00<br>(0.07) | -0.16<br>(0.14) | 0.13<br>(0.11) |
| Category fluency 12 months change | 83.42% | 0.45<br>(0.14) | 0.15<br>(0.07) | 0.05<br>(0.10)<sup>b</sup> | 0.28<br>(0.12) |
| Letter fluency 12 months change | 82.33% | 0.12<br>(0.15) | 0.30<br>(0.08) | 0.23<br>(0.11) | 0.18<br>(0.13) |
| Digit span 12 Months change | 82.33% | 0.27<br>(0.24) | 0.30<br>(0.12) | 0.11<br>(0.17) | 0.04<br>(0.22) |

<sup>a</sup>Adjusted $p < 0.05$ (Versus S0)
<sup>b</sup>Unadjusted $p < 0.05$ (Versus S0)
<sup>c</sup>Adjusted $p < 0.05$ (Subtype versus Subtype, excluding S0)

### Comparison against Subtype 0

The verbal memory subtype had significantly greater decline in list learning (Estimate = −0.519, adjusted *p* = 0.037) and approaching significance after FDR adjustment for list recall (Estimate = −0.480, adjusted *p* = 0.053), story recall immediate (Estimate = −0.410, adjusted *p* = 0.053) and story recall delayed (Estimate = −0.401, adjusted *p* = 0.053).

The naming subtype had significantly greater decline in list learning (Estimate = −0.441, adjusted *p* = 0.037), story recall immediate (Estimate = −0.500, adjusted *p* = 0.022) and story recall delayed (Estimate = −0.531, adjusted *p* = 0.018) and approaching significance after correction for category fluency (Estimate = −0.399, adjusted *p* = 0.052).

The visual memory subtype had significantly greater decline in design recall (Estimate = −0.788, adjusted *p* = 0.025).

### Comparison Between Subtypes

The visual memory subtype had significantly greater decline of Design recall (Estimate=−0.904, *p* < 0.001). The verbal memory subtype approached significance for having a better outcome of Design recall (Estimate = 0.408, adjusted *p* = 0.060).

### Association of Stage and Postoperative Change

Increased SuStaIn assigned stage severity was related to greater postoperative decline in List learning (Estimate = −0.070, adjusted *p* < 0.001), List recall (Estimate = −0.037, adjusted *p* = 0.010), Design learning (Estimate = −0.067, adjusted *p* < 0.001), Design recall (Estimate = −0.108, adjusted *p* < 0.001), Story recall (immediate) (Estimate = −0.038, adjusted *p* = 0.003), Picture naming (Estimate = −0.314, adjusted *p* = 0.005), Category fluency (Estimate = −0.569, adjusted *p* < 0.001), and letter fluency outcome (Estimate = −0.491, adjusted *p* < 0.001).

### Predictive utility

In-sample nested linear regression models (ANOVA) confirmed that incorporating SuStaIn stage and subtype significantly improved variance explained across all measured postoperative scores relative to baseline clinical covariates alone (all 0.01; Supplementary Materials 2). Crucially, to evaluate true out-of-sample predictive utility on unseen data, we performed 5-fold cross-validation where test-set subjects were phenotyped out-of-sample using fold-specific SuStaIn models. As shown in Figure 2, full models consistently (except in the case of Story recall (delayed)) maintained higher out-of-sample explained variance (*R*^2^_OOS_) across unseen test folds, confirming that the incremental value of SuStaIn phenotypes and stages translates to genuine prognostic utility.

**Figure 2.**
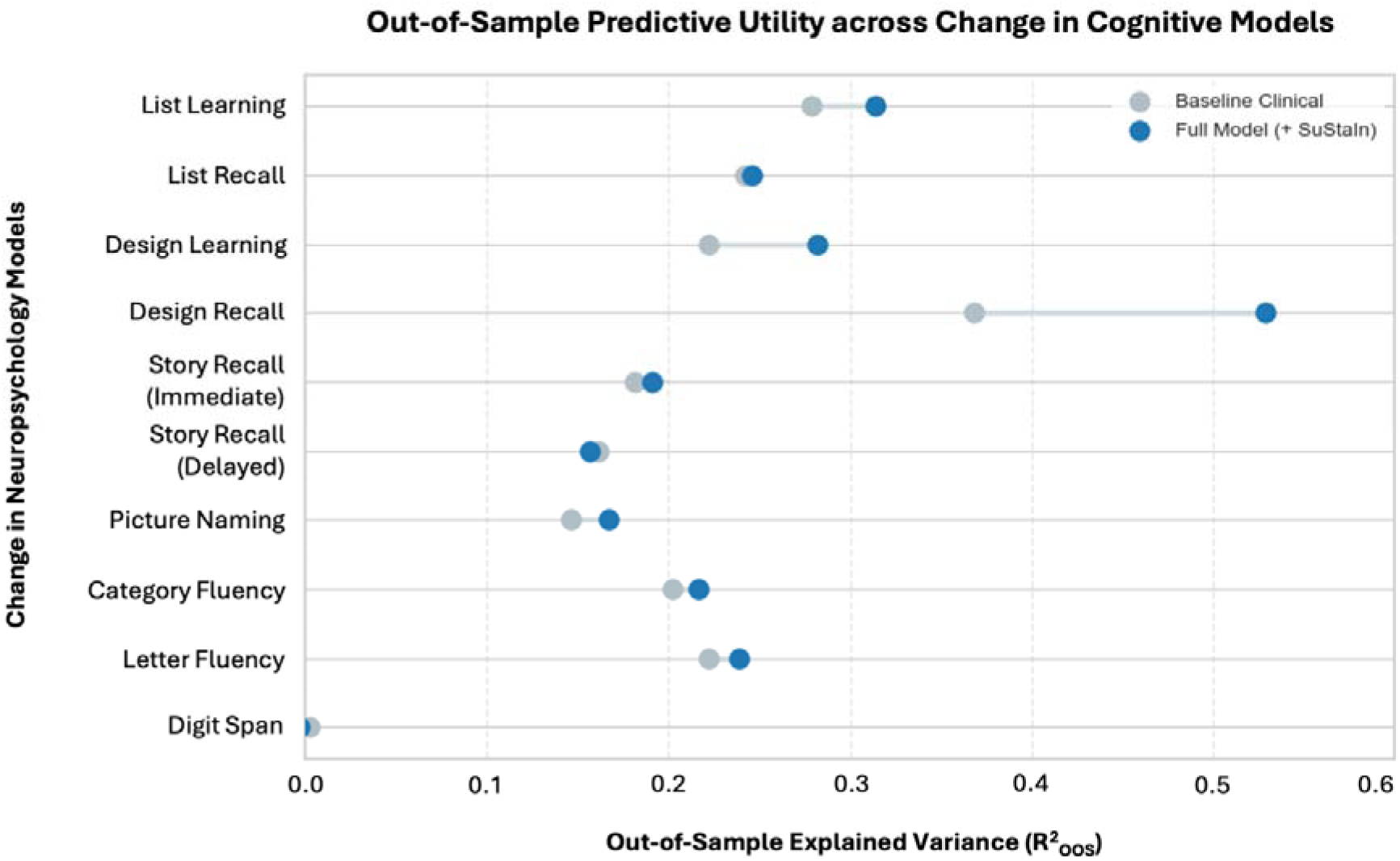
Out-of-sample predictive utility of SuStaIn phenotypes in 12-month postoperative cognitive outcomes. Dumbbell plot illustrating cross-validated out-of-sample explained variance (R^2^_OOS_) evaluated via strict 5-fold cross-validation on unseen test data across nine cognitive parameters. Light grey circles represent baseline clinical models incorporating standard covariates (sex, age at onset, age at surgery, TLE side, handedness, and preoperative cognitive score). Blue circles represent full models additionally incorporating out-of-sample SuStaIn disease stage and subtype assignments. The connecting line highlights the incremental variance gained(ΔR^2^_OOS_)by incorporating disease progression phenotypes. To prevent data leakage, test-set subjects were phenotyped out-of-sample using fold-specific SuStaIn models trained strictly on four-fifths of the cohort.

### Floor Sensitivity Analysis

To account for capacity to decline, we conducted sensitivity analyses iteratively excluding preoperative scores near the test floors. SuStaIn stage and subtype associations remained robust to floor effects across most cognitive domains. The only exceptions occurred at higher exclusion thresholds: stage associations lost significance on the List recall (when excluding scores ≤ 6), while the verbal and visual memory subtypes lost significance on the List recall and Design recall, respectively (when excluding scores ≤ 4) (Supplementary Material 3).

### Postoperative Seizure Outcome

Up to five years of seizure free information was available. We converted international league against epilepsy (ILAE) outcome to binary (ILAE class 1 vs. rest). To determine whether the presence of any specific subtype was associated with seizure relapse employing a Cox Proportional Hazards model, adjusting for duration, TLE side, sex handedness, age of onset, subtype and stage. We employed the same methodology, assessing outcome compared to subtype 0, and subtype vs. subtype excluding subtype 0. There were no statistically significant differences between subtype and ILAE outcome.

### Neuroimaging Correlates

A structural T1-weighted MRI scan was available for 231 subjects. We utilised cortical atrophy to probe structural differences. W-scored meta regions-of-interest (ROI) were created to minimise the number of comparisons regressing out age, gender and scanner differences, these included bilateral hippocampus and amygdala, medial-temporal, extra-temporal, sub-cortical, frontal, parietal, occipital, and cingulate (see Supplementary Materials 4 for full list of regions included). We compared subtype to each subtype excluding subtype 0 (resulting in N=188 subjects) to investigate unique subtype specific profiles. A linear regression was used to compare regions regressing out subtype stage severity. These effects did not survive correction for multiple comparisons and should therefore be interpreted cautiously

The T-value is visualised for regions that were significant (unadjusted) in Figure 3. Subjects in the verbal memory subtype had significantly greater atrophy in bilateral subcortical regions. The naming subtype had significantly greater atrophy in the right extra-temporal and left frontal lobe. Finally, the visual memory subtype had significantly greater cortical thickness in the right extra-temporal, occipital hippocampus/amygdala, and sub-cortical regions.

**Figure 3.**
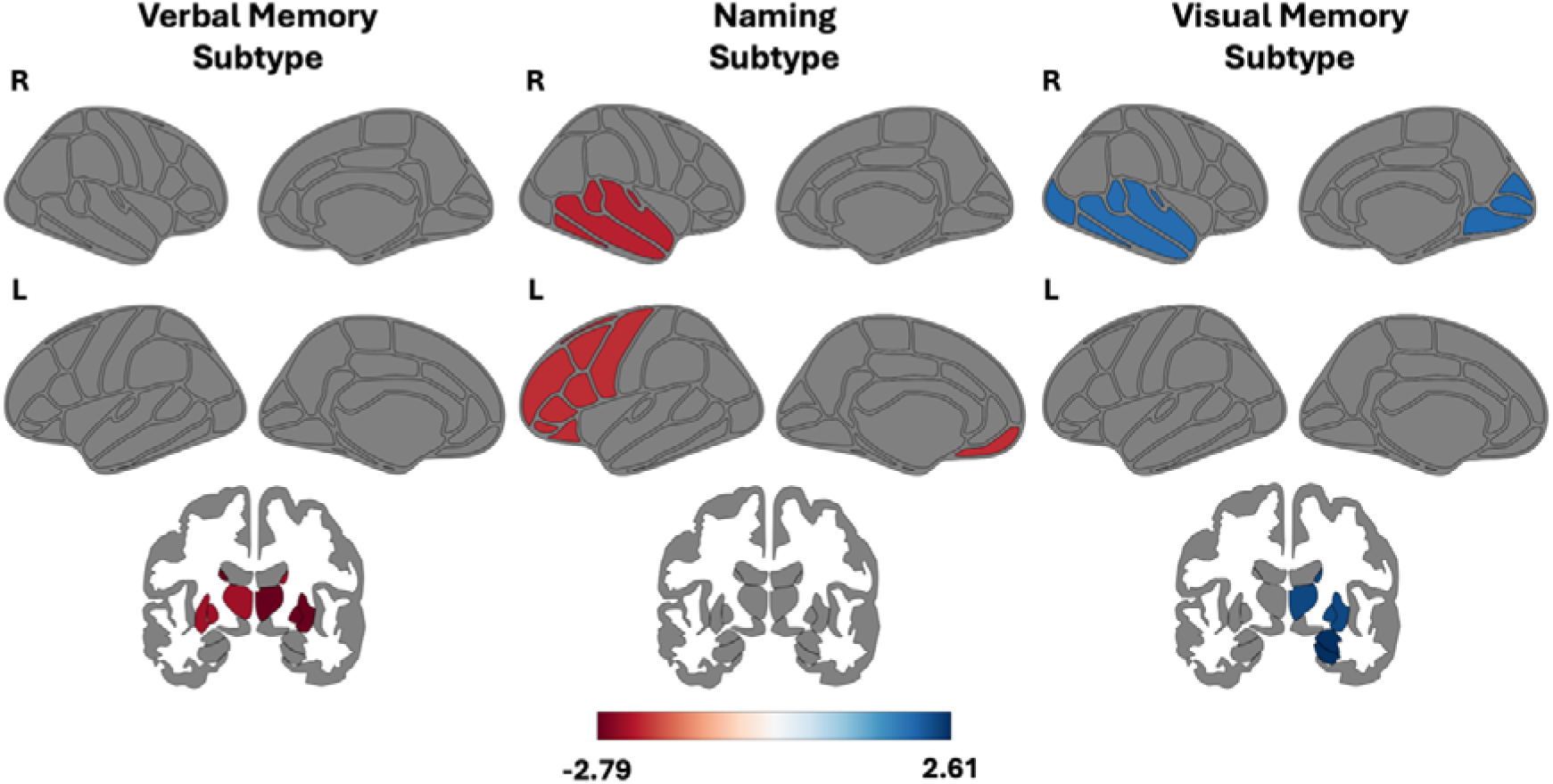
Distinct regional patterns of cortical thickness abnormalities across clinical subtypes. Statistical maps illustrating focal structural deviations for the Verbal Memory, Naming, and Visual Memory patient subtypes, statistically adjusted for overall disease stage. Surface renderings project subtype-specific differences onto lateral and medial views of the right (R) and left (L) hemispheres, alongside representative coronal slices to capture subcortical involvement. The bi-directional colour bar denotes T-value from a linear regression; warm colours (red) indicate regions of significant relative cortical thinning (atrophy), whereas cool colours (blue) represent areas of relative structural preservation or increased thickness compared to other subtypes.

## Discussion

Temporal lobe epilepsy is increasingly recognised as a progressive disorder of distributed brain networks rather than an isolated hippocampal disease^4–10^. Yet neuropsychological classification has largely remained based on static cognitive profiles that provide only limited insight into disease progression or postoperative vulnerability.^21,22^. By applying SuStaIn to comprehensive preoperative neuropsychological data, we demonstrate that cognitive impairment in TLE is better understood as a series of continuous trajectories that differ both in phenotype and stage of progression (Figure 1). Importantly, these latent trajectories predicted postoperative cognitive outcome beyond typical demographic, clinical and baseline neuropsychological variables, suggesting that they capture biologically meaningful differences in network organisation rather than simply classifying existing impairment.

Rather than identifying isolated deficits, SuStaIn revealed three distinct subtypes, which appear to represent different modes of network failure: “Verbal”, “Visual” and “Naming” subtype”. These subtypes align with contemporary models describing epilepsy as a disorder of large-scale network organisation in which chronic seizures progressively disrupt distributed cognitive systems ^5,6^ Importantly, these trajectories are progressive rather than categorical. Individuals classified within one subtype do not simply exhibit a fixed pattern of impairment but occupy different stages along evolving cognitive pathways, indicating that apparently isolated deficits may represent early manifestations of broader network dysfunction. This concept extends previous cognitive taxonomies^21,22^ by introducing temporal progression as a fundamental characteristic of cognitive heterogeneity in TLE.

The largest trajectory, the Verbal memory subtype, closely reflected the classical mesial temporal memory phenotype.^29^ Its association with hippocampal sclerosis and prominent impairment of list learning and story recall is consistent with extensive evidence that hippocampal and hippocampal-diencephalic networks underpin episodic verbal memory^30,31^ ^32,33^ However, this subtype also demonstrated the greatest postoperative decline in list learning, recall and story recall (immediate and delayed), suggesting that preserved preoperative memory performance reflects residual functional capacity within compromised mesial temporal structures rather than successful compensation elsewhere. These findings therefore remain compatible with the functional adequacy model, whereby postoperative decline reflects removal of tissue that continues to support memory despite chronic pathology^34,35^.

The naming subtype represents the most biologically informative finding of the present study. Unlike the classic mesial temporal phenotype, this trajectory was characterised by a progressive impairment of naming, verbal fluency, working memory, and immediate story recall suggesting predominant dysfunction of distributed neocortical language networks rather than isolated hippocampal pathology. Naming depends upon coordinated interactions between lateral temporal cortex, inferior frontal regions and intact white matter connectivity^36,37^ linking these systems, including the ventral and dorsal language streams^38^ The accompanying impairment of verbal fluency and working memory therefore suggests disruption extending beyond the temporal lobe into broader frontotemporal networks, consistent with a “Temporal Plus” profile^39^.

This subtype also demonstrated the greatest vulnerability to post-operative verbal memory decline despite having a lower probability of hippocampal sclerosis. One possible explanation is that hippocampal sclerosis itself promotes adaptive functional reorganisation over many years. Previous functional imaging studies have shown posterior displacement and bilateral redistribution of language in patients with longstanding mesial temporal pathology, thereby preserving language despite progressive hippocampal damage^40^. In contrast, patients without hippocampal sclerosis may retain critical language representation within the anterior and lateral temporal neocortex, rendering these regions particularly vulnerable during standard surgical resections. Rather than reflecting hippocampal dysfunction alone, postoperative decline therefore appears to arise from disruption of distributed language networks that have not undergone compensatory reorganisation.

The Visual subtype similarly illustrates how network organisation influences postoperative outcome. This trajectory was characterised by right-sided pathology, hippocampal sclerosis and selective visuospatial memory impairment, consistent with classical model of specific memory lateralisation^3,41^. Unlike language, visuospatial memory exhibits more limited opportunities for bilateral functional redistribution, remaining highly dependent upon right mesial temporal structures. Consequently, despite the presence of hippocampal sclerosis, resection produced significant postoperative decline in delayed visual recall, indicating that even structurally abnormal hippocampi continue to contribute meaningfully to visuospatial processing. Together with the Verbal subtype, these findings suggest that postoperative cognitive outcome reflects the interaction between residual hippocampal function and the capacity of distributed networks to reorganise.

The observed sex distribution across trajectories further clarifies how biological sex interacts with cognitive network reserve. In contrast to studies reporting minimal influence of sex on cognitive phenotypes in epilepsy^14^, men were proportionally over-represented in the Verbal Subtype, whereas women were relatively over-represented in the Visual Trajectory. This finding aligns with established neuropsychological literature demonstrating material-specific sex differences: men typically exhibit lower baseline verbal memory reserve^42^ and stronger left-hemispheric language lateralization^43^, rendering them more susceptible to localized verbal memory decline following left temporal pathology^3^. Conversely, men possess a well-documented baseline advantage in visuospatial memory, providing a functional buffer against visual domain deficits^44,45^. The over-representation of women in the visual trajectory may reflect a lower baseline visuospatial reserve coupled with a greater reliance on distributed temporal-parietal networks and verbal mediation strategies for visual processing ^46^. Such prolonged compensation may delay overt cognitive impairment but simultaneously increase dependence on distributed neocortical systems, potentially explaining the greater postoperative vulnerability observed within the Naming subtype. Rather than representing a simple demographic association, these findings support the broader concept that reserve and network organisation modify the clinical expression of epilepsy.

Taken together, the three trajectories suggest that reserve, rather than pathology alone, determines clinical vulnerability to a worsened deficit after temporal lobe surgery. Cognitive performance appears to remain relatively preserved while distributed networks retain sufficient redundancy but declines once compensatory capacity becomes exhausted. This interpretation closely aligns with emerging network theories of cognition^35,47,48^ and with recent evidence that chronic epilepsy accelerates biological brain ageing through progressive network dysfunction.^5,6^ Rather than representing discrete neuropsychological syndromes, the identified trajectories may therefore reflect different reserve pathways through which distributed cognitive systems adapt and, ultimately, decompensate in response to longstanding epilepsy

These observations also explain why progression-based modelling outperformed conventional neuropsychological classification. Traditional cognitive taxonomies assign individuals to fixed categories according to arbitrary thresholds^14^ ignoring both disease stage and interactions between cognitive domains. SuStaIn instead models’ cognition as a continuous process in which deficits emerge sequentially across interconnected networks. By separating subtype from stage, the algorithm captures meaningful biological heterogeneity that static clustering approaches cannot distinguish^18,18,32^. The resulting trajectories therefore provide substantially richer information regarding future cognitive vulnerability than conventional classifications based solely on impaired versus preserved performance.

The clinical implications are considerable. Because SuStaIn operates using routine neuropsychological assessment, it could readily be incorporated into preoperative evaluation without additional investigations. More importantly, trajectory-based classification may inform increasingly personalised surgical strategies. Patients demonstrating distributed neocortical language trajectories may benefit from more extensive language mapping or surgical approaches designed to preserve critical white matter pathways supporting language function^36,37,49^. Conversely, patients exhibiting more focal mesial temporal trajectories may be suitable for conventional resections. More broadly, cognitive trajectory modelling may complement emerging network-based approaches to epilepsy surgery, in which structural and functional connectivity increasingly guide resection planning^39,50^.

Several limitations of the present study warrant consideration. First, the data were derived from a single tertiary centre, lacking an internal healthy control cohort for direct comparison. Neuropsychological scores were referenced to published normative data rather than sex- and education-adjusted controls. Educational attainment, an important contributor to cognitive reserve, was unavailable and could therefore not be incorporated into the present analyses. External validation across independent multicentre cohorts will be required to determine the generalisability of the identified trajectories. Finally, neuropsychological performance, as well as the change in neuropsychology scores remains susceptible to transient influences including fatigue, seizure proximity and patient engagement. Although probabilistic modelling reduces the impact of measurement variability preoperatively, prospective longitudinal studies will be important to determine the stability of subtype assignment over time as well as the long-term outcome of these subtypes.

In conclusion, we demonstrate that cognitive heterogeneity in temporal lobe epilepsy reflects distinct trajectories of network organisation and reserve rather than fixed neuropsychological phenotypes. Progression-based modelling identifies biologically meaningful pathways that explain postoperative cognitive outcome beyond conventional threshold-based clinical predictors by capturing the granular variance and temporal progression of network-wide erosion. The-predominant ‘Naming Subtypes’ further suggests that distributed language networks provide prolonged functional compensation, particularly in individuals with greater bilateral language organisation, but become vulnerable when these un-reorganised neocortical systems are disrupted surgically. These findings establish cognitive trajectories a network-based biomarkers of reserve, providing a framework for personalised risk prediction and network-informed tailored surgical interventions. More broadly, they integrate naturally with emerging models of epilepsy as a progressive network disorder in which cognitive decline reflects the interaction between pathological burden, functional reserve and adaptive brain reorganisation.

## Methods

### Participants

Neuropsychology data from 549 subjects who underwent temporal lobe resection for TLE at the National Hospital for Neurology and Neurosurgery, London, United Kingdom between 1991 and 2019 were included. Eligible participants had completed preoperative neuropsychological assessment, with valid data available from at least two cognitive domains, and had at least one neuropsychological test score available at least one year after surgery. This project was approved by the Health Research Authority and London – Bloomsbury Research Ethics Committee (REC reference: 20/LO/0149). All patients had the opportunity to opt out of research. This project did not carry any risk to participants and was conducted retrospectively on clinically acquired data.

A recent public release of healthy controls (N=100) using the same scanner as patients were identified^51^. These were included for creating w-scores for the “neuroimaging correlates” analysis.

### Neuropsychology

All patients completed a standardised presurgical neuropsychological assessment evaluating intellectual functioning, memory, language, and executive abilities. This study utilised scores from the Wechsler Adult Intelligence Scales (WAIS-R and WAIS-III)^52^, the Adult Memory and Information Processing Battery (AMIPB)^53^ or its updated version, the Birt Memory and Information Processing Battery (BMIPB)^54^, the Graded Naming Test^55^, and verbal fluency tasks^28^. Twelve scores derived from these assessments were selected to capture performance across four core cognitive domains (see Table 1).

The BMIPB is an updated version of the AMIPB, retaining the same test structures but incorporating revised normative data and alternative stimuli. Several subtests from the AMIPB/BMIPB were used to assess verbal and visual episodic memory.

In the Story Recall task, participants listen to a brief narrative comprising 60 scorable informational units and are asked to recall as much detail as possible immediately after hearing the story and again following a 40-minute delay. The List Learning task—structurally analogous to the California Verbal Learning Test but standardised on a British population—involves the auditory presentation of 15 common words. Participants recall the list across five trials (maximum score = 75). In the delayed recall condition, following exposure to a distractor list, participants are asked to recall the original word list.

The Design Learning task evaluates visuospatial learning and memory. Participants are shown a geometric design consisting of nine connected lines arranged on a 4×4 dot matrix. After a 10-second presentation, they are required to reproduce the design on a blank grid. This procedure is repeated across five trials, and the total number of correctly drawn lines is recorded (maximum score = 45). A delayed recall trial is administered following a distractor design, in which participants must reproduce the original pattern.

The Graded Naming Test assesses lexical retrieval and naming ability. Participants are presented with 30 black-and-white line drawings of objects ordered by increasing difficulty. They are asked to name each item, with performance scored by the total number of correct responses.

Verbal fluency was evaluated using both phonemic and semantic fluency tasks. In the phonemic fluency condition (commonly referred to as the FAS task), participants are asked to generate as many unique words as possible within 60 seconds that begin with a specific letter (typically “F,” “A,” or “S”), excluding proper nouns and morphological variants (e.g., “farm” and “farming”). This task primarily engages executive function, lexical retrieval, and cognitive flexibility. The total number of valid responses across the three trials serves as the outcome measure. As this was a pre-surgical neuropsychological screening, only the letter “S” was performed.

In the semantic fluency condition, participants are asked to name as many animals as possible within 60 seconds. This task depends more heavily on intact semantic memory and temporal lobe function and is particularly sensitive to early cognitive changes seen in neurodegenerative conditions. Performance is indexed by the total number of correct animal names produced.

The Digit Span subtest of the WAIS was used to assess auditory attention and working memory. It includes two components: Digit Span Forward and Digit Span Backward. In the Forward task, participants are read sequences of digits and asked to repeat them in the same order, reflecting short-term memory span. In the Backward task, they must repeat the digits in reverse order, engaging working memory and mental manipulation. Sequence length increases incrementally, and testing continues until the participant fails both trials at a given span length. A combined total score across both components provides an overall index of attentional and working memory capacity.

Neuropsychology data was also collected at 12 months. For identifying changes in cognition from preoperative to postoperative scores, we subtracted 12-month outcome from the preoperative scores.

### MRI Data Acquisition

Between 2004-2013 (patients=130, controls=70) patients were scanned on a 3T GE Signa Excite HDx. A 3D T1-weighted FSPGR sequence at 0.94×0.94×1.1 mm was acquired.^56^ Between 2014-2019 (patients=101, controls=30) patients were scanned on a 3T GE Discovery MR750. A 3D T1-weighted MPRAGE sequence at 1×1×1 mm resolution was acquired.^57^

### Data Preprocessing

Neuropsychology scores from the standardised tests were converted to z-scores based on the published means and standard deviations for the healthy control sample in each test. For the Graded Naming Test we utilised updated normative database^58^. These z-scores were then flipped for SuStaIn so positive z-scores represented worse scores compared to healthy controls.

T1-weighted MRI images were pre-processed using FreeSurfer 8.^59^ All segmentations were manually inspected to ensure accurate cortical segmentation, errors were manually corrected for and then re-inspected. Cortical thickness measures were then extracted according to the Desikan-Killiany atlas^60^ with subcortical volumes being obtained for bilateral hippocampus, amygdala, caudate, nucleus accumbens, putamen, pallidum and thalamus. Meta-ROI were created to reduce the number of comparisons, these included bilateral hippocampus and amygdala, medial-temporal, extra-temporal, sub-cortical, frontal, parietal, occipital, and cingulate (see Supplementary Materials 4 for full list of regions included). Each meta-ROI was expressed as a w-score relative to a control population. A linear mixed effect model was used to create w-scores treating age and gender as fixed and site differences as a random effect on a cohort of healthy controls.

### Subtype Stage and Inference

Efforts to cluster cognitive subtypes in TLE using data-driven approaches are often confounded by variation in disease stage. It is well established that cortical abnormalities in TLE progress with increasing duration since seizure onset. Prior clustering studies have identified either focal or global cognitive impairments, but these patterns may simply reflect different points along a shared trajectory of disease progression. Without accounting for disease stage, clustering algorithms are likely to separate individuals based primarily on their progression level, which limits the ability to detect distinct subtypes of cognitive impairment that are theoretically orthogonal to disease severity.

The SuStaIn algorithm addresses this limitation by integrating clustering with disease progression modelling. Detailed methodological descriptions of SuStaIn are available elsewhere^23^. SuStaIn models linear transitions across discrete z-score thresholds that represent increasing severity levels, separately for each biomarker. The input to SuStaIn is a subject-by-feature matrix. In this application the features correspond to z-scored preoperative neuropsychological test scores. The number of discrete severity thresholds defines the temporal resolution of the progression model, and the effective feature space is the product of the number of neuropsychological measures and the number of thresholds.

While input features are z-scored, some neuropsychological tests are disproportionately affected by TLE. To ensure interpretability, thresholds that exceeded the maximum observed z-score for a given measure were excluded from modeming, so that the modelled trajectory ranged from normal (z=0) to the maximum observed abnormality. For the primary analysis, we used thresholds of z=1, 2, and 3 for all metrics, selected arbitrarily to reflect increasing severity. This resulted in the max stage of 23 due to some neuropsychology tests 95^th^ percentile not exceeding the maximum z-score. Additional analyses were performed with alternative thresholds to confirm that the findings were not dependent on the specific z-score cutoffs (see Supplementary Materials 5).

We utilised cross-validation to determine the optimal number of neuropsychological subtypes. Separately for each *1-5* subtypes fivefold cross-validation was performed, in each fold, SuStaIn was trained on 80% of the data and tested on the remaining 20%, with this process repeated across folds. Cross-validation-based information criterion (CVIC) as well as the out-of-sample log-likelihood was used to evaluate the optimal number of subtypes. Figures from this cross-validation process can be seen in Supplementary Materials 6. The CVIC score flattened out at a three-subtype model indicating the best fit to the data.

We then applied SuStaIn to the full dataset using the selected three subtypes. The model assumes a uniform prior over subtypes and stages (i.e., all combinations are equally probable a priori) and is initialised using an expectation–maximisation procedure, which eliminates the need for a burn-in period^23^. Uncertainty in model estimates was quantified using 10,000 iterations of Markov chain Monte Carlo sampling. For each individual, SuStaIn estimated the probability of belonging to every subtype-stage combination, with individuals assigned to their maximum likelihood subtype and stage. Participants showing no evidence of abnormality across any features were labelled as ‘stage 0’ and not assigned to a subtype.

### Statistical Analysis

Reported in the results are a truncated version of the full statistical information, full details can be seen in Supplementary Materials 1.

Several demographic, cognitive and diagnostic variables were available for nearly all individuals. These variables included TLE hemisphere, age, sex, hippocampal sclerosis presence, age of onset, handedness and ILAE outcome (for up to five years). Neuropsychology information was available as defined in Table 1.

We statistically compared these demographic, clinical and neuroimaging markers between subtypes. For continuous variables (age, age of onset, and all neuropsychology z-scores) we utilised an ordinary least squares linear regression. For categorical variables (TLE hemisphere, sex, and hippocampal sclerosis) we utilised a logistic regression. For these statistical comparisons, we regressed out covariates including, TLE hemisphere, sex, handedness, age and age of onset sustain subtype and stage unless they were the dependent variable. For handedness, due to the multinomial variable (left, right, bilateral) we utilised a contingency table with chi squared test.

Subtype characterisation was performed using two methods: (1) Comparison with sustain negative group: Subjects that were characterised as subtype 0, that is not having any neuropsychology value reaching the abnormality threshold (i.e., stage 0) were compared against subjects that were assigned into specific subtypes. This was achieved via a regression treating subtype 0 as the reference group. (2) Comparison to all other sustain subtypes: A one-versus-all approach was applied to subtyped individuals only to assess how different subtypes differed from one another. Separately for each subtype, models were fitted with a single dummy variable entered indicating membership to that subtype.

All models were subjected to diagnostics to ensure our data fulfilled the relevant assumptions of statistical tests. For ordinary least squares regression, we utilised Durbin-Watson for autocorrelation, Cook’s distance for outliers, and Breusch-Pagen to assess homoscedasticity. If models violated homoscedasticity, the model was corrected, and the corrected results are reported. *p*-values were adjusted using FDR method with an alpha threshold of 0.05.

To evaluate the out-of-sample predictive utility of SuStaIn phenotypes on 12-month postoperative cognitive score changes while strictly preventing data leakage, we implemented a 5-fold cross-validation framework. Within each cross-validation fold, SuStaIn subtype and stage assignments for both training and unseen test subjects were inferred using fold-specific SuStaIn model parameters trained exclusively on four-fifths of the cohort. We then fitted two nested linear regression models on the training fold: (1) a baseline clinical model including standard covariates (TLE side, sex, handedness, age at onset, age at surgery, and baseline preoperative score); and (2) a full model that additionally incorporated out-of-sample SuStaIn disease stage (treated continuously) and subtype assignment (one-hot encoded). Categorical features were dummy encoded. Missing values across features and target variables were handled via complete-case analysis per fold. Out-of-sample predictions across all five unseen test folds were pooled to evaluate global model performance using out-of-sample explained variance (R^2^_OOS_)

For these cases, ILAE seizure outcome was available at one-year periods from 1 year up to 5 years. We converted ILAE outcome to a binary variable (ILAE1 == 1, ILAE>1 ==0). To assess whether these neuropsychology subtypes impacted ILAE seizure recurrence we utilised a cox-hazard proportional model, the same covariates as above were regressed out.

The neuroimaging analysis involved removing subjects assigned to subtype 0 to assess each subtype against other subtypes. We utilised a linear regression using covariates of measured subtype and neuropsychological stage against each meta-ROI. We visualise the T-statistic from the regions that were statistically significant.

## Data Availability

All data produced in the present study are available upon reasonable request to the authors. Code and inference scripts will be made available online at: https://github.com/lbinding/TLE_Neuropsych_SuStaIn

## Acknowledgements

The authors gratefully acknowledge the Epilepsy Society for their essential infrastructural and clinical support, which facilitated this research. We extend our sincere thanks to the dedicated clinical neuropsychologists who administered the cognitive assessments over the years, and to the radiographers responsible for the MRI data acquisition. Most importantly, we dedicate this work to the individuals with temporal lobe epilepsy who generously contributed their time and data. This research would not have been possible without their participation.

## Disclosures / Competing Interests

No disclosures.

## Funding

LPB and AY are funded by the Wellcome Trust [227341/Z/23/Z]. For the purpose of open access, the author has applied a CC-BY public copyright licence to any author accepted manuscript version arising from this submission. JD is supported by NIHR 208398.

## Notes

### Competing Interest Statement

The authors have declared no competing interest.

### Author Declarations

This project was approved by the Health Research Authority and London Bloomsbury Research Ethics Committee (REC reference: 20/LO/0149). All patients had the opportunity to opt out of research. This project did not carry any risk to participants and was conducted retrospectively on clinically acquired data.

## References

1. Stretton, J. & Thompson, P. Frontal lobe function in temporal lobe epilepsy. Epilepsy Res. 98, 1–13 (2012).

2. Bell, B., Lin, J. J., Seidenberg, M. & Hermann, B. The neurobiology of cognitive disorders in temporal lobe epilepsy. Nat. Rev. Neurol. 7, 154–164 (2011).

3. Saling, M. M. Verbal memory in mesial temporal lobe epilepsy: beyond material specificity. Brain 132, 570–582 (2009).

4. Kramer, M. A. & Cash, S. S. Epilepsy as a disorder of cortical network organization. The Neuroscientist 18, 360–372 (2012).

5. Vrinda, M., Arun, S., Srikumar, B., Kutty, B. M. & Rao, B. S. Temporal lobe epilepsy-induced neurodegeneration and cognitive deficits: Implications for aging. J. Chem. Neuroanat. 95, 146–153 (2019).

6. Tai, X. et al. Neurodegenerative processes in temporal lobe epilepsy with hippocampal sclerosis: Clinical, pathological and neuroimaging evidence. Neuropathol. Appl. Neurobiol. 44, 70–90 (2018).

7. Hermann, B. P. et al. Cognitive prognosis in chronic temporal lobe epilepsy. Ann. Neurol. 60, 80–87 (2006).

8. Helmstaedter, C., Hermann, B., Lassonde, M., Kahane, P. & Arzimanoglou, A. Neuropsychology in the Care of People with Epilepsy. vol. 11 (John Libbey Eurotext, 2011).

9. Helmstaedter, C., Kurthen, M., Lux, S., Reuber, M. & Elger, C. E. Chronic epilepsy and cognition: a longitudinal study in temporal lobe epilepsy. Ann. Neurol. Off. J. Am. Neurol. Assoc. Child Neurol. Soc. 54, 425–432 (2003).

10. Thompson, P. J. & Duncan, J. S. Cognitive decline in severe intractable epilepsy. Epilepsia 46, 1780–1787 (2005).

11. Mohanraj, R. et al. Mortality in adults with newly diagnosed and chronic epilepsy: a retrospective comparative study. Lancet Neurol. 5, 481–487 (2006).

12. De Tisi, J. et al. The long-term outcome of adult epilepsy surgery, patterns of seizure remission, and relapse: a cohort study. The Lancet 378, 1388–1395 (2011).

13. Sherman, E. M. et al. Neuropsychological outcomes after epilepsy surgery: systematic review and pooled estimates. Epilepsia 52, 857–869 (2011).

14. Baxendale, S. & Thompson, P. The association of cognitive phenotypes with postoperative outcomes after epilepsy surgery in patients with temporal lobe epilepsy. Epilepsy Behav. 112, 107386 (2020).

15. Stroup, E. et al. Predicting verbal memory decline following anterior temporal lobectomy (ATL). Neurology 60, 1266–1273 (2003).

16. Potter, J. L. et al. Presurgical neuropsychological testing predicts cognitive and seizure outcomes after anterior temporal lobectomy. Epilepsy Behav. 16, 246– 253 (2009).

17. Elshorst, N. et al. Postoperative memory prediction in left temporal lobe epilepsy: the Wada test is of no added value to preoperative neuropsychological assessment and MRI. Epilepsy Behav. 16, 335–340 (2009).

18. Thompson, P., Baxendale, S., McEvoy, A. & Duncan, J. Cognitive outcomes of temporal lobe epilepsy surgery in older patients. Seizure 29, 41–45 (2015).

19. Baxendale, S., Thompson, P., Harkness, W. & Duncan, J. Predicting memory decline following epilepsy surgery: a multivariate approach. Epilepsia 47, 1887– 1894 (2006).

20. Reyes, A. et al. Validity of the MoCA as a cognitive screening tool in epilepsy: Are there implications for global care and research? Epilepsia Open 9, 1526–1537 (2024).

21. Reyes, A. et al. Cognitive phenotypes in temporal lobe epilepsy utilizing data-and clinically driven approaches: moving toward a new taxonomy. Epilepsia 61, 1211–1220 (2020).

22. Elverman, K. H. et al. Temporal lobe epilepsy is associated with distinct cognitive phenotypes. Epilepsy Behav. 96, 61–68 (2019).

23. Young, A. L. et al. Uncovering the heterogeneity and temporal complexity of neurodegenerative diseases with Subtype and Stage Inference. Nat. Commun. 9, 4273 (2018).

24. Jain, A. K. Data clustering: 50 years beyond K-means. Pattern Recognit. Lett. 31, 651–666 (2010).

25. Marquand, A. F., Rezek, I., Buitelaar, J. & Beckmann, C. F. Understanding heterogeneity in clinical cohorts using normative models: beyond case-control studies. Biol. Psychiatry 80, 552–561 (2016).

26. Young, A. L. et al. Data-driven neuropathological staging and subtyping of TDP-43 proteinopathies. Brain 146, 2975–2988 (2023).

27. Vogel, J. W. et al. Four distinct trajectories of tau deposition identified in Alzheimer’s disease. Nat. Med. 27, 871–881 (2021).

28. Tombaugh, T. N., Kozak, J. & Rees, L. Normative data stratified by age and education for two measures of verbal fluency: FAS and animal naming. Arch. Clin. Neuropsychol. 14, 167–177 (1999).

29. Douet, V. & Chang, L. Fornix as an imaging marker for episodic memory deficits in healthy aging and in various neurological disorders. Front. Aging Neurosci. 6, 343 (2015).

30. Aggleton, J. P. et al. Hippocampal–anterior thalamic pathways for memory: uncovering a network of direct and indirect actions. Eur. J. Neurosci. 31, 2292– 2307 (2010).

31. Aggleton, J. P., Vann, S. D. & O’Mara, S. M. Converging diencephalic and hippocampal supports for episodic memory. Neuropsychologia 191, 108728 (2023).

32. Bird, C. M. & Burgess, N. The hippocampus and memory: insights from spatial processing. Nat. Rev. Neurosci. 9, 182–194 (2008).

33. Squire, L. R. Mechanisms of memory. Science 232, 1612–1619 (1986).

34. Chelune, G. J. Hippocampal adequacy versus functional reserve: predicting memory functions following temporal lobectomy. Arch. Clin. Neuropsychol. 10, 413–432 (1995).

35. de Moura Targino, R., et al. Intersection of Brain Complexity, Functional Connectivity, and Neuropsychology: A Systematic Review. Cureus 17, (2025).

36. Binding, L. P. et al. Contribution of white matter fiber bundle damage to language change after surgery for temporal lobe epilepsy. Neurology 100, e1621–e1633 (2023).

37. Binding, L. P. et al. The impact of temporal lobe epilepsy surgery on picture naming and its relationship to network metric change. NeuroImage Clin. 38, 103444 (2023).

38. Henry, J. D. & Crawford, J. R. A meta-analytic review of verbal fluency performance following focal cortical lesions. Neuropsychology 18, 284 (2004).

39. Barba, C. et al. Temporal plus epilepsy is a major determinant of temporal lobe surgery failures. Brain 139, 444–451 (2016).

40. Hamberger, M. J. et al. Evidence for cortical reorganization of language in patients with hippocampal sclerosis. Brain 130, 2942–2950 (2007).

41. Milner, B. Interhemispheric differences in the localization of psychological processes in man. Br. Med. Bull. (1971).

42. Sundermann, E. E. et al. Female advantage in verbal memory: Evidence of sex-specific cognitive reserve. Neurology 87, 1916–1924 (2016).

43. Hodgetts, S. & Hausmann, M. Sex/gender differences in brain lateralisation and connectivity. in Sex Differences in Brain Function and Dysfunction 71–99 (Springer, 2022).

44. Voyer, D., Voyer, S. & Bryden, M. P. Magnitude of sex differences in spatial abilities: a meta-analysis and consideration of critical variables. Psychol. Bull. 117, 250 (1995).

45. Weiss, E. M., Kemmler, G., Deisenhammer, E. A., Fleischhacker, W. W. & Delazer, M. Sex differences in cognitive functions. Personal. Individ. Differ. 35, 863–875 (2003).

46. Paivio, A. Dual coding theory: Retrospect and current status. Can. J. Psychol. Can. Psychol. 45, 255 (1991).

47. McIntosh, A. R. Towards a network theory of cognition. Neural Netw. 13, 861–870 (2000).

48. Petersen, S. E. & Sporns, O. Brain networks and cognitive architectures. Neuron 88, 207–219 (2015).

49. Binding, L. P. et al. White matter resection and verbal memory deficits after temporal lobe epilepsy surgery. Brain Commun. fcag033 (2026).

50. Giampiccolo, D. et al. Thalamostriatal disconnection underpins long-term seizure freedom in frontal lobe epilepsy surgery. Brain 146, 2377–2388 (2023).

51. Taylor, P. N. et al. The imaging database for epilepsy and surgery (IDEAS). Epilepsia 66, 471–481 (2025).

52. Wechsler, D. Wechsler adult intelligence scale–. Arch. Clin. Neuropsychol. (1955).

53. Coughlan, A. & Hollows, S. The adult memory and information processing battery (AMIPB). Leeds Psychol. Dep. St James Univ. Hosp. (1985).

54. Coughlan, A., Oddy, M. & Crawford, J. BIRT memory and information processing battery (BMIPB). Lond. Brain Inj. Rehabil. Trust (2007).

55. McKenna, P. & Warrington, E. K. Graded Naming Test: Manual. (NFER-NELSON, 1983).

56. Winston, G. P. et al. Optic radiation tractography and vision in anterior temporal lobe resection. Ann. Neurol. 71, 334–341 (2012).

57. Vos, S. B. et al. Hippocampal profiling: Localized magnetic resonance imaging volumetry and T2 relaxometry for hippocampal sclerosis. Epilepsia 61, 297–309 (2020).

58. Murphy, P., Chan, E., Mo, S. & Cipolotti, L. A new revised Graded Naming Test and new normative data including older adults (80–97 years). J. Neuropsychol. 14, 449–466 (2020).

59. Fischl, B. FreeSurfer. Neuroimage 62, 774–781 (2012).

60. Desikan, R. S. et al. An automated labeling system for subdividing the human cerebral cortex on MRI scans into gyral based regions of interest. Neuroimage 31, 968–980 (2006).

