## Supplementary Materials for "Preoperative Neuropsychology Subtypes Predict Neuropsychological Change after Temporal Lobe resection"

### 1. Full Statistical Information

#### 1.1. **Stability of Subtypes**

##### 1.1.1. ML Stage distribution of cases below 50% probability

| Stage | Proportion |
| --- | --- |
| 1 | 29.55% |
| 5 | 11.36% |
| 2 | 9.09% |
| 23 | 6.82% |
| 13 | 6.82% |
| 22 | 6.82% |
| 11 | 4.55% |
| 20 | 4.55% |
| 3 | 4.55% |
| 10 | 4.55% |
| 8 | 2.27% |
| 19 | 2.27% |
| 7 | 2.27% |
| 15 | 2.27% |
| 18 | 2.27% |

##### 1.1.2. TLE side distribution of cases below 50% probability

| TLE Side | Proportion |
| --- | --- |
| Right TLE | 56.82% |
| Left TLE | 43.18% |

#### 1.2. Subtypes Characterised by Distinct Clinical, Demographic, and preoperative cognitive profiles

##### 1.2.1. Against Subtype 0

1.2.1.1. Age

| Variable | Statistic |
| --- | --- |
| ml_subtype_1 | Estimate=0.096, p-value=0.950, 95CI=-2.931:3.124 |
| ml_subtype_2 | Estimate=1.855, p-value=0.248, 95CI=-1.298:5.008 |
| ml_subtype_3 | Estimate=0.902, p-value=0.681, 95CI=-3.409:5.213 |
| TLE side Right | Estimate=1.643, p-value=0.091, 95CI=-0.263:3.549 |
| Gender: Male | Estimate=0.240, p-value=0.801, 95CI=-1.632:2.111 |
| Handedness_2.0 | Estimate=0.202, p-value=0.890, 95CI=-2.660:3.065 |
| Handedness_3.0 | Estimate=0.219, p-value=0.936, 95CI=-5.155:5.594 |
| Age of Onset | Estimate=0.306, p-value=0.000, 95CI=0.216:0.397 |
| ml_stage | Estimate=0.024, p-value=0.814, 95CI=-0.172:0.219 |
| Full Model | F(9,450)=6.025, p=0.000, R2=0.099, Breusch-Pagan=0.006, Durbin-Watson=1.935, Cook's Distance=0.046 |

1.2.1.2. Age of Onset

| Variable | Statistic |
| --- | --- |
| ml_subtype_1 | Estimate=-0.775, p-value=0.619, 95CI=-3.842:2.291 |
| ml_subtype_2 | Estimate=-2.689, p-value=0.102, 95CI=-5.915:0.537 |
| ml_subtype_3 | Estimate=-0.061, p-value=0.976, 95CI=-4.031:3.910 |
| TLE side Right | Estimate=-0.053, p-value=0.956, 95CI=-1.934:1.828 |
| Gender: Male | Estimate=0.003, p-value=0.997, 95CI=-1.888:1.894 |
| Handedness_2.0 | Estimate=-1.875, p-value=0.091, 95CI=-4.051:0.300 |
| Handedness_3.0 | Estimate=-1.951, p-value=0.512, 95CI=-7.791:3.888 |
| Age | Estimate=0.293, p-value=0.000, 95CI=0.190:0.396 |
| ml_stage | Estimate=-0.174, p-value=0.073, 95CI=-0.364:0.016 |
| Full Model | F(9,450)=4.642, p=0.000, R2=0.112, Breusch-Pagan=0.000, Durbin-Watson=2.058, Cook's Distance=0.029 |

1.2.1.3. TLE side (Left)

| Variable | Statistic |
| --- | --- |
| ml_subtype_1 | Odds Ratio=1.547, z-value=1.383, p-value=0.167, 95CI=0.834:2.873 |
| ml_subtype_2 | Odds Ratio=1.830, z-value=1.768, p-value=0.077, 95CI=0.936:3.577 |
| ml_subtype_3 | Odds Ratio=0.707, z-value=-0.863, p-value=0.388, 95CI=0.321:1.555 |
| Gender: Male | Odds Ratio=1.041, z-value=0.204, p-value=0.839, 95CI=0.709:1.528 |
| Handedness_2.0 | Odds Ratio=1.196, z-value=0.647, p-value=0.518, 95CI=0.695:2.060 |
| Handedness_3.0 | Odds Ratio=0.911, z-value=-0.143, p-value=0.886, 95CI=0.253:3.282 |
| Age of Onset | Odds Ratio=1.001, z-value=0.135, p-value=0.893, 95CI=0.982:1.021 |
| ml_stage | Odds Ratio=1.046, z-value=2.218, p-value=0.027, 95CI=1.005:1.088 |
| Age | Odds Ratio=0.984, z-value=-1.728, p-value=0.084, 95CI=0.965:1.002 |
| Full Model | Chi2(9)=22.884, p=0.006, Pseudo-R2 (McFadden)=0.036 |

1.2.1.4. Gender

| Variable | Statistic |
| --- | --- |
| ml_subtype_1 | Odds Ratio=1.501, z-value=1.292, p-value=0.196, 95CI=0.811:2.778 |
| ml_subtype_2 | Odds Ratio=0.861, z-value=-0.441, p-value=0.659, 95CI=0.444:1.671 |
| ml_subtype_3 | Odds Ratio=0.488, z-value=-1.802, p-value=0.072, 95CI=0.224:1.065 |
| TLE side Right | Odds Ratio=0.974, z-value=-0.131, p-value=0.896, 95CI=0.662:1.435 |
| Handedness_2.0 | Odds Ratio=0.847, z-value=-0.604, p-value=0.546, 95CI=0.495:1.451 |
| Handedness_3.0 | Odds Ratio=1.352, z-value=0.447, p-value=0.655, 95CI=0.361:5.066 |
| Age of Onset | Odds Ratio=0.999, z-value=-0.089, p-value=0.929, 95CI=0.980:1.018 |
| ml_stage | Odds Ratio=1.010, z-value=0.488, p-value=0.626, 95CI=0.971:1.051 |
| Age | Odds Ratio=1.003, z-value=0.274, p-value=0.784, 95CI=0.984:1.022 |
| Full Model | Chi2(9)=18.066, p=0.034, Pseudo-R2 (McFadden)=0.029 |

1.2.1.5. Handedness
         1.2.1.5.1. Handedness==1

| Handedness | 0 | 1 |
| --- | --- | --- |
| 1.0 | 68 | 194 |
| 2.0 | 7 | 34 |
| 3.0 | 1 | 6 |

1.2.1.5.2. Handedness==2

| Handedness | 0 | 1 |
| --- | --- | --- |
| 1.0 | 68 | 86 |
| 2.0 | 7 | 20 |
| 3.0 | 1 | 2 |

1.2.1.5.3. Handedness==3

| Handedness | 0 | 1 |
| --- | --- | --- |
| 1.0 | 68 | 56 |
| 2.0 | 7 | 11 |
| 3.0 | 1 | 3 |

1.2.1.6. Hippocampal Sclerosis

| Variable | Statistic |
| --- | --- |
| ml_subtype_1 | Odds Ratio=2.704, z-value=2.891, p-value=0.004, 95CI=1.378:5.306 |
| ml_subtype_2 | Odds Ratio=1.037, z-value=0.100, p-value=0.920, 95CI=0.509:2.111 |
| ml_subtype_3 | Odds Ratio=0.684, z-value=-0.890, p-value=0.374, 95CI=0.296:1.579 |
| TLE side Right | Odds Ratio=1.344, z-value=1.331, p-value=0.183, 95CI=0.870:2.075 |
| Gender: Male | Odds Ratio=0.598, z-value=-2.316, p-value=0.021, 95CI=0.387:0.924 |
| Handedness_2.0 | Odds Ratio=1.160, z-value=0.472, p-value=0.637, 95CI=0.626:2.149 |
| Handedness_3.0 | Odds Ratio=1.042, z-value=0.056, p-value=0.956, 95CI=0.247:4.401 |
| Age of Onset | Odds Ratio=0.938, z-value=-5.656, p-value=0.000, 95CI=0.918:0.959 |
| ml_stage | Odds Ratio=1.044, z-value=1.823, p-value=0.068, 95CI=0.997:1.093 |
| Age | Odds Ratio=1.014, z-value=1.311, p-value=0.190, 95CI=0.993:1.036 |
| Full Model | Chi2(10)=72.046, p=0.000, Pseudo-R2 (McFadden)=0.123 |

1.2.1.7. List Learning Preoperative Score

| Variable | Statistic |
| --- | --- |
| ml_subtype_1 | Estimate=-1.098, p-value=0.000, 95CI=-1.336:-0.860 |
| ml_subtype_2 | Estimate=-0.005, p-value=0.971, 95CI=-0.262:0.252 |
| ml_subtype_3 | Estimate=-0.262, p-value=0.086, 95CI=-0.562:0.037 |
| TLE side Right | Estimate=0.027, p-value=0.724, 95CI=-0.121:0.174 |
| Gender: Male | Estimate=-0.053, p-value=0.480, 95CI=-0.200:0.094 |
| Handedness_2.0 | Estimate=-0.081, p-value=0.441, 95CI=-0.288:0.126 |
| Handedness_3.0 | Estimate=-0.147, p-value=0.556, 95CI=-0.639:0.344 |
| Age of Onset | Estimate=0.001, p-value=0.711, 95CI=-0.006:0.009 |
| ml_stage | Estimate=-0.144, p-value=0.000, 95CI=-0.159:-0.129 |
| Age | Estimate=-0.000, p-value=0.963, 95CI=-0.007:0.007 |
| Full Model | F(10,442)=83.543, p=0.000, R2=0.654, Breusch-Pagan=0.251, Durbin-Watson=1.990, Cook's Distance=0.052 |

1.2.1.8. List Recall Preoperative Score

| Variable | Statistic |
| --- | --- |
| ml_subtype_1 | Estimate=-1.138, p-value=0.000, 95CI=-1.373:-0.903 |
| ml_subtype_2 | Estimate=-0.050, p-value=0.701, 95CI=-0.303:0.204 |
| ml_subtype_3 | Estimate=-0.252, p-value=0.095, 95CI=-0.547:0.044 |
| TLE side Right | Estimate=0.138, p-value=0.064, 95CI=-0.008:0.284 |
| Gender: Male | Estimate=-0.038, p-value=0.604, 95CI=-0.184:0.107 |
| Handedness_2.0 | Estimate=-0.151, p-value=0.147, 95CI=-0.356:0.053 |
| Handedness_3.0 | Estimate=-0.237, p-value=0.337, 95CI=-0.723:0.248 |
| Age of Onset | Estimate=0.002, p-value=0.515, 95CI=-0.005:0.010 |
| ml_stage | Estimate=-0.132, p-value=0.000, 95CI=-0.147:-0.117 |
| Age | Estimate=-0.002, p-value=0.637, 95CI=-0.009:0.005 |
| Full Model | F(10,441)=78.841, p=0.000, R2=0.641, Breusch-Pagan=0.617, Durbin-Watson=1.968, Cook's Distance=0.083 |

1.2.1.9. Design Learning Preoperative Score

| Variable | Statistic |
| --- | --- |
| ml_subtype_1 | Estimate=0.032, p-value=0.801, 95CI=-0.216:0.280 |
| ml_subtype_2 | Estimate=0.070, p-value=0.608, 95CI=-0.198:0.338 |
| ml_subtype_3 | Estimate=-1.354, p-value=0.000, 95CI=-1.672:-1.035 |
| TLE side Right | Estimate=-0.289, p-value=0.000, 95CI=-0.445:-0.133 |
| Gender: Male | Estimate=0.057, p-value=0.466, 95CI=-0.097:0.211 |
| Handedness_2.0 | Estimate=0.245, p-value=0.028, 95CI=0.026:0.463 |
| Handedness_3.0 | Estimate=0.133, p-value=0.622, 95CI=-0.398:0.664 |
| Age of Onset | Estimate=0.003, p-value=0.476, 95CI=-0.005:0.011 |
| ml_stage | Estimate=-0.096, p-value=0.000, 95CI=-0.111:-0.080 |
| Age | Estimate=0.006, p-value=0.136, 95CI=-0.002:0.013 |
| Full Model | F(10,423)=47.461p=0.000, R2=0.529, Breusch-Pagan=0.758, Durbin-Watson=2.128, Cook's Distance=0.035 |

1.2.1.10. Design Recall Preoperative Score

| Variable | Statistic |
| --- | --- |
| ml_subtype_1 | Estimate=0.277, p-value=0.204, 95CI=-0.151:0.704 |
| ml_subtype_2 | Estimate=0.196, p-value=0.405, 95CI=-0.265:0.657 |
| ml_subtype_3 | Estimate=-1.872, p-value=0.000, 95CI=-2.420:-1.324 |
| TLE side Right | Estimate=0.046, p-value=0.735, 95CI=-0.222:0.314 |
| Gender: Male | Estimate=0.127, p-value=0.347, 95CI=-0.138:0.392 |
| Handedness_2.0 | Estimate=0.096, p-value=0.615, 95CI=-0.280:0.473 |
| Handedness_3.0 | Estimate=-0.397, p-value=0.394, 95CI=-1.312:0.517 |
| Age of Onset | Estimate=-0.000, p-value=0.988, 95CI=-0.014:0.014 |
| ml_stage | Estimate=-0.097, p-value=0.000, 95CI=-0.124:-0.069 |
| Age | Estimate=-0.009, p-value=0.172, 95CI=-0.022:0.004 |
| Full Model | F(10,423)=24.263, p=0.000, R2=0.365, Breusch-Pagan=0.714, Durbin-Watson=2.058, Cook's Distance=0.466 |

1.2.1.11. Story Recall (Immediate) Preoperative Score

| Variable | Statistic |
| --- | --- |
| ml_subtype_1 | Estimate=-0.839, p-value=0.000, 95CI=-1.081:-0.597 |
| ml_subtype_2 | Estimate=-0.449, p-value=0.001, 95CI=-0.711:-0.187 |
| ml_subtype_3 | Estimate=0.004, p-value=0.979, 95CI=-0.302:0.310 |
| TLE side Right | Estimate=-0.178, p-value=0.022, 95CI=-0.329:-0.026 |
| Gender: Male | Estimate=-0.198, p-value=0.010, 95CI=-0.349:-0.047 |
| Handedness_2.0 | Estimate=-0.140, p-value=0.193, 95CI=-0.351:0.071 |
| Handedness_3.0 | Estimate=-0.039, p-value=0.896, 95CI=-0.627:0.549 |
| Age of Onset | Estimate=0.009, p-value=0.018, 95CI=0.002:0.017 |
| ml_stage | Estimate=-0.130, p-value=0.000, 95CI=-0.146:-0.114 |
| Age | Estimate=0.002, p-value=0.626, 95CI=-0.005:0.009 |
| Full Model | F(10,423)=55.999, p=0.000, R2=0.570, Breusch-Pagan=0.107, Durbin-Watson=1.853, Cook's Distance=0.077 |

1.2.1.12. Story Recall (Delayed) Preoperative Score

| Variable | Statistic |
| --- | --- |
| ml_subtype_1 | Estimate=-0.760, p-value=0.000, 95CI=-1.000:-0.520 |
| ml_subtype_2 | Estimate=-0.289, p-value=0.029, 95CI=-0.549:-0.029 |
| ml_subtype_3 | Estimate=0.151, p-value=0.328, 95CI=-0.152:0.455 |
| TLE side Right | Estimate=-0.096, p-value=0.208, 95CI=-0.247:0.054 |
| Gender: Male | Estimate=-0.153, p-value=0.045, 95CI=-0.303:-0.003 |
| Handedness_2.0 | Estimate=-0.124, p-value=0.243, 95CI=-0.334:0.085 |
| Handedness_3.0 | Estimate=-0.156, p-value=0.599, 95CI=-0.740:0.427 |
| Age of Onset | Estimate=0.007, p-value=0.080, 95CI=-0.001:0.014 |
| ml_stage | Estimate=-0.138, p-value=0.000, 95CI=-0.153:-0.122 |
| Age | Estimate=0.007, p-value=0.055, 95CI=-0.000:0.014 |
| Full Model | F(10,424)=59.959, p=0.000, R2=0.586, Breusch-Pagan=0.114, Durbin-Watson=1.877, Cook's Distance=0.076 |

1.2.1.13. Picture Naming Preoperative Score

| Variable | Statistic |
| --- | --- |
| ml_subtype_1 | Estimate=-0.282, p-value=0.053, 95CI=-0.568:0.004 |
| ml_subtype_2 | Estimate=-1.965, p-value=0.000, 95CI=-2.279:-1.650 |
| ml_subtype_3 | Estimate=-0.055, p-value=0.768, 95CI=-0.423:0.312 |
| TLE side Right | Estimate=0.305, p-value=0.002, 95CI=0.113:0.497 |
| Gender: Male | Estimate=0.011, p-value=0.907, 95CI=-0.182:0.205 |
| Handedness_2.0 | Estimate=0.180, p-value=0.242, 95CI=-0.122:0.483 |
| Handedness_3.0 | Estimate=-0.031, p-value=0.941, 95CI=-0.855:0.793 |
| Age of Onset | Estimate=-0.002, p-value=0.639, 95CI=-0.010:0.006 |
| ml_stage | Estimate=-0.168, p-value=0.000, 95CI=-0.192:-0.145 |
| Age | Estimate=-0.008, p-value=0.061, 95CI=-0.017:0.000 |
| Full Model | F(10,426)=61.761, p=0.000, R2=0.617, Breusch-Pagan=0.010, Durbin-Watson=2.058, Cook's Distance=0.077 |

1.2.1.14. Categorical Fluency Preoperative Score

| Variable | Statistic |
| --- | --- |
| ml_subtype_1 | Estimate=-0.193, p-value=0.212, 95CI=-0.498:0.111 |
| ml_subtype_2 | Estimate=-0.789, p-value=0.000, 95CI=-1.117:-0.461 |
| ml_subtype_3 | Estimate=-0.149, p-value=0.438, 95CI=-0.528:0.229 |
| TLE side Right | Estimate=-0.056, p-value=0.561, 95CI=-0.245:0.133 |
| Gender: Male | Estimate=-0.114, p-value=0.232, 95CI=-0.302:0.074 |
| Handedness_2.0 | Estimate=-0.141, p-value=0.298, 95CI=-0.407:0.125 |
| Handedness_3.0 | Estimate=-0.077, p-value=0.808, 95CI=-0.700:0.545 |
| Age of Onset | Estimate=-0.000, p-value=0.972, 95CI=-0.010:0.009 |
| ml_stage | Estimate=-0.120, p-value=0.000, 95CI=-0.140:-0.101 |
| Age | Estimate=-0.010, p-value=0.031, 95CI=-0.019:-0.001 |
| Full Model | F(10,432)=26.637, p=0.000, R2=0.381, Breusch-Pagan=0.402, Durbin-Watson=2.061, Cook's Distance=0.063 |

1.2.1.15. Letter Fluency Preoperative Score

| Variable | Statistic |
| --- | --- |
| ml_subtype_1 | Estimate=0.028, p-value=0.875, 95CI=-0.323:0.379 |
| ml_subtype_2 | Estimate=-0.797, p-value=0.000, 95CI=-1.174:-0.420 |
| ml_subtype_3 | Estimate=-0.223, p-value=0.318, 95CI=-0.660:0.215 |
| TLE side Right | Estimate=0.105, p-value=0.342, 95CI=-0.112:0.321 |
| Gender: Male | Estimate=-0.099, p-value=0.366, 95CI=-0.315:0.116 |
| Handedness_2.0 | Estimate=0.053, p-value=0.733, 95CI=-0.253:0.359 |
| Handedness_3.0 | Estimate=-0.205, p-value=0.571, 95CI=-0.915:0.505 |
| Age of Onset | Estimate=-0.006, p-value=0.290, 95CI=-0.017:0.005 |
| ml_stage | Estimate=-0.103, p-value=0.000, 95CI=-0.125:-0.080 |
| Age | Estimate=0.006, p-value=0.265, 95CI=-0.005:0.016 |
| Full Model | F(10,428)=15.811p=0.000, R2=0.270, Breusch-Pagan=0.384, Durbin-Watson=2.108, Cook's Distance=0.047 |

1.2.1.16. Digit Span Preoperative Score

| Variable | Statistic |
| --- | --- |
| ml_subtype_1 | Estimate=-0.304, p-value=0.018, 95CI=-0.555:-0.053 |
| ml_subtype_2 | Estimate=-0.550, p-value=0.000, 95CI=-0.822:-0.279 |
| ml_subtype_3 | Estimate=-0.811, p-value=0.000, 95CI=-1.126:-0.497 |
| TLE side Right | Estimate=-0.020, p-value=0.799, 95CI=-0.176:0.136 |
| Gender: Male | Estimate=0.106, p-value=0.181, 95CI=-0.049:0.261 |
| Handedness_2.0 | Estimate=-0.148, p-value=0.185, 95CI=-0.367:0.071 |
| Handedness_3.0 | Estimate=0.120, p-value=0.667, 95CI=-0.427:0.667 |
| Age of Onset | Estimate=0.001, p-value=0.731, 95CI=-0.006:0.009 |
| ml_stage | Estimate=-0.080, p-value=0.000, 95CI=-0.096:-0.064 |
| Age | Estimate=0.005, p-value=0.191, 95CI=-0.003:0.013 |
| Full Model | F(10,443)=24.159, p=0.000, R2=0.353, Breusch-Pagan=0.135, Durbin-Watson=1.993, Cook's Distance=0.089 |

##### 1.2.2. Between Subtypes

###### 1.2.2.1. Age

1.2.2.1.1. Subtype 1

| Variable | Statistic |
| --- | --- |
| ml_subtype | Estimate=-1.284, p-value=0.218, 95CI=-3.333:0.764 |
| TLE side: Right | Estimate=1.964, p-value=0.065, 95CI=-0.125:4.053 |
| Handedness_2.0 | Estimate=-0.013, p-value=0.990, 95CI=-2.081:2.054 |
| Handedness_3.0 | Estimate=0.616, p-value=0.698, 95CI=-2.498:3.729 |
| Age of Onset | Estimate=1.253, p-value=0.655, 95CI=-4.265:6.771 |
| ml_stage | Estimate=0.284, p-value=0.000, 95CI=0.185:0.383 |
| Full Model | F(7,381)=5.667, p=0.000, R2=0.090, Breusch-Pagan=0.012, Durbin-Watson=1.866, Cook's Distance=0.037 |

1.2.2.1.2. Subtype 2

| Variable | Statistic |
| --- | --- |
| ml_subtype | Estimate=1.563, p-value=0.191, 95CI=-0.781:3.908 |
| TLE side: Right | Estimate=2.128, p-value=0.046, 95CI=0.039:4.217 |
| Handedness_2.0 | Estimate=-0.159, p-value=0.882, 95CI=-2.262:1.945 |
| Handedness_3.0 | Estimate=0.642, p-value=0.686, 95CI=-2.480:3.764 |
| Age of Onset | Estimate=1.487, p-value=0.596, 95CI=-4.027:7.001 |
| ml_stage | Estimate=0.287, p-value=0.000, 95CI=0.188:0.387 |
| Full Model | F(7,381)=5.754, p=0.000, R2=0.090, Breusch-Pagan=0.012, Durbin-Watson=1.863, Cook's Distance=0.041 |

1.2.2.1.3. Subtype 3

| Variable | Statistic |
| --- | --- |
| ml_subtype | Estimate=0.095, p-value=0.954, 95CI=-3.135:3.325 |
| TLE side: Right | Estimate=2.012, p-value=0.058, 95CI=-0.067:4.090 |
| Handedness_2.0 | Estimate=-0.249, p-value=0.814, 95CI=-2.332:1.834 |
| Handedness_3.0 | Estimate=0.667, p-value=0.677, 95CI=-2.483:3.816 |
| Age of Onset | Estimate=1.275, p-value=0.657, 95CI=-4.368:6.919 |
| ml_stage | Estimate=0.282, p-value=0.000, 95CI=0.182:0.382 |
| Full Model | F(7,381)=5.206, p=0.000, R2=0.086, Breusch-Pagan=0.000, Durbin-Watson=1.872, Cook's Distance=0.058 |

###### 1.2.2.2. TLE Side (Left)

1.2.2.2.1. Subtype 1

| Variable | Statistic |
| --- | --- |
| ml_subtype | Odds Ratio=1.203, z-value=0.867, p-value=0.386, 95CI=0.792:1.826 |
| Handedness_2.0 | Odds Ratio=1.203, z-value=0.872, p-value=0.383, 95CI=0.794:1.822 |
| Handedness_3.0 | Odds Ratio=1.266, z-value=0.806, p-value=0.420, 95CI=0.714:2.245 |
| Age of Onset | Odds Ratio=0.962, z-value=-0.056, p-value=0.955, 95CI=0.250:3.699 |
| ml_stage | Odds Ratio=0.997, z-value=-0.267, p-value=0.789, 95CI=0.977:1.018 |
| Age | Odds Ratio=1.032, z-value=1.601, p-value=0.109, 95CI=0.993:1.072 |
| Full Model | Chi2(7)=10.326, p=0.171, Pseudo-R2 (McFadden)=0.019 |

1.2.2.2.2. Subtype 2

| Variable | Statistic |
| --- | --- |
| ml_subtype | Odds Ratio=1.438, z-value=1.497, p-value=0.134, 95CI=0.894:2.315 |
| Handedness_2.0 | Odds Ratio=1.278, z-value=1.172, p-value=0.241, 95CI=0.848:1.925 |
| Handedness_3.0 | Odds Ratio=1.246, z-value=0.753, p-value=0.451, 95CI=0.703:2.208 |
| Age of Onset | Odds Ratio=1.004, z-value=0.005, p-value=0.996, 95CI=0.260:3.877 |
| ml_stage | Odds Ratio=0.999, z-value=-0.086, p-value=0.931, 95CI=0.979:1.020 |
| Age | Odds Ratio=1.036, z-value=1.787, p-value=0.074, 95CI=0.997:1.077 |
| Full Model | Chi2(7)=11.845, p=0.106, Pseudo-R2 (McFadden)=0.022, |

1.2.2.2.3. Subtype 3

| Variable | Statistic |
| --- | --- |
| ml_subtype | Odds Ratio=0.440, z-value=-2.869, p-value=0.004, 95CI=0.251:0.771 |
| Handedness_2.0 | Odds Ratio=1.124, z-value=0.550, p-value=0.583, 95CI=0.741:1.706 |
| Handedness_3.0 | Odds Ratio=1.283, z-value=0.845, p-value=0.398, 95CI=0.720:2.288 |
| Age of Onset | Odds Ratio=1.089, z-value=0.123, p-value=0.902, 95CI=0.279:4.259 |
| ml_stage | Odds Ratio=0.999, z-value=-0.091, p-value=0.927, 95CI=0.978:1.020 |
| Age | Odds Ratio=1.044, z-value=2.132, p-value=0.033, 95CI=1.003:1.086 |
| Full Model | Chi2(7)=18.020, p=0.012, Pseudo-R2 (McFadden)=0.034 |

###### 1.2.2.3. Gender

1.2.2.3.1. Subtype 1

| Variable | Statistic |
| --- | --- |
| ml_subtype | Odds Ratio=2.170, z-value=3.672, p-value=0.000, 95CI=1.435:3.281 |
| TLE Side: Right | Odds Ratio=0.829, z-value=-0.882, p-value=0.378, 95CI=0.546:1.258 |
| Handedness_2.0 | Odds Ratio=0.823, z-value=-0.671, p-value=0.502, 95CI=0.466:1.453 |
| Handedness_3.0 | Odds Ratio=1.658, z-value=0.694, p-value=0.488, 95CI=0.398:6.908 |
| Age of Onset | Odds Ratio=0.993, z-value=-0.698, p-value=0.485, 95CI=0.972:1.013 |
| ml_stage | Odds Ratio=1.000, z-value=-0.000, p-value=1.000, 95CI=0.963:1.039 |
| Age | Odds Ratio=1.001, z-value=0.086, p-value=0.932, 95CI=0.981:1.021 |
| Full Model | Chi2(7)=16.399, p=0.022, Pseudo-R2 (McFadden)=0.031 |

1.2.2.3.2. Subtype 2

| Variable | Statistic |
| --- | --- |
| ml_subtype | Odds Ratio=0.721, z-value=-1.369, p-value=0.171, 95CI=0.452:1.152 |
| TLE side: Right | Odds Ratio=0.784, z-value=-1.157, p-value=0.247, 95CI=0.519:1.184 |
| Handedness_2.0 | Odds Ratio=0.801, z-value=-0.778, p-value=0.436, 95CI=0.458:1.401 |
| Handedness_3.0 | Odds Ratio=1.556, z-value=0.615, p-value=0.538, 95CI=0.381:6.359 |
| Age of Onset | Odds Ratio=0.993, z-value=-0.670, p-value=0.503, 95CI=0.973:1.014 |
| ml_stage | Odds Ratio=0.995, z-value=-0.278, p-value=0.781, 95CI=0.957:1.033 |
| Age | Odds Ratio=0.999, z-value=-0.057, p-value=0.955, 95CI=0.980:1.020 |
| Full Model | Chi2(7)=4.569, p=0.712, Pseudo-R2 (McFadden)=0.009 |

1.2.2.3.3. Subtype 3

| Variable | Statistic |
| --- | --- |
| ml_subtype | Odds Ratio=0.392, z-value=-3.268, p-value=0.001, 95CI=0.223:0.687 |
| TLE side: Right | Odds Ratio=0.898, z-value=-0.499, p-value=0.618, 95CI=0.590:1.368 |
| Handedness_2.0 | Odds Ratio=0.815, z-value=-0.711, p-value=0.477, 95CI=0.463:1.434 |
| Handedness_3.0 | Odds Ratio=1.904, z-value=0.876, p-value=0.381, 95CI=0.451:8.033 |
| Age of Onset | Odds Ratio=0.996, z-value=-0.394, p-value=0.694, 95CI=0.975:1.017 |
| ml_stage | Odds Ratio=1.012, z-value=0.602, p-value=0.547, 95CI=0.973:1.052 |
| Age | Odds Ratio=0.999, z-value=-0.132, p-value=0.895, 95CI=0.979:1.019 |
| Full Model | Chi2(7)=13.840, p=0.054, Pseudo-R2 (McFadden)=0.026 |

###### 1.2.2.4. Hippocampal Sclerosis

1.2.2.4.1. Subtype 1

| Variable | Statistic |
| --- | --- |
| ml_subtype | Odds Ratio=3.042, z-value=4.496, p-value=0.000, 95CI=1.873:4.940 |
| TLE Side: Right | Odds Ratio=1.094, z-value=0.371, p-value=0.710, 95CI=0.681:1.755 |
| Gender | Odds Ratio=0.640, z-value=-1.817, p-value=0.069, 95CI=0.395:1.036 |
| Handedness_2.0 | Odds Ratio=1.307, z-value=0.782, p-value=0.434, 95CI=0.668:2.558 |
| Handedness_3.0 | Odds Ratio=0.817, z-value=-0.268, p-value=0.789, 95CI=0.187:3.578 |
| Age of Onset | Odds Ratio=0.936, z-value=-5.393, p-value=0.000, 95CI=0.913:0.959 |
| ml_stage | Odds Ratio=1.036, z-value=1.569, p-value=0.117, 95CI=0.991:1.084 |
| Age | Odds Ratio=1.020, z-value=1.629, p-value=0.103, 95CI=0.996:1.044 |
| Full Model | Chi2(8)=54.849, p=0.000, Pseudo-R2 (McFadden)=0.114 |

1.2.2.4.2. Subtype 2

| Variable | Statistic |
| --- | --- |
| ml_subtype | Odds Ratio=0.532, z-value=-2.411, p-value=0.016, 95CI=0.319:0.889 |
| TLE Side: Right | Odds Ratio=1.010, z-value=0.041, p-value=0.968, 95CI=0.634:1.608 |
| Gender | Odds Ratio=0.762, z-value=-1.153, p-value=0.249, 95CI=0.480:1.210 |
| Handedness_2.0 | Odds Ratio=1.261, z-value=0.691, p-value=0.490, 95CI=0.654:2.431 |
| Handedness_3.0 | Odds Ratio=0.758, z-value=-0.369, p-value=0.712, 95CI=0.173:3.312 |
| Age of Onset | Odds Ratio=0.938, z-value=-5.303, p-value=0.000, 95CI=0.915:0.960 |
| ml_stage | Odds Ratio=1.025, z-value=1.084, p-value=0.278, 95CI=0.980:1.072 |
| Age | Odds Ratio=1.019, z-value=1.579, p-value=0.114, 95CI=0.995:1.043 |
| Full Model | (8)=39.385, p=0.000, Pseudo-R2 (McFadden)=0.082 |

1.2.2.4.3. Subtype 3

| Variable | Statistic |
| --- | --- |
| ml_subtype | Odds Ratio=0.372, z-value=-3.197, p-value=0.001, 95CI=0.203:0.682 |
| TLE Side: Right | Odds Ratio=1.195, z-value=0.734, p-value=0.463, 95CI=0.743:1.921 |
| Gender | Odds Ratio=0.698, z-value=-1.491, p-value=0.136, 95CI=0.435:1.120 |
| Handedness_2.0 | Odds Ratio=1.272, z-value=0.713, p-value=0.476, 95CI=0.657:2.464 |
| Handedness_3.0 | Odds Ratio=0.962, z-value=-0.051, p-value=0.959, 95CI=0.217:4.267 |
| Age of Onset | Odds Ratio=0.941, z-value=-5.030, p-value=0.000, 95CI=0.919:0.964 |
| ml_stage | Odds Ratio=1.048, z-value=2.024, p-value=0.043, 95CI=1.001:1.096 |
| Age | Odds Ratio=1.017, z-value=1.405, p-value=0.160, 95CI=0.993:1.041 |
| Full Model | Chi2(8)=43.894, p=0.000, Pseudo-R2 (McFadden)=0.092 |

###### 1.2.2.5. List Learning

1.2.2.5.1. Subtype 1

| Variable | Statistic |
| --- | --- |
| ml_subtype | Estimate=-1.001, p-value=0.000, 95CI=-1.162:-0.840 |
| TLE Side: Right | Estimate=0.064, p-value=0.418, 95CI=-0.092:0.220 |
| Gender | Estimate=-0.002, p-value=0.977, 95CI=-0.161:0.157 |
| Handedness_2.0 | Estimate=-0.100, p-value=0.295, 95CI=-0.288:0.087 |
| Handedness_3.0 | Estimate=-0.169, p-value=0.711, 95CI=-1.064:0.727 |
| Age of Onset | Estimate=0.001, p-value=0.714, 95CI=-0.006:0.009 |
| ml_stage | Estimate=-0.147, p-value=0.000, 95CI=-0.164:-0.130 |
| Age | Estimate=-0.002, p-value=0.725, 95CI=-0.010:0.007 |
| Full Model | F(8,374)=58.293, p=0.000, R2=0.603, Breusch-Pagan=0.040, Durbin-Watson=2.082, Cook's Distance=0.078 |

1.2.2.5.2. Subtype 2

| Variable | Statistic |
| --- | --- |
| ml_subtype | Estimate=0.920, p-value=0.000, 95CI=0.732:1.108 |
| TLE Side: Right | Estimate=0.182, p-value=0.032, 95CI=0.016:0.349 |
| Gender | Estimate=-0.136, p-value=0.105, 95CI=-0.300:0.029 |
| Handedness_2.0 | Estimate=-0.069, p-value=0.550, 95CI=-0.297:0.158 |
| Handedness_3.0 | Estimate=-0.022, p-value=0.936, 95CI=-0.563:0.519 |
| Age of Onset | Estimate=0.003, p-value=0.464, 95CI=-0.005:0.011 |
| ml_stage | Estimate=-0.133, p-value=0.000, 95CI=-0.148:-0.117 |
| Age | Estimate=-0.001, p-value=0.760, 95CI=-0.009:0.007 |
| Full Model | F(8,374)=56.747, p=0.000, R2=0.548, Breusch-Pagan=0.182, Durbin-Watson=2.017, Cook's Distance=0.100 |

1.2.2.5.3. Subtype 3

| Variable | Statistic |
| --- | --- |
| ml_subtype | Estimate=0.483, p-value=0.000, 95CI=0.259:0.708 |
| TLE Side: Right | Estimate=0.053, p-value=0.581, 95CI=-0.134:0.240 |
| Gender | Estimate=-0.132, p-value=0.158, 95CI=-0.314:0.051 |
| Handedness_2.0 | Estimate=-0.068, p-value=0.555, 95CI=-0.293:0.157 |
| Handedness_3.0 | Estimate=-0.221, p-value=0.651, 95CI=-1.178:0.737 |
| Age of Onset | Estimate=-0.001, p-value=0.762, 95CI=-0.010:0.007 |
| ml_stage | Estimate=-0.151, p-value=0.000, 95CI=-0.169:-0.134 |
| Age | Estimate=0.001, p-value=0.796, 95CI=-0.008:0.011 |
| Full Model | F(8,374)=40.874, p=0.000, R2=0.458, Breusch-Pagan=0.018, Durbin-Watson=2.148, Cook's Distance=0.077 |

###### 1.2.2.6. List Recall

1.2.2.6.1. Subtype 1

| Variable | Statistic |
| --- | --- |
| ml_subtype | Estimate=-1.004, p-value=0.000, 95CI=-1.164:-0.843 |
| TLE Side: Right | Estimate=0.192, p-value=0.018, 95CI=0.033:0.350 |
| Gender | Estimate=-0.039, p-value=0.627, 95CI=-0.199:0.120 |
| Handedness_2.0 | Estimate=-0.129, p-value=0.246, 95CI=-0.346:0.089 |
| Handedness_3.0 | Estimate=-0.212, p-value=0.419, 95CI=-0.728:0.303 |
| Age of Onset | Estimate=-0.001, p-value=0.900, 95CI=-0.008:0.007 |
| ml_stage | Estimate=-0.134, p-value=0.000, 95CI=-0.149:-0.120 |
| Age | Estimate=-0.002, p-value=0.570, 95CI=-0.010:0.005 |
| Full Model | F(8,373)=61.838, p=0.000, R2=0.570, Breusch-Pagan=0.517, Durbin-Watson=2.012, Cook's Distance=0.118 |

1.2.2.6.2. Subtype 2

| Variable | Statistic |
| --- | --- |
| ml_subtype | Estimate=0.894, p-value=0.000, 95CI=0.701:1.087 |
| TLE Side: Right | Estimate=0.307, p-value=0.000, 95CI=0.137:0.478 |
| Gender | Estimate=-0.174, p-value=0.042, 95CI=-0.343:-0.006 |
| Handedness_2.0 | Estimate=-0.098, p-value=0.411, 95CI=-0.331:0.136 |
| Handedness_3.0 | Estimate=-0.070, p-value=0.804, 95CI=-0.623:0.484 |
| Age of Onset | Estimate=0.001, p-value=0.814, 95CI=-0.008:0.010 |
| ml_stage | Estimate=-0.121, p-value=0.000, 95CI=-0.136:-0.105 |
| Age | Estimate=-0.002, p-value=0.649, 95CI=-0.010:0.006 |
| Full Model | F(8,373)=47.728, p=0.000, R2=0.506, Breusch-Pagan=0.117, Durbin-Watson=1.995, Cook's Distance=0.070 |

1.2.2.6.3. Subtype 3

| Variable | Statistic |
| --- | --- |
| ml_subtype | Estimate=0.523, p-value=0.000, 95CI=0.275:0.772 |
| TLE Side: Right | Estimate=0.174, p-value=0.067, 95CI=-0.012:0.360 |
| Gender | Estimate=-0.163, p-value=0.084, 95CI=-0.347:0.022 |
| Handedness_2.0 | Estimate=-0.098, p-value=0.445, 95CI=-0.350:0.154 |
| Handedness_3.0 | Estimate=-0.272, p-value=0.373, 95CI=-0.871:0.327 |
| Age of Onset | Estimate=-0.003, p-value=0.462, 95CI=-0.013:0.006 |
| ml_stage | Estimate=-0.139, p-value=0.000, 95CI=-0.156:-0.122 |
| Age | Estimate=0.001, p-value=0.908, 95CI=-0.008:0.009 |
| Full Model | F(8,373)=34.077, p=0.000, R2=0.422, Breusch-Pagan=0.649, Durbin-Watson=2.109, Cook's Distance=0.060 |

###### 1.2.2.7. Design Learning

1.2.2.7.1. Subtype 1

| Variable | Statistic |
| --- | --- |
| ml_subtype | Estimate=0.489, p-value=0.000, 95CI=0.284:0.695 |
| TLE Side: Right | Estimate=-0.465, p-value=0.000, 95CI=-0.661:-0.270 |
| Gender | Estimate=0.100, p-value=0.316, 95CI=-0.096:0.297 |
| Handedness_2.0 | Estimate=0.195, p-value=0.145, 95CI=-0.067:0.458 |
| Handedness_3.0 | Estimate=-0.084, p-value=0.798, 95CI=-0.725:0.557 |
| Age of Onset | Estimate=0.003, p-value=0.522, 95CI=-0.007:0.013 |
| ml_stage | Estimate=-0.117, p-value=0.000, 95CI=-0.135:-0.099 |
| Age | Estimate=0.005, p-value=0.284, 95CI=-0.004:0.014 |
| Full Model | F(8,357)=24.771, p=0.000, R2=0.385, Breusch-Pagan=0.004, Durbin-Watson=1.936, Cook's Distance=0.040 |

1.2.2.7.2. Subtype 2

| Variable | Statistic |
| --- | --- |
| ml_subtype | Estimate=0.332, p-value=0.004, 95CI=0.108:0.555 |
| TLE Side: Right | Estimate=-0.459, p-value=0.000, 95CI=-0.658:-0.260 |
| Gender | Estimate=0.208, p-value=0.037, 95CI=0.012:0.403 |
| Handedness_2.0 | Estimate=0.181, p-value=0.194, 95CI=-0.093:0.454 |
| Handedness_3.0 | Estimate=-0.078, p-value=0.819, 95CI=-0.743:0.587 |
| Age of Onset | Estimate=0.005, p-value=0.382, 95CI=-0.006:0.015 |
| ml_stage | Estimate=-0.115, p-value=0.000, 95CI=-0.133:-0.097 |
| Age | Estimate=0.002, p-value=0.627, 95CI=-0.007:0.012 |
| Full Model | F(8,357)=24.966, p=0.000, R2=0.359, Breusch-Pagan=0.626, Durbin-Watson=1.927, Cook's Distance=0.047 |

1.2.2.7.3. Subtype 3

| Variable | Statistic |
| --- | --- |
| ml_subtype | Estimate=-1.409, p-value=0.000, 95CI=-1.643:-1.175 |
| TLE Side: Right | Estimate=-0.314, p-value=0.000, 95CI=-0.487:-0.142 |
| Gender | Estimate=0.024, p-value=0.779, 95CI=-0.145:0.194 |
| Handedness_2.0 | Estimate=0.230, p-value=0.054, 95CI=-0.004:0.465 |
| Handedness_3.0 | Estimate=0.173, p-value=0.552, 95CI=-0.399:0.744 |
| Age of Onset | Estimate=0.008, p-value=0.079, 95CI=-0.001:0.017 |
| ml_stage | Estimate=-0.095, p-value=0.000, 95CI=-0.111:-0.079 |
| Age | Estimate=0.003, p-value=0.405, 95CI=-0.005:0.012 |
| Full Model | F(8,357)=50.095, p=0.000, R2=0.529, Breusch-Pagan=0.872, Durbin-Watson=1.990, Cook's Distance=0.048 |

###### 1.2.2.8. Design Recall

1.2.2.8.1. Subtype 1

| Variable | Statistic |
| --- | --- |
| ml_subtype | Estimate=0.855, p-value=0.000, 95CI=0.518:1.193 |
| TLE Side: Right | Estimate=-0.137, p-value=0.425, 95CI=-0.473:0.200 |
| Gender | Estimate=0.174, p-value=0.308, 95CI=-0.161:0.509 |
| Handedness_2.0 | Estimate=0.060, p-value=0.800, 95CI=-0.403:0.523 |
| Handedness_3.0 | Estimate=-0.818, p-value=0.153, 95CI=-1.943:0.306 |
| Age of Onset | Estimate=-0.006, p-value=0.502, 95CI=-0.023:0.011 |
| ml_stage | Estimate=-0.128, p-value=0.000, 95CI=-0.159:-0.097 |
| Age | Estimate=-0.008, p-value=0.336, 95CI=-0.024:0.008 |
| Full Model | F(8,357)=13.223, p=0.000, R2=0.229, Breusch-Pagan=0.601, Durbin-Watson=2.150, Cook's Distance=0.479 |

1.2.2.8.2. Subtype 2

| Variable | Statistic |
| --- | --- |
| ml_subtype | Estimate=0.366, p-value=0.065, 95CI=-0.023:0.755 |
| TLE Side: Right | Estimate=-0.143, p-value=0.419, 95CI=-0.490:0.204 |
| Gender | Estimate=0.349, p-value=0.045, 95CI=0.008:0.689 |
| Handedness_2.0 | Estimate=0.034, p-value=0.890, 95CI=-0.443:0.510 |
| Handedness_3.0 | Estimate=-0.836, p-value=0.157, 95CI=-1.994:0.322 |
| Age of Onset | Estimate=-0.004, p-value=0.632, 95CI=-0.022:0.014 |
| ml_stage | Estimate=-0.127, p-value=0.000, 95CI=-0.159:-0.095 |
| Age | Estimate=-0.012, p-value=0.173, 95CI=-0.028:0.005 |
| Full Model | F(8,357)=9.974, p=0.000, R2=0.183, Breusch-Pagan=0.543, Durbin-Watson=2.126, Cook's Distance=0.426 |

1.2.2.8.3. Subtype 3

| Variable | Statistic |
| --- | --- |
| ml_subtype | Estimate=-2.145, p-value=0.000, 95CI=-2.567:-1.724 |
| TLE Side: Right | Estimate=0.089, p-value=0.575, 95CI=-0.222:0.400 |
| Gender | Estimate=0.078, p-value=0.617, 95CI=-0.228:0.383 |
| Handedness_2.0 | Estimate=0.109, p-value=0.612, 95CI=-0.314:0.533 |
| Handedness_3.0 | Estimate=-0.436, p-value=0.406, 95CI=-1.467:0.595 |
| Age of Onset | Estimate=0.001, p-value=0.882, 95CI=-0.015:0.017 |
| ml_stage | Estimate=-0.095, p-value=0.000, 95CI=-0.124:-0.066 |
| Age | Estimate=-0.010, p-value=0.170, 95CI=-0.025:0.004 |
| Full Model | F(8,357)=24.618, p=0.000, R2=0.356, Breusch-Pagan=0.648, Durbin-Watson=2.166, Cook's Distance=0.525 |

###### 1.2.2.9. Story Recall (Immediate)

1.2.2.9.1. Subtype 1

| Variable | Statistic |
| --- | --- |
| ml_subtype | Estimate=-0.572, p-value=0.000, 95CI=-0.730:-0.413 |
| TLE Side: Right | Estimate=-0.116, p-value=0.146, 95CI=-0.273:0.041 |
| Gender | Estimate=-0.163, p-value=0.043, 95CI=-0.320:-0.006 |
| Handedness_2.0 | Estimate=-0.135, p-value=0.214, 95CI=-0.347:0.078 |
| Handedness_3.0 | Estimate=0.234, p-value=0.447, 95CI=-0.370:0.839 |
| Age of Onset | Estimate=0.009, p-value=0.034, 95CI=0.001:0.017 |
| ml_stage | Estimate=-0.124, p-value=0.000, 95CI=-0.138:-0.109 |
| Age | Estimate=0.004, p-value=0.296, 95CI=-0.004:0.012 |
| Full Model | F(8,356)=43.600, p=0.000, R2=0.495, Breusch-Pagan=0.567, Durbin-Watson=1.990, Cook's Distance=0.158 |

1.2.2.9.2. Subtype 2

| Variable | Statistic |
| --- | --- |
| ml_subtype | Estimate=0.209, p-value=0.030, 95CI=0.020:0.397 |
| TLE Side: Right | Estimate=-0.081, p-value=0.343, 95CI=-0.247:0.086 |
| Gender | Estimate=-0.264, p-value=0.002, 95CI=-0.428:-0.100 |
| Handedness_2.0 | Estimate=-0.117, p-value=0.308, 95CI=-0.343:0.109 |
| Handedness_3.0 | Estimate=0.241, p-value=0.462, 95CI=-0.402:0.884 |
| Age of Onset | Estimate=0.009, p-value=0.052, 95CI=-0.000:0.017 |
| ml_stage | Estimate=-0.120, p-value=0.000, 95CI=-0.135:-0.104 |
| Age | Estimate=0.005, p-value=0.202, 95CI=-0.003:0.013 |
| Full Model | F(8,356)=33.711, p=0.000, R2=0.431, Breusch-Pagan=0.061, Durbin-Watson=2.017, Cook's Distance=0.175 |

1.2.2.9.3. Subtype 3

| Variable | Statistic |
| --- | --- |
| ml_subtype | Estimate=0.726, p-value=0.000, 95CI=0.512:0.939 |
| TLE Side: Right | Estimate=-0.181, p-value=0.026, 95CI=-0.341:-0.021 |
| Gender | Estimate=-0.188, p-value=0.019, 95CI=-0.345:-0.031 |
| Handedness_2.0 | Estimate=-0.147, p-value=0.178, 95CI=-0.361:0.067 |
| Handedness_3.0 | Estimate=0.074, p-value=0.812, 95CI=-0.536:0.684 |
| Age of Onset | Estimate=0.007, p-value=0.113, 95CI=-0.002:0.015 |
| ml_stage | Estimate=-0.133, p-value=0.000, 95CI=-0.148:-0.118 |
| Age | Estimate=0.005, p-value=0.154, 95CI=-0.002:0.013 |
| Full Model | F(8,356)=42.371, p=0.000, R2=0.488, Breusch-Pagan=0.709, Durbin-Watson=2.003, Cook's Distance=0.158 |

###### 1.2.2.10. Story Recall (Delayed)

1.2.2.10.1. Subtype 1

| Variable | Statistic |
| --- | --- |
| ml_subtype | Estimate=-0.640, p-value=0.000, 95CI=-0.800:-0.480 |
| TLE Side: Right | Estimate=-0.082, p-value=0.310, 95CI=-0.239:0.076 |
| Gender | Estimate=-0.163, p-value=0.044, 95CI=-0.322:-0.004 |
| Handedness_2.0 | Estimate=-0.101, p-value=0.355, 95CI=-0.316:0.114 |
| Handedness_3.0 | Estimate=0.084, p-value=0.787, 95CI=-0.526:0.695 |
| Age of Onset | Estimate=0.005, p-value=0.235, 95CI=-0.003:0.013 |
| ml_stage | Estimate=-0.132, p-value=0.000, 95CI=-0.147:-0.118 |
| Age | Estimate=0.008, p-value=0.032, 95CI=0.001:0.016 |
| Full Model | F(8,357)=49.370, p=0.000, R2=0.525, Breusch-Pagan=0.392, Durbin-Watson=1.956, Cook's Distance=0.153 |

1.2.2.10.2. Subtype 2

| Variable | Statistic |
| --- | --- |
| ml_subtype | Estimate=0.263, p-value=0.006, 95CI=0.078:0.449 |
| TLE Side: Right | Estimate=-0.040, p-value=0.648, 95CI=-0.215:0.134 |
| Gender | Estimate=-0.275, p-value=0.002, 95CI=-0.450:-0.101 |
| Handedness_2.0 | Estimate=-0.081, p-value=0.478, 95CI=-0.306:0.143 |
| Handedness_3.0 | Estimate=0.100, p-value=0.865, 95CI=-1.055:1.254 |
| Age of Onset | Estimate=0.005, p-value=0.280, 95CI=-0.004:0.013 |
| ml_stage | Estimate=-0.127, p-value=0.000, 95CI=-0.143:-0.112 |
| Age | Estimate=0.010, p-value=0.027, 95CI=0.001:0.018 |
| Full Model | F(8,357)=40.128, p=0.000, R2=0.454, Breusch-Pagan=0.023, Durbin-Watson=1.972, Cook's Distance=0.170 |

1.2.2.10.3. Subtype 3

| Variable | Statistic |
| --- | --- |
| ml_subtype | Estimate=0.771, p-value=0.000, 95CI=0.554:0.988 |
| TLE Side: Right | Estimate=-0.150, p-value=0.070, 95CI=-0.312:0.012 |
| Gender | Estimate=-0.197, p-value=0.016, 95CI=-0.357:-0.037 |
| Handedness_2.0 | Estimate=-0.113, p-value=0.308, 95CI=-0.332:0.105 |
| Handedness_3.0 | Estimate=-0.088, p-value=0.780, 95CI=-0.710:0.533 |
| Age of Onset | Estimate=0.003, p-value=0.549, 95CI=-0.006:0.011 |
| ml_stage | Estimate=-0.142, p-value=0.000, 95CI=-0.157:-0.126 |
| Age | Estimate=0.010, p-value=0.011, 95CI=0.002:0.018 |
| Full Model | F(8,357)=46.366, p=0.000, R2=0.510, Breusch-Pagan=0.335, Durbin-Watson=1.992, Cook's Distance=0.153 |

###### 1.2.2.11. Picture Naming

1.2.2.11.1. Subtype 1

| Variable | Statistic |
| --- | --- |
| ml_subtype | Estimate=0.935, p-value=0.000, 95CI=0.678:1.193 |
| TLE Side: Right | Estimate=0.569, p-value=0.000, 95CI=0.314:0.823 |
| Gender | Estimate=-0.116, p-value=0.356, 95CI=-0.363:0.131 |
| Handedness_2.0 | Estimate=0.210, p-value=0.239, 95CI=-0.140:0.560 |
| Handedness_3.0 | Estimate=0.073, p-value=0.885, 95CI=-0.916:1.061 |
| Age of Onset | Estimate=0.003, p-value=0.588, 95CI=-0.008:0.014 |
| ml_stage | Estimate=-0.144, p-value=0.000, 95CI=-0.173:-0.115 |
| Age | Estimate=-0.012, p-value=0.045, 95CI=-0.023:-0.000 |
| Full Model | F(8,359)=24.221, p=0.000, R2=0.401, Breusch-Pagan=0.000, Durbin-Watson=1.902, Cook's Distance=0.084 |

1.2.2.11.2. Subtype 2

| Variable | Statistic |
| --- | --- |
| ml_subtype | Estimate=-1.728, p-value=0.000, 95CI=-1.977:-1.479 |
| TLE Side: Right | Estimate=0.395, p-value=0.000, 95CI=0.179:0.610 |
| Gender | Estimate=-0.037, p-value=0.734, 95CI=-0.248:0.175 |
| Handedness_2.0 | Estimate=0.207, p-value=0.207, 95CI=-0.115:0.529 |
| Handedness_3.0 | Estimate=-0.174, p-value=0.679, 95CI=-1.002:0.653 |
| Age of Onset | Estimate=-0.002, p-value=0.592, 95CI=-0.011:0.007 |
| ml_stage | Estimate=-0.166, p-value=0.000, 95CI=-0.190:-0.141 |
| Age | Estimate=-0.010, p-value=0.047, 95CI=-0.019:-0.000 |
| Full Model | F(8,359)=48.620, p=0.000, R2=0.552, Breusch-Pagan=0.004, Durbin-Watson=1.919, Cook's Distance=0.080 |

1.2.2.11.3. Subtype 3

| Variable | Statistic |
| --- | --- |
| ml_subtype | Estimate=0.719, p-value=0.000, 95CI=0.371:1.067 |
| TLE Side: Right | Estimate=0.468, p-value=0.000, 95CI=0.208:0.728 |
| Gender | Estimate=0.156, p-value=0.233, 95CI=-0.101:0.413 |
| Handedness_2.0 | Estimate=0.156, p-value=0.383, 95CI=-0.195:0.506 |
| Handedness_3.0 | Estimate=-0.082, p-value=0.844, 95CI=-0.902:0.737 |
| Age of Onset | Estimate=0.004, p-value=0.569, 95CI=-0.009:0.016 |
| ml_stage | Estimate=-0.153, p-value=0.000, 95CI=-0.177:-0.128 |
| Age | Estimate=-0.016, p-value=0.011, 95CI=-0.029:-0.004 |
| Full Model | F(8,359)=23.080, p=0.000, R2=0.340, Breusch-Pagan=0.802, Durbin-Watson=2.033, Cook's Distance=0.063 |

###### 1.2.2.12. Categorical Fluency

1.2.2.12.1. Subtype 1

| Variable | Statistic |
| --- | --- |
| ml_subtype | Estimate=0.341, p-value=0.001, 95CI=0.142:0.540 |
| TLE Side: Right | Estimate=-0.050, p-value=0.620, 95CI=-0.248:0.148 |
| Gender | Estimate=-0.168, p-value=0.096, 95CI=-0.367:0.030 |
| Handedness_2.0 | Estimate=-0.171, p-value=0.213, 95CI=-0.441:0.099 |
| Handedness_3.0 | Estimate=0.043, p-value=0.895, 95CI=-0.592:0.677 |
| Age of Onset | Estimate=0.000, p-value=0.947, 95CI=-0.010:0.010 |
| ml_stage | Estimate=-0.112, p-value=0.000, 95CI=-0.130:-0.094 |
| Age | Estimate=-0.011, p-value=0.026, 95CI=-0.020:-0.001 |
| Full Model | F(8,365)=21.362, p=0.000, R2=0.319, Breusch-Pagan=0.588, Durbin-Watson=2.112, Cook's Distance=0.061 |

1.2.2.12.2. Subtype 2

| Variable | Statistic |
| --- | --- |
| ml_subtype | Estimate=-0.613, p-value=0.000, 95CI=-0.832:-0.394 |
| TLE Side: Right | Estimate=-0.110, p-value=0.266, 95CI=-0.303:0.084 |
| Gender | Estimate=-0.138, p-value=0.157, 95CI=-0.328:0.053 |
| Handedness_2.0 | Estimate=-0.172, p-value=0.200, 95CI=-0.435:0.091 |
| Handedness_3.0 | Estimate=-0.043, p-value=0.893, 95CI=-0.662:0.577 |
| Age of Onset | Estimate=-0.002, p-value=0.762, 95CI=-0.011:0.008 |
| ml_stage | Estimate=-0.121, p-value=0.000, 95CI=-0.139:-0.103 |
| Age | Estimate=-0.010, p-value=0.035, 95CI=-0.019:-0.001 |
| Full Model | F(8,365)=24.720, p=0.000, R2=0.351, Breusch-Pagan=0.912, Durbin-Watson=2.120, Cook's Distance=0.058 |

1.2.2.12.3. Subtype 3

| Variable | Statistic |
| --- | --- |
| ml_subtype | Estimate=0.241, p-value=0.074, 95CI=-0.023:0.506 |
| TLE Side: Right | Estimate=-0.094, p-value=0.364, 95CI=-0.296:0.109 |
| Gender | Estimate=-0.077, p-value=0.447, 95CI=-0.278:0.123 |
| Handedness_2.0 | Estimate=-0.194, p-value=0.163, 95CI=-0.466:0.079 |
| Handedness_3.0 | Estimate=0.002, p-value=0.995, 95CI=-0.641:0.645 |
| Age of Onset | Estimate=0.000, p-value=0.937, 95CI=-0.010:0.011 |
| ml_stage | Estimate=-0.117, p-value=0.000, 95CI=-0.135:-0.098 |
| Age | Estimate=-0.012, p-value=0.017, 95CI=-0.021:-0.002 |
| Full Model | F(8,365)=19.909, p=0.000, R2=0.304, Breusch-Pagan=0.431, Durbin-Watson=2.171, Cook's Distance=0.072 |

###### 1.2.2.13. Letter Fluency

1.2.2.13.1. Subtype 1

| Variable | Statistic |
| --- | --- |
| ml_subtype | Estimate=0.612, p-value=0.000, 95CI=0.380:0.844 |
| TLE Side: Right | Estimate=0.089, p-value=0.446, 95CI=-0.141:0.319 |
| Gender | Estimate=-0.163, p-value=0.167, 95CI=-0.395:0.069 |
| Handedness_2.0 | Estimate=0.104, p-value=0.516, 95CI=-0.211:0.420 |
| Handedness_3.0 | Estimate=0.002, p-value=0.995, 95CI=-0.736:0.740 |
| Age of Onset | Estimate=-0.004, p-value=0.475, 95CI=-0.016:0.007 |
| ml_stage | Estimate=-0.096, p-value=0.000, 95CI=-0.117:-0.074 |
| Age | Estimate=0.006, p-value=0.268, 95CI=-0.005:0.017 |
| Full Model | F(8,363)=13.455, p=0.000, R2=0.229, Breusch-Pagan=0.446, Durbin-Watson=1.968, Cook's Distance=0.083 |

1.2.2.13.2. Subtype 2

| Variable | Statistic |
| --- | --- |
| ml_subtype | Estimate=-0.778, p-value=0.000, 95CI=-1.034:-0.521 |
| TLE Side: Right | Estimate=0.005, p-value=0.965, 95CI=-0.223:0.233 |
| Gender | Estimate=-0.095, p-value=0.409, 95CI=-0.320:0.131 |
| Handedness_2.0 | Estimate=0.101, p-value=0.524, 95CI=-0.211:0.414 |
| Handedness_3.0 | Estimate=-0.115, p-value=0.758, 95CI=-0.845:0.616 |
| Age of Onset | Estimate=-0.006, p-value=0.279, 95CI=-0.018:0.005 |
| ml_stage | Estimate=-0.106, p-value=0.000, 95CI=-0.128:-0.085 |
| Age | Estimate=0.006, p-value=0.253, 95CI=-0.005:0.017 |
| Full Model | F(8,363)=14.774, p=0.000, R2=0.246, Breusch-Pagan=0.391, Durbin-Watson=1.989, Cook's Distance=0.072 |

1.2.2.13.3. Subtype 3

| Variable | Statistic |
| --- | --- |
| ml_subtype | Estimate=0.022, p-value=0.893, 95CI=-0.300:0.344 |
| TLE Side: Right | Estimate=0.061, p-value=0.622, 95CI=-0.180:0.301 |
| Gender | Estimate=-0.045, p-value=0.709, 95CI=-0.285:0.194 |
| Handedness_2.0 | Estimate=0.081, p-value=0.628, 95CI=-0.246:0.408 |
| Handedness_3.0 | Estimate=-0.013, p-value=0.974, 95CI=-0.779:0.753 |
| Age of Onset | Estimate=-0.003, p-value=0.590, 95CI=-0.015:0.009 |
| ml_stage | Estimate=-0.097, p-value=0.000, 95CI=-0.120:-0.074 |
| Age | Estimate=0.004, p-value=0.457, 95CI=-0.007:0.016 |
| Full Model | F(8,363)=9.400, p=0.000, R2=0.172, Breusch-Pagan=0.633, Durbin-Watson=2.039, Cook's Distance=0.058 |

###### 1.2.2.14. Digit Span

1.2.2.14.1. Subtype 1

| Variable | Statistic |
| --- | --- |
| ml_subtype | Estimate=0.364, p-value=0.000, 95CI=0.199:0.529 |
| TLE Side: Right | Estimate=-0.108, p-value=0.195, 95CI=-0.271:0.056 |
| Gender | Estimate=0.021, p-value=0.799, 95CI=-0.143:0.185 |
| Handedness_2.0 | Estimate=-0.133, p-value=0.242, 95CI=-0.357:0.090 |
| Handedness_3.0 | Estimate=0.369, p-value=0.198, 95CI=-0.193:0.931 |
| Age of Onset | Estimate=0.002, p-value=0.688, 95CI=-0.006:0.010 |
| ml_stage | Estimate=-0.084, p-value=0.000, 95CI=-0.099:-0.069 |
| Age | Estimate=0.007, p-value=0.101, 95CI=-0.001:0.015 |
| Full Model | F(8,374)=18.828, p=0.000, R2=0.287, Breusch-Pagan=0.246, Durbin-Watson=1.971, Cook's Distance=0.033 |

1.2.2.14.2. Subtype 2

| Variable | Statistic |
| --- | --- |
| ml_subtype | Estimate=-0.142, p-value=0.142, 95CI=-0.331:0.048 |
| TLE Side: Right | Estimate=-0.131, p-value=0.125, 95CI=-0.299:0.036 |
| Gender | Estimate=0.080, p-value=0.344, 95CI=-0.086:0.245 |
| Handedness_2.0 | Estimate=-0.148, p-value=0.204, 95CI=-0.376:0.081 |
| Handedness_3.0 | Estimate=0.362, p-value=0.216, 95CI=-0.213:0.937 |
| Age of Onset | Estimate=0.002, p-value=0.671, 95CI=-0.007:0.010 |
| ml_stage | Estimate=-0.087, p-value=0.000, 95CI=-0.102:-0.071 |
| Age | Estimate=0.006, p-value=0.155, 95CI=-0.002:0.014 |
| Full Model | F(8,374)=16.050, p=0.000, R2=0.256, Breusch-Pagan=0.293, Durbin-Watson=1.952, Cook's Distance=0.043 |

1.2.2.14.3. Subtype 3

| Variable | Statistic |
| --- | --- |
| ml_subtype | Estimate=-0.446, p-value=0.000, 95CI=-0.667:-0.226 |
| TLE Side: Right | Estimate=-0.070, p-value=0.407, 95CI=-0.236:0.096 |
| Gender | Estimate=0.032, p-value=0.698, 95CI=-0.132:0.197 |
| Handedness_2.0 | Estimate=-0.139, p-value=0.225, 95CI=-0.363:0.086 |
| Handedness_3.0 | Estimate=0.418, p-value=0.146, 95CI=-0.147:0.982 |
| Age of Onset | Estimate=0.003, p-value=0.439, 95CI=-0.005:0.011 |
| ml_stage | Estimate=-0.079, p-value=0.000, 95CI=-0.094:-0.063 |
| Age | Estimate=0.006, p-value=0.169, 95CI=-0.002:0.013 |
| Full Model | F(8,374)=18.335, p=0.000, R2=0.282, Breusch-Pagan=0.147, Durbin-Watson=1.988, Cook's Distance=0.027 |

#### 1.3. Association with Postoperative Cognitive Outcome

##### 1.3.1. Against Subtype 0

1.3.1.1. List Learning Change in Score

| Variable | Statistic |
| --- | --- |
| ml_subtype_1 | Estimate=-0.519, p-value=0.004, 95CI=-0.867:-0.170 |
| ml_subtype_2 | Estimate=-0.441, p-value=0.011, 95CI=-0.781:-0.101 |
| ml_subtype_3 | Estimate=-0.348, p-value=0.085, 95CI=-0.745:0.048 |
| TLE side Right | Estimate=0.621, p-value=0.000, 95CI=0.427:0.816 |
| Gender: Male | Estimate=-0.239, p-value=0.016, 95CI=-0.434:-0.045 |
| Handedness_2.0 | Estimate=-0.113, p-value=0.413, 95CI=-0.386:0.159 |
| Handedness_3.0 | Estimate=-0.760, p-value=0.019, 95CI=-1.392:-0.127 |
| Age of Onset | Estimate=0.006, p-value=0.194, 95CI=-0.003:0.016 |
| ml_stage | Estimate=-0.070, p-value=0.000, 95CI=-0.097:-0.044 |
| Age | Estimate=-0.011, p-value=0.024, 95CI=-0.020:-0.001 |
| Preoperative Score | Estimate=-0.776, p-value=0.000, 95CI=-0.901:-0.651 |
| Full Model | F(11,416)=23.341, p=0.000, R2=0.382, Breusch-Pagan=0.608, Durbin-Watson=2.057, Cook's Distance=0.033 |

1.3.1.2. List Recall Change in Score

| Variable | Statistic |
| --- | --- |
| ml_subtype_1 | Estimate=-0.480, p-value=0.011, 95CI=-0.850:-0.111 |
| ml_subtype_2 | Estimate=-0.290, p-value=0.120, 95CI=-0.656:0.075 |
| ml_subtype_3 | Estimate=-0.294, p-value=0.174, 95CI=-0.718:0.130 |
| TLE side Right | Estimate=0.637, p-value=0.000, 95CI=0.427:0.847 |
| Gender: Male | Estimate=-0.117, p-value=0.270, 95CI=-0.326:0.091 |
| Handedness_2.0 | Estimate=0.030, p-value=0.839, 95CI=-0.263:0.323 |
| Handedness_3.0 | Estimate=-0.676, p-value=0.052, 95CI=-1.356:0.004 |
| Age of Onset | Estimate=0.001, p-value=0.849, 95CI=-0.009:0.011 |
| ml_stage | Estimate=-0.037, p-value=0.008, 95CI=-0.065:-0.010 |
| Age | Estimate=-0.008, p-value=0.123, 95CI=-0.018:0.002 |
| Preoperative Score | Estimate=-0.696, p-value=0.000, 95CI=-0.830:-0.561 |
| Full Model | F(11,416)=16.662, p=0.000, R2=0.306, Breusch-Pagan=0.855, Durbin-Watson=1.926, Cook's Distance=0.065 |

1.3.1.3. Design Learning Change in Score

| Variable | Statistic |
| --- | --- |
| ml_subtype_1 | Estimate=-0.052, p-value=0.748, 95CI=-0.373:0.268 |
| ml_subtype_2 | Estimate=-0.123, p-value=0.482, 95CI=-0.466:0.220 |
| ml_subtype_3 | Estimate=-0.301, p-value=0.177, 95CI=-0.738:0.137 |
| TLE side Right | Estimate=-0.344, p-value=0.001, 95CI=-0.545:-0.143 |
| Gender: Male | Estimate=0.348, p-value=0.001, 95CI=0.151:0.544 |
| Handedness_2.0 | Estimate=-0.084, p-value=0.558, 95CI=-0.365:0.197 |
| Handedness_3.0 | Estimate=-0.251, p-value=0.478, 95CI=-0.946:0.444 |
| Age of Onset | Estimate=-0.005, p-value=0.299, 95CI=-0.015:0.005 |
| ml_stage | Estimate=-0.067, p-value=0.000, 95CI=-0.090:-0.043 |
| Age | Estimate=-0.003, p-value=0.533, 95CI=-0.013:0.007 |
| Preoperative Score | Estimate=-0.733, p-value=0.000, 95CI=-0.856:-0.610 |
| Full Model | F(11,394)=16.874, p=0.000, R2=0.320, Breusch-Pagan=0.363, Durbin-Watson=2.028, Cook's Distance=0.047 |

1.3.1.4. Design Recall Change in Score

| Variable | Statistic |
| --- | --- |
| ml_subtype_1 | Estimate=0.147, p-value=0.455, 95CI=-0.239:0.532 |
| ml_subtype_2 | Estimate=-0.021, p-value=0.919, 95CI=-0.433:0.391 |
| ml_subtype_3 | Estimate=-0.788, p-value=0.002, 95CI=-1.296:-0.279 |
| TLE side Right | Estimate=-0.462, p-value=0.000, 95CI=-0.699:-0.224 |
| Gender: Male | Estimate=0.311, p-value=0.010, 95CI=0.074:0.547 |
| Handedness_2.0 | Estimate=0.000, p-value=0.999, 95CI=-0.335:0.336 |
| Handedness_3.0 | Estimate=-0.475, p-value=0.263, 95CI=-1.307:0.358 |
| Age of Onset | Estimate=-0.004, p-value=0.495, 95CI=-0.016:0.008 |
| ml_stage | Estimate=-0.108, p-value=0.000, 95CI=-0.134:-0.083 |
| Age | Estimate=0.001, p-value=0.866, 95CI=-0.010:0.012 |
| Preoperative Score | Estimate=-1.003, p-value=0.000, 95CI=-1.087:-0.919 |
| Full Model | F(11,393)=56.428, p=0.000, R2=0.612, Breusch-Pagan=0.394, Durbin-Watson=2.185, Cook's Distance=3.299 |

1.3.1.5. Story Recall (Immediate) Change in Score

| Variable | Statistic |
| --- | --- |
| ml_subtype_1 | Estimate=-0.410, p-value=0.021, 95CI=-0.759:-0.062 |
| ml_subtype_2 | Estimate=-0.500, p-value=0.004, 95CI=-0.843:-0.157 |
| ml_subtype_3 | Estimate=-0.226, p-value=0.261, 95CI=-0.620:0.168 |
| TLE side Right | Estimate=0.199, p-value=0.030, 95CI=0.019:0.380 |
| Gender: Male | Estimate=0.013, p-value=0.892, 95CI=-0.169:0.194 |
| Handedness_2.0 | Estimate=-0.192, p-value=0.135, 95CI=-0.443:0.060 |
| Handedness_3.0 | Estimate=-0.684, p-value=0.057, 95CI=-1.390:0.021 |
| Age of Onset | Estimate=0.007, p-value=0.129, 95CI=-0.002:0.016 |
| ml_stage | Estimate=-0.038, p-value=0.002, 95CI=-0.062:-0.014 |
| Age | Estimate=-0.001, p-value=0.803, 95CI=-0.010:0.008 |
| Preoperative Score | Estimate=-0.578, p-value=0.000, 95CI=-0.728:-0.428 |
| Full Model | F(11,392)=8.586, p=0.000, R2=0.269, Breusch-Pagan=0.003, Durbin-Watson=1.995, Cook's Distance=0.055 |

1.3.1.6. Story Recall (Delayed) Change in Score

| Variable | Statistic |
| --- | --- |
| ml_subtype_1 | Estimate=-0.401, p-value=0.016, 95CI=-0.728:-0.074 |
| ml_subtype_2 | Estimate=-0.531, p-value=0.002, 95CI=-0.864:-0.198 |
| ml_subtype_3 | Estimate=-0.287, p-value=0.150, 95CI=-0.678:0.104 |
| TLE side Right | Estimate=0.300, p-value=0.001, 95CI=0.123:0.478 |
| Gender: Male | Estimate=-0.028, p-value=0.763, 95CI=-0.209:0.153 |
| Handedness_2.0 | Estimate=-0.208, p-value=0.107, 95CI=-0.461:0.045 |
| Handedness_3.0 | Estimate=-0.645, p-value=0.017, 95CI=-1.172:-0.117 |
| Age of Onset | Estimate=0.008, p-value=0.065, 95CI=-0.001:0.017 |
| ml_stage | Estimate=-0.021, p-value=0.075, 95CI=-0.043:0.002 |
| Age | Estimate=-0.004, p-value=0.417, 95CI=-0.013:0.005 |
| Preoperative Score | Estimate=-0.471, p-value=0.000, 95CI=-0.594:-0.348 |
| Full Model | F(11,392)=9.117, p=0.000, R2=0.237, Breusch-Pagan=0.020, Durbin-Watson=2.025, Cook's Distance=0.029 |

1.3.1.7. Picture Naming Change in Score

| Variable | Statistic |
| --- | --- |
| ml_subtype_1 | Estimate=0.036, p-value=0.779, 95CI=-0.216:0.288 |
| ml_subtype_2 | Estimate=-0.128, p-value=0.487, 95CI=-0.491:0.234 |
| ml_subtype_3 | Estimate=0.164, p-value=0.323, 95CI=-0.161:0.488 |
| TLE side Right | Estimate=0.704, p-value=0.000, 95CI=0.530:0.879 |
| Gender: Male | Estimate=-0.077, p-value=0.362, 95CI=-0.244:0.090 |
| Handedness_2.0 | Estimate=-0.174, p-value=0.231, 95CI=-0.459:0.111 |
| Handedness_3.0 | Estimate=-0.320, p-value=0.319, 95CI=-0.949:0.310 |
| Age of Onset | Estimate=-0.002, p-value=0.698, 95CI=-0.010:0.007 |
| ml_stage | Estimate=-0.052, p-value=0.004, 95CI=-0.088:-0.017 |
| Age | Estimate=-0.015, p-value=0.000, 95CI=-0.023:-0.007 |
| Preoperative Score | Estimate=-0.314, p-value=0.000, 95CI=-0.454:-0.174 |
| Full Model | F(11,393)=8.450, p=0.000, R2=0.234, Breusch-Pagan=0.013, Durbin-Watson=1.921, Cook's Distance=0.245 |

1.3.1.8. Categorical Fluency Change in Score

| Variable | Statistic |
| --- | --- |
| ml_subtype_1 | Estimate=-0.305, p-value=0.055, 95CI=-0.617:0.006 |
| ml_subtype_2 | Estimate=-0.399, p-value=0.021, 95CI=-0.738:-0.061 |
| ml_subtype_3 | Estimate=-0.173, p-value=0.370, 95CI=-0.550:0.205 |
| TLE side Right | Estimate=0.247, p-value=0.011, 95CI=0.057:0.436 |
| Gender: Male | Estimate=0.033, p-value=0.735, 95CI=-0.157:0.223 |
| Handedness_2.0 | Estimate=-0.129, p-value=0.346, 95CI=-0.398:0.140 |
| Handedness_3.0 | Estimate=0.448, p-value=0.142, 95CI=-0.151:1.046 |
| Age of Onset | Estimate=-0.004, p-value=0.368, 95CI=-0.014:0.005 |
| ml_stage | Estimate=-0.046, p-value=0.000, 95CI=-0.069:-0.023 |
| Age | Estimate=-0.017, p-value=0.000, 95CI=-0.026:-0.007 |
| Preoperative Score | Estimate=-0.569, p-value=0.000, 95CI=-0.663:-0.474 |
| Full Model | F(11,390)=15.484, p=0.000, R2=0.304, Breusch-Pagan=0.116, Durbin-Watson=1.940, Cook's Distance=0.059 |

1.3.1.9. Letter Fluency Change in Score

| Variable | Statistic |
| --- | --- |
| ml_subtype_1 | Estimate=0.176, p-value=0.314, 95CI=-0.167:0.519 |
| ml_subtype_2 | Estimate=0.105, p-value=0.577, 95CI=-0.265:0.475 |
| ml_subtype_3 | Estimate=0.056, p-value=0.792, 95CI=-0.359:0.470 |
| TLE side Right | Estimate=0.326, p-value=0.002, 95CI=0.119:0.532 |
| Gender: Male | Estimate=-0.131, p-value=0.214, 95CI=-0.338:0.076 |
| Handedness_2.0 | Estimate=-0.102, p-value=0.494, 95CI=-0.394:0.190 |
| Handedness_3.0 | Estimate=0.137, p-value=0.678, 95CI=-0.510:0.784 |
| Age of Onset | Estimate=0.001, p-value=0.835, 95CI=-0.009:0.011 |
| ml_stage | Estimate=-0.044, p-value=0.000, 95CI=-0.067:-0.020 |
| Age | Estimate=-0.019, p-value=0.000, 95CI=-0.029:-0.009 |
| Preoperative Score | Estimate=-0.491, p-value=0.000, 95CI=-0.581:-0.400 |
| Full Model | F(11,384)=13.752, p=0.000, R2=0.283, Breusch-Pagan=0.159, Durbin-Watson=1.993, Cook's Distance=0.096 |

1.3.1.10. Digit Span Change in Score

| Variable | Statistic |
| --- | --- |
| ml_subtype_1 | Estimate=0.031, p-value=0.911, 95CI=-0.512:0.574 |
| ml_subtype_2 | Estimate=-0.153, p-value=0.607, 95CI=-0.738:0.432 |
| ml_subtype_3 | Estimate=-0.231, p-value=0.506, 95CI=-0.914:0.452 |
| TLE side Right | Estimate=0.051, p-value=0.759, 95CI=-0.273:0.374 |
| Gender: Male | Estimate=-0.125, p-value=0.451, 95CI=-0.451:0.201 |
| Handedness_2.0 | Estimate=-0.040, p-value=0.863, 95CI=-0.497:0.417 |
| Handedness_3.0 | Estimate=-0.463, p-value=0.488, 95CI=-1.775:0.849 |
| Age of Onset | Estimate=-0.010, p-value=0.200, 95CI=-0.026:0.005 |
| ml_stage | Estimate=-0.009, p-value=0.617, 95CI=-0.046:0.027 |
| Age | Estimate=-0.008, p-value=0.328, 95CI=-0.023:0.008 |
| Preoperative Score | Estimate=-0.294, p-value=0.004, 95CI=-0.493:-0.095 |
| Full Model | F(11,386)=1.536, p=0.116, R2=0.042, Breusch-Pagan=0.834, Durbin-Watson=2.032, Cook's Distance=0.432 |

##### 1.3.2. Between Subtypes

###### 1.3.2.1. List Learning

1.3.2.1.1. Subtype 1

| Variable | Statistic |
| --- | --- |
| ml_subtype | Estimate=-0.166, p-value=0.203, 95CI=-0.421:0.090 |
| TLE Side: Right | Estimate=0.610, p-value=0.000, 95CI=0.401:0.820 |
| Gender | Estimate=-0.231, p-value=0.032, 95CI=-0.443:-0.020 |
| Handedness_2.0 | Estimate=-0.076, p-value=0.602, 95CI=-0.365:0.212 |
| Handedness_3.0 | Estimate=-0.727, p-value=0.033, 95CI=-1.394:-0.060 |
| Age of Onset | Estimate=0.005, p-value=0.301, 95CI=-0.005:0.016 |
| ml_stage | Estimate=-0.076, p-value=0.000, 95CI=-0.104:-0.048 |
| Age | Estimate=-0.010, p-value=0.062, 95CI=-0.020:0.000 |
| Preoperative Score | Estimate=-0.825, p-value=0.000, 95CI=-0.964:-0.686 |
| Full Model | F(9,354)=24.740, p=0.000, R2=0.386, Breusch-Pagan=0.503, Durbin-Watson=2.079, Cook's Distance=0.033 |

1.3.2.1.2. Subtype 2

| Variable | Statistic |
| --- | --- |
| ml_subtype | Estimate=0.044, p-value=0.749, 95CI=-0.228:0.316 |
| TLE Side: Right | Estimate=0.618, p-value=0.000, 95CI=0.406:0.829 |
| Gender | Estimate=-0.253, p-value=0.018, 95CI=-0.462:-0.044 |
| Handedness_2.0 | Estimate=-0.070, p-value=0.632, 95CI=-0.359:0.218 |
| Handedness_3.0 | Estimate=-0.711, p-value=0.037, 95CI=-1.379:-0.043 |
| Age of Onset | Estimate=0.005, p-value=0.319, 95CI=-0.005:0.016 |
| ml_stage | Estimate=-0.069, p-value=0.000, 95CI=-0.095:-0.044 |
| Age | Estimate=-0.009, p-value=0.071, 95CI=-0.020:0.001 |
| Preoperative Score | Estimate=-0.785, p-value=0.000, 95CI=-0.916:-0.654 |
| Full Model | F(9,354)=24.465, p=0.000, R2=0.383, Breusch-Pagan=0.722, Durbin-Watson=2.085, Cook's Distance=0.033 |

1.3.2.1.3. Subtype 3

| Variable | Statistic |
| --- | --- |
| ml_subtype | Estimate=0.158, p-value=0.277, 95CI=-0.127:0.443 |
| TLE Side: Right | Estimate=0.597, p-value=0.000, 95CI=0.385:0.809 |
| Gender | Estimate=-0.235, p-value=0.029, 95CI=-0.447:-0.024 |
| Handedness_2.0 | Estimate=-0.077, p-value=0.602, 95CI=-0.365:0.212 |
| Handedness_3.0 | Estimate=-0.741, p-value=0.030, 95CI=-1.410:-0.073 |
| Age of Onset | Estimate=0.005, p-value=0.355, 95CI=-0.005:0.015 |
| ml_stage | Estimate=-0.073, p-value=0.000, 95CI=-0.099:-0.046 |
| Age | Estimate=-0.009, p-value=0.072, 95CI=-0.020:0.001 |
| Preoperative Score | Estimate=-0.787, p-value=0.000, 95CI=-0.905:-0.670 |
| Full Model | F(9,354)=24.659, p=0.000, R2=0.385, Breusch-Pagan=0.633, Durbin-Watson=2.088, Cook's Distance=0.041 |

###### 1.3.2.2. List Recall

1.3.2.2.1. Subtype 1

| Variable | Statistic |
| --- | --- |
| ml_subtype | Estimate=-0.188, p-value=0.169, 95CI=-0.455:0.080 |
| TLE Side: Right | Estimate=0.646, p-value=0.000, 95CI=0.420:0.873 |
| Gender | Estimate=-0.099, p-value=0.392, 95CI=-0.326:0.128 |
| Handedness_2.0 | Estimate=0.015, p-value=0.925, 95CI=-0.294:0.324 |
| Handedness_3.0 | Estimate=-0.712, p-value=0.051, 95CI=-1.427:0.003 |
| Age of Onset | Estimate=-0.002, p-value=0.737, 95CI=-0.013:0.009 |
| ml_stage | Estimate=-0.037, p-value=0.011, 95CI=-0.066:-0.009 |
| Age | Estimate=-0.008, p-value=0.144, 95CI=-0.019:0.003 |
| Preoperative Score | Estimate=-0.692, p-value=0.000, 95CI=-0.837:-0.547 |
| Full Model | F(9,354)=17.105, p=0.000, R2=0.303, Breusch-Pagan=0.718, Durbin-Watson=2.003, Cook's Distance=0.086 |

1.3.2.2.2. Subtype 2

| Variable | Statistic |
| --- | --- |
| ml_subtype | Estimate=0.115, p-value=0.427, 95CI=-0.169:0.398 |
| TLE Side: Right | Estimate=0.656, p-value=0.000, 95CI=0.427:0.885 |
| Gender | Estimate=-0.120, p-value=0.294, 95CI=-0.345:0.105 |
| Handedness_2.0 | Estimate=0.023, p-value=0.885, 95CI=-0.287:0.332 |
| Handedness_3.0 | Estimate=-0.687, p-value=0.060, 95CI=-1.404:0.030 |
| Age of Onset | Estimate=-0.002, p-value=0.751, 95CI=-0.013:0.009 |
| ml_stage | Estimate=-0.031, p-value=0.022, 95CI=-0.058:-0.005 |
| Age | Estimate=-0.008, p-value=0.156, 95CI=-0.019:0.003 |
| Preoperative Score | Estimate=-0.662, p-value=0.000, 95CI=-0.798:-0.526 |
| Full Model | F(9,354)=16.904, p=0.000, R2=0.301, Breusch-Pagan=0.773, Durbin-Watson=2.004, Cook's Distance=0.088 |

1.3.2.2.3. Subtype 3

| Variable | Statistic |
| --- | --- |
| ml_subtype | Estimate=0.111, p-value=0.477, 95CI=-0.196:0.418 |
| TLE Side: Right | Estimate=0.631, p-value=0.000, 95CI=0.403:0.860 |
| Gender | Estimate=-0.110, p-value=0.340, 95CI=-0.338:0.117 |
| Handedness_2.0 | Estimate=0.018, p-value=0.910, 95CI=-0.292:0.328 |
| Handedness_3.0 | Estimate=-0.717, p-value=0.051, 95CI=-1.435:0.002 |
| Age of Onset | Estimate=-0.002, p-value=0.675, 95CI=-0.014:0.009 |
| ml_stage | Estimate=-0.033, p-value=0.021, 95CI=-0.061:-0.005 |
| Age | Estimate=-0.008, p-value=0.173, 95CI=-0.018:0.003 |
| Preoperative Score | Estimate=-0.648, p-value=0.000, 95CI=-0.774:-0.522 |
| Full Model | F(9,354)=16.884, p=0.000, R2=0.300, Breusch-Pagan=0.856, Durbin-Watson=2.010, Cook's Distance=0.089 |

###### 1.3.2.3. Design Learning

1.3.2.3.1. Subtype 1

| Variable | Statistic |
| --- | --- |
| ml_subtype | Estimate=0.110, p-value=0.350, 95CI=-0.121:0.340 |
| TLE Side: Right | Estimate=-0.281, p-value=0.015, 95CI=-0.507:-0.056 |
| Gender | Estimate=0.370, p-value=0.001, 95CI=0.150:0.591 |
| Handedness_2.0 | Estimate=-0.077, p-value=0.619, 95CI=-0.383:0.228 |
| Handedness_3.0 | Estimate=-0.322, p-value=0.407, 95CI=-1.085:0.441 |
| Age of Onset | Estimate=-0.010, p-value=0.087, 95CI=-0.021:0.001 |
| ml_stage | Estimate=-0.063, p-value=0.000, 95CI=-0.087:-0.038 |
| Age | Estimate=-0.003, p-value=0.640, 95CI=-0.013:0.008 |
| Preoperative Score | Estimate=-0.679, p-value=0.000, 95CI=-0.798:-0.561 |
| Full Model | F(9,334)=16.744, p=0.000, R2=0.311, Breusch-Pagan=0.443, Durbin-Watson=2.000, Cook's Distance=0.048 |

1.3.2.3.2. Subtype 2

| Variable | Statistic |
| --- | --- |
| ml_subtype | Estimate=-0.038, p-value=0.765, 95CI=-0.290:0.213 |
| TLE Side: Right | Estimate=-0.281, p-value=0.015, 95CI=-0.508:-0.055 |
| Gender | Estimate=0.385, p-value=0.001, 95CI=0.166:0.603 |
| Handedness_2.0 | Estimate=-0.082, p-value=0.599, 95CI=-0.388:0.224 |
| Handedness_3.0 | Estimate=-0.331, p-value=0.395, 95CI=-1.095:0.434 |
| Age of Onset | Estimate=-0.010, p-value=0.084, 95CI=-0.021:0.001 |
| ml_stage | Estimate=-0.062, p-value=0.000, 95CI=-0.086:-0.037 |
| Age | Estimate=-0.003, p-value=0.604, 95CI=-0.013:0.008 |
| Preoperative Score | Estimate=-0.661, p-value=0.000, 95CI=-0.777:-0.546 |
| Full Model | F(9,334)=16.617, p=0.000, R2=0.309, Breusch-Pagan=0.367, Durbin-Watson=2.001, Cook's Distance=0.043 |

1.3.2.3.3. Subtype 3

| Variable | Statistic |
| --- | --- |
| ml_subtype | Estimate=-0.182, p-value=0.312, 95CI=-0.534:0.171 |
| TLE Side: Right | Estimate=-0.276, p-value=0.017, 95CI=-0.501:-0.050 |
| Gender | Estimate=0.374, p-value=0.001, 95CI=0.155:0.593 |
| Handedness_2.0 | Estimate=-0.066, p-value=0.674, 95CI=-0.372:0.241 |
| Handedness_3.0 | Estimate=-0.297, p-value=0.445, 95CI=-1.062:0.467 |
| Age of Onset | Estimate=-0.009, p-value=0.111, 95CI=-0.020:0.002 |
| ml_stage | Estimate=-0.063, p-value=0.000, 95CI=-0.087:-0.038 |
| Age | Estimate=-0.003, p-value=0.611, 95CI=-0.013:0.008 |
| Preoperative Score | Estimate=-0.701, p-value=0.000, 95CI=-0.836:-0.566 |
| Full Model | F(9,334)=16.768, p=0.000, R2=0.311, Breusch-Pagan=0.396, Durbin-Watson=2.003, Cook's Distance=0.050 |

###### 1.3.2.4. Design Recall

1.3.2.4.1. Subtype 1

| Variable | Statistic |
| --- | --- |
| ml_subtype | Estimate=0.408, p-value=0.006, 95CI=0.118:0.698 |
| TLE Side: Right | Estimate=-0.512, p-value=0.000, 95CI=-0.788:-0.237 |
| Gender | Estimate=0.344, p-value=0.015, 95CI=0.067:0.621 |
| Handedness_2.0 | Estimate=-0.029, p-value=0.881, 95CI=-0.412:0.354 |
| Handedness_3.0 | Estimate=-0.581, p-value=0.233, 95CI=-1.539:0.377 |
| Age of Onset | Estimate=-0.008, p-value=0.244, 95CI=-0.022:0.006 |
| ml_stage | Estimate=-0.112, p-value=0.000, 95CI=-0.140:-0.085 |
| Age | Estimate=0.004, p-value=0.517, 95CI=-0.009:0.018 |
| Preoperative Score | Estimate=-0.943, p-value=0.000, 95CI=-1.028:-0.859 |
| Full Model | F(9,333)=55.936, p=0.000, R2=0.602, Breusch-Pagan=0.205, Durbin-Watson=2.206, Cook's Distance=4.284 |

1.3.2.4.2. Subtype 2

| Variable | Statistic |
| --- | --- |
| ml_subtype | Estimate=0.002, p-value=0.992, 95CI=-0.405:0.410 |
| TLE Side: Right | Estimate=-0.531, p-value=0.004, 95CI=-0.892:-0.171 |
| Gender | Estimate=0.412, p-value=0.066, 95CI=-0.028:0.851 |
| Handedness_2.0 | Estimate=-0.029, p-value=0.872, 95CI=-0.390:0.331 |
| Handedness_3.0 | Estimate=-0.578, p-value=0.330, 95CI=-1.743:0.587 |
| Age of Onset | Estimate=-0.008, p-value=0.229, 95CI=-0.021:0.005 |
| ml_stage | Estimate=-0.108, p-value=0.003, 95CI=-0.179:-0.038 |
| Age | Estimate=0.004, p-value=0.601, 95CI=-0.010:0.017 |
| Preoperative Score | Estimate=-0.910, p-value=0.002, 95CI=-1.478:-0.343 |
| Full Model | F(9,333)=2.319, p=0.015, R2=0.593, Breusch-Pagan=0.027, Durbin-Watson=2.182, Cook's Distance=4.813 |

1.3.2.4.3. Subtype 3

| Variable | Statistic |
| --- | --- |
| ml_subtype | Estimate=-0.875, p-value=0.000, 95CI=-1.293:-0.457 |
| TLE Side: Right | Estimate=-0.437, p-value=0.002, 95CI=-0.712:-0.162 |
| Gender | Estimate=0.341, p-value=0.014, 95CI=0.069:0.612 |
| Handedness_2.0 | Estimate=0.009, p-value=0.963, 95CI=-0.369:0.387 |
| Handedness_3.0 | Estimate=-0.496, p-value=0.303, 95CI=-1.442:0.450 |
| Age of Onset | Estimate=-0.006, p-value=0.407, 95CI=-0.020:0.008 |
| ml_stage | Estimate=-0.106, p-value=0.000, 95CI=-0.134:-0.079 |
| Age | Estimate=0.002, p-value=0.720, 95CI=-0.011:0.016 |
| Preoperative Score | Estimate=-1.000, p-value=0.000, 95CI=-1.091:-0.909 |
| Full Model | F(9,333)=58.472, p=0.000, R2=0.612, Breusch-Pagan=0.842, Durbin-Watson=2.196, Cook's Distance=4.507 |

###### 1.3.2.5. Story Recall (Immediate)

1.3.2.5.1. Subtype 1

| Variable | Statistic |
| --- | --- |
| ml_subtype | Estimate=-0.026, p-value=0.823, 95CI=-0.256:0.204 |
| TLE Side: Right | Estimate=0.261, p-value=0.007, 95CI=0.071:0.450 |
| Gender | Estimate=0.011, p-value=0.912, 95CI=-0.181:0.203 |
| Handedness_2.0 | Estimate=-0.147, p-value=0.264, 95CI=-0.405:0.111 |
| Handedness_3.0 | Estimate=-0.506, p-value=0.222, 95CI=-1.318:0.307 |
| Age of Onset | Estimate=0.007, p-value=0.150, 95CI=-0.003:0.016 |
| ml_stage | Estimate=-0.035, p-value=0.004, 95CI=-0.059:-0.011 |
| Age | Estimate=-0.003, p-value=0.470, 95CI=-0.013:0.006 |
| Preoperative Score | Estimate=-0.586, p-value=0.000, 95CI=-0.751:-0.421 |
| Full Model | F(9,332)=9.164, p=0.000, R2=0.273, Breusch-Pagan=0.001, Durbin-Watson=1.992, Cook's Distance=0.059 |

1.3.2.5.2. Subtype 2

| Variable | Statistic |
| --- | --- |
| ml_subtype | Estimate=-0.124, p-value=0.305, 95CI=-0.361:0.113 |
| TLE Side: Right | Estimate=0.253, p-value=0.009, 95CI=0.064:0.442 |
| Gender | Estimate=-0.001, p-value=0.990, 95CI=-0.188:0.186 |
| Handedness_2.0 | Estimate=-0.152, p-value=0.246, 95CI=-0.408:0.105 |
| Handedness_3.0 | Estimate=-0.537, p-value=0.195, 95CI=-1.350:0.277 |
| Age of Onset | Estimate=0.006, p-value=0.186, 95CI=-0.003:0.016 |
| ml_stage | Estimate=-0.035, p-value=0.003, 95CI=-0.058:-0.012 |
| Age | Estimate=-0.003, p-value=0.521, 95CI=-0.012:0.006 |
| Preoperative Score | Estimate=-0.572, p-value=0.000, 95CI=-0.723:-0.422 |
| Full Model | F(9,332)=9.508, p=0.000, R2=0.276, Breusch-Pagan=0.003, Durbin-Watson=2.012, Cook's Distance=0.054 |

1.3.2.5.3. Subtype 3

| Variable | Statistic |
| --- | --- |
| ml_subtype | Estimate=0.239, p-value=0.117, 95CI=-0.060:0.537 |
| TLE Side: Right | Estimate=0.231, p-value=0.016, 95CI=0.043:0.420 |
| Gender | Estimate=0.028, p-value=0.770, 95CI=-0.161:0.217 |
| Handedness_2.0 | Estimate=-0.164, p-value=0.215, 95CI=-0.425:0.096 |
| Handedness_3.0 | Estimate=-0.550, p-value=0.188, 95CI=-1.369:0.270 |
| Age of Onset | Estimate=0.007, p-value=0.155, 95CI=-0.003:0.016 |
| ml_stage | Estimate=-0.042, p-value=0.001, 95CI=-0.067:-0.017 |
| Age | Estimate=-0.003, p-value=0.476, 95CI=-0.012:0.006 |
| Preoperative Score | Estimate=-0.618, p-value=0.000, 95CI=-0.774:-0.462 |
| Full Model | F(9,332)=8.891, p=0.000, R2=0.279, Breusch-Pagan=0.003, Durbin-Watson=1.994, Cook's Distance=0.065 |

###### 1.3.2.6. Story Recall (Delayed)

1.3.2.6.1. Subtype 1

| Variable | Statistic |
| --- | --- |
| ml_subtype | Estimate=-0.008, p-value=0.942, 95CI=-0.232:0.215 |
| TLE Side: Right | Estimate=0.366, p-value=0.000, 95CI=0.177:0.556 |
| Gender | Estimate=-0.007, p-value=0.942, 95CI=-0.201:0.186 |
| Handedness_2.0 | Estimate=-0.179, p-value=0.166, 95CI=-0.433:0.075 |
| Handedness_3.0 | Estimate=-0.521, p-value=0.092, 95CI=-1.128:0.085 |
| Age of Onset | Estimate=0.007, p-value=0.122, 95CI=-0.002:0.017 |
| ml_stage | Estimate=-0.024, p-value=0.039, 95CI=-0.047:-0.001 |
| Age | Estimate=-0.005, p-value=0.338, 95CI=-0.014:0.005 |
| Preoperative Score | Estimate=-0.524, p-value=0.000, 95CI=-0.665:-0.382 |
| Full Model | F(9,332)=10.495, p=0.000, R2=0.273, Breusch-Pagan=0.001, Durbin-Watson=1.999, Cook's Distance=0.051 |

1.3.2.6.2. Subtype 2

| Variable | Statistic |
| --- | --- |
| ml_subtype | Estimate=-0.136, p-value=0.263, 95CI=-0.374:0.102 |
| TLE Side: Right | Estimate=0.357, p-value=0.000, 95CI=0.169:0.545 |
| Gender | Estimate=-0.016, p-value=0.869, 95CI=-0.204:0.173 |
| Handedness_2.0 | Estimate=-0.185, p-value=0.150, 95CI=-0.437:0.067 |
| Handedness_3.0 | Estimate=-0.551, p-value=0.075, 95CI=-1.160:0.057 |
| Age of Onset | Estimate=0.007, p-value=0.149, 95CI=-0.002:0.016 |
| ml_stage | Estimate=-0.024, p-value=0.033, 95CI=-0.047:-0.002 |
| Age | Estimate=-0.004, p-value=0.363, 95CI=-0.014:0.005 |
| Preoperative Score | Estimate=-0.511, p-value=0.000, 95CI=-0.641:-0.382 |
| Full Model | F(9,332)=10.811, p=0.000, R2=0.277, Breusch-Pagan=0.002, Durbin-Watson=2.008, Cook's Distance=0.039 |

1.3.2.6.3. Subtype 3

| Variable | Statistic |
| --- | --- |
| ml_subtype | Estimate=0.224, p-value=0.153, 95CI=-0.084:0.532 |
| TLE Side: Right | Estimate=0.340, p-value=0.000, 95CI=0.153:0.528 |
| Gender | Estimate=0.011, p-value=0.908, 95CI=-0.180:0.202 |
| Handedness_2.0 | Estimate=-0.194, p-value=0.135, 95CI=-0.449:0.061 |
| Handedness_3.0 | Estimate=-0.571, p-value=0.065, 95CI=-1.178:0.035 |
| Age of Onset | Estimate=0.007, p-value=0.130, 95CI=-0.002:0.016 |
| ml_stage | Estimate=-0.031, p-value=0.012, 95CI=-0.056:-0.007 |
| Age | Estimate=-0.005, p-value=0.348, 95CI=-0.014:0.005 |
| Preoperative Score | Estimate=-0.557, p-value=0.000, 95CI=-0.697:-0.418 |
| Full Model | F(9,332)=10.400, p=0.000, R2=0.279, Breusch-Pagan=0.003, Durbin-Watson=2.003, Cook's Distance=0.043 |

###### 1.3.2.7. Picture Naming

1.3.2.7.1. Subtype 1

| Variable | Statistic |
| --- | --- |
| ml_subtype | Estimate=0.003, p-value=0.984, 95CI=-0.244:0.249 |
| TLE Side: Right | Estimate=0.685, p-value=0.000, 95CI=0.497:0.873 |
| Gender | Estimate=-0.098, p-value=0.298, 95CI=-0.282:0.086 |
| Handedness_2.0 | Estimate=-0.202, p-value=0.209, 95CI=-0.517:0.114 |
| Handedness_3.0 | Estimate=-0.336, p-value=0.343, 95CI=-1.033:0.360 |
| Age of Onset | Estimate=0.001, p-value=0.906, 95CI=-0.008:0.009 |
| ml_stage | Estimate=-0.042, p-value=0.012, 95CI=-0.074:-0.009 |
| Age | Estimate=-0.014, p-value=0.002, 95CI=-0.023:-0.005 |
| Preoperative Score | Estimate=-0.268, p-value=0.000, 95CI=-0.399:-0.137 |
| Full Model | F(9,332)=8.477, p=0.000, R2=0.215, Breusch-Pagan=0.046, Durbin-Watson=2.004, Cook's Distance=0.303 |

1.3.2.7.2. Subtype 2

| Variable | Statistic |
| --- | --- |
| ml_subtype | Estimate=-0.179, p-value=0.302, 95CI=-0.521:0.162 |
| TLE Side: Right | Estimate=0.695, p-value=0.000, 95CI=0.507:0.882 |
| Gender | Estimate=-0.105, p-value=0.270, 95CI=-0.293:0.082 |
| Handedness_2.0 | Estimate=-0.194, p-value=0.221, 95CI=-0.505:0.117 |
| Handedness_3.0 | Estimate=-0.356, p-value=0.313, 95CI=-1.047:0.336 |
| Age of Onset | Estimate=-0.000, p-value=0.988, 95CI=-0.008:0.008 |
| ml_stage | Estimate=-0.049, p-value=0.010, 95CI=-0.087:-0.012 |
| Age | Estimate=-0.014, p-value=0.003, 95CI=-0.023:-0.005 |
| Preoperative Score | Estimate=-0.305, p-value=0.000, 95CI=-0.457:-0.152 |
| Full Model | F(9,332)=8.506, p=0.000, R2=0.219, Breusch-Pagan=0.026, Durbin-Watson=1.995, Cook's Distance=0.287 |

1.3.2.7.3. Subtype 3

| Variable | Statistic |
| --- | --- |
| ml_subtype | Estimate=0.148, p-value=0.258, 95CI=-0.108:0.403 |
| TLE Side: Right | Estimate=0.674, p-value=0.000, 95CI=0.480:0.868 |
| Gender | Estimate=-0.078, p-value=0.415, 95CI=-0.267:0.110 |
| Handedness_2.0 | Estimate=-0.204, p-value=0.124, 95CI=-0.464:0.056 |
| Handedness_3.0 | Estimate=-0.359, p-value=0.226, 95CI=-0.940:0.223 |
| Age of Onset | Estimate=0.000, p-value=0.960, 95CI=-0.009:0.009 |
| ml_stage | Estimate=-0.045, p-value=0.000, 95CI=-0.066:-0.024 |
| Age | Estimate=-0.014, p-value=0.003, 95CI=-0.024:-0.005 |
| Preoperative Score | Estimate=-0.277, p-value=0.000, 95CI=-0.353:-0.200 |
| Full Model | F(9,332)=10.274, p=0.000, R2=0.218, Breusch-Pagan=0.094, Durbin-Watson=1.998, Cook's Distance=0.248 |

###### 1.3.2.8. Categorical Fluency

1.3.2.8.1. Subtype 1

| Variable | Statistic |
| --- | --- |
| ml_subtype | Estimate=-0.020, p-value=0.848, 95CI=-0.228:0.187 |
| TLE Side: Right | Estimate=0.278, p-value=0.008, 95CI=0.075:0.481 |
| Gender | Estimate=0.079, p-value=0.450, 95CI=-0.126:0.284 |
| Handedness_2.0 | Estimate=-0.112, p-value=0.431, 95CI=-0.392:0.168 |
| Handedness_3.0 | Estimate=0.566, p-value=0.076, 95CI=-0.059:1.190 |
| Age of Onset | Estimate=-0.006, p-value=0.227, 95CI=-0.016:0.004 |
| ml_stage | Estimate=-0.038, p-value=0.001, 95CI=-0.061:-0.016 |
| Age | Estimate=-0.014, p-value=0.004, 95CI=-0.024:-0.005 |
| Preoperative Score | Estimate=-0.528, p-value=0.000, 95CI=-0.634:-0.423 |
| Full Model | F(9,331)=14.104, p=0.000, R2=0.277, Breusch-Pagan=0.108, Durbin-Watson=1.940, Cook's Distance=0.076 |

1.3.2.8.2. Subtype 2

| Variable | Statistic |
| --- | --- |
| ml_subtype | Estimate=-0.112, p-value=0.358, 95CI=-0.352:0.128 |
| TLE Side: Right | Estimate=0.270, p-value=0.010, 95CI=0.066:0.473 |
| Gender | Estimate=0.068, p-value=0.506, 95CI=-0.133:0.269 |
| Handedness_2.0 | Estimate=-0.115, p-value=0.419, 95CI=-0.395:0.165 |
| Handedness_3.0 | Estimate=0.551, p-value=0.084, 95CI=-0.074:1.176 |
| Age of Onset | Estimate=-0.007, p-value=0.200, 95CI=-0.017:0.004 |
| ml_stage | Estimate=-0.042, p-value=0.000, 95CI=-0.065:-0.018 |
| Age | Estimate=-0.014, p-value=0.004, 95CI=-0.024:-0.004 |
| Preoperative Score | Estimate=-0.545, p-value=0.000, 95CI=-0.653:-0.436 |
| Full Model | F(9,331)=14.229, p=0.000, R2=0.279, Breusch-Pagan=0.087, Durbin-Watson=1.948, Cook's Distance=0.074 |

1.3.2.8.3. Subtype 3

| Variable | Statistic |
| --- | --- |
| ml_subtype | Estimate=0.167, p-value=0.211, 95CI=-0.095:0.430 |
| TLE Side: Right | Estimate=0.259, p-value=0.014, 95CI=0.054:0.464 |
| Gender | Estimate=0.098, p-value=0.346, 95CI=-0.106:0.301 |
| Handedness_2.0 | Estimate=-0.118, p-value=0.406, 95CI=-0.398:0.161 |
| Handedness_3.0 | Estimate=0.544, p-value=0.088, 95CI=-0.080:1.168 |
| Age of Onset | Estimate=-0.007, p-value=0.201, 95CI=-0.017:0.004 |
| ml_stage | Estimate=-0.042, p-value=0.000, 95CI=-0.065:-0.019 |
| Age | Estimate=-0.014, p-value=0.004, 95CI=-0.024:-0.005 |
| Preoperative Score | Estimate=-0.537, p-value=0.000, 95CI=-0.641:-0.433 |
| Full Model | F(9,331)=14.340, p=0.000, R2=0.281, Breusch-Pagan=0.115, Durbin-Watson=1.935, Cook's Distance=0.092 |

###### 1.3.2.9. Letter Fluency

1.3.2.9.1. Subtype 1

| Variable | Statistic |
| --- | --- |
| ml_subtype | Estimate=0.081, p-value=0.498, 95CI=-0.154:0.316 |
| TLE Side: Right | Estimate=0.310, p-value=0.007, 95CI=0.085:0.535 |
| Gender | Estimate=-0.109, p-value=0.348, 95CI=-0.337:0.119 |
| Handedness_2.0 | Estimate=-0.139, p-value=0.381, 95CI=-0.451:0.173 |
| Handedness_3.0 | Estimate=0.302, p-value=0.390, 95CI=-0.389:0.992 |
| Age of Onset | Estimate=0.002, p-value=0.685, 95CI=-0.009:0.013 |
| ml_stage | Estimate=-0.044, p-value=0.000, 95CI=-0.067:-0.021 |
| Age | Estimate=-0.019, p-value=0.001, 95CI=-0.030:-0.008 |
| Preoperative Score | Estimate=-0.484, p-value=0.000, 95CI=-0.584:-0.384 |
| Full Model | F(9,328)=13.027, p=0.000, R2=0.263, Breusch-Pagan=0.099, Durbin-Watson=1.890, Cook's Distance=0.109 |

1.3.2.9.2. Subtype 2

| Variable | Statistic |
| --- | --- |
| ml_subtype | Estimate=-0.025, p-value=0.855, 95CI=-0.289:0.240 |
| TLE Side: Right | Estimate=0.304, p-value=0.008, 95CI=0.079:0.530 |
| Gender | Estimate=-0.094, p-value=0.409, 95CI=-0.318:0.130 |
| Handedness_2.0 | Estimate=-0.140, p-value=0.377, 95CI=-0.452:0.172 |
| Handedness_3.0 | Estimate=0.299, p-value=0.397, 95CI=-0.393:0.991 |
| Age of Onset | Estimate=0.002, p-value=0.681, 95CI=-0.009:0.014 |
| ml_stage | Estimate=-0.044, p-value=0.000, 95CI=-0.068:-0.020 |
| Age | Estimate=-0.019, p-value=0.001, 95CI=-0.030:-0.008 |
| Preoperative Score | Estimate=-0.477, p-value=0.000, 95CI=-0.579:-0.376 |
| Full Model | F(9,328)=12.963, p=0.000, R2=0.262, Breusch-Pagan=0.110, Durbin-Watson=1.888, Cook's Distance=0.099 |

1.3.2.9.3. Subtype 3

| Variable | Statistic |
| --- | --- |
| ml_subtype | Estimate=-0.097, p-value=0.518, 95CI=-0.392:0.198 |
| TLE Side: Right | Estimate=0.317, p-value=0.006, 95CI=0.090:0.545 |
| Gender | Estimate=-0.106, p-value=0.359, 95CI=-0.332:0.121 |
| Handedness_2.0 | Estimate=-0.139, p-value=0.382, 95CI=-0.450:0.173 |
| Handedness_3.0 | Estimate=0.316, p-value=0.370, 95CI=-0.376:1.008 |
| Age of Onset | Estimate=0.003, p-value=0.639, 95CI=-0.009:0.014 |
| ml_stage | Estimate=-0.042, p-value=0.001, 95CI=-0.065:-0.018 |
| Age | Estimate=-0.019, p-value=0.001, 95CI=-0.030:-0.008 |
| Preoperative Score | Estimate=-0.474, p-value=0.000, 95CI=-0.571:-0.378 |
| Full Model | F(9,328)=13.021, p=0.000, R2=0.263, Breusch-Pagan=0.103, Durbin-Watson=1.895, Cook's Distance=0.108 |

###### 1.3.2.10. Digit Span

1.3.2.10.1. Subtype 1

| Variable | Statistic |
| --- | --- |
| ml_subtype | Estimate=0.204, p-value=0.293, 95CI=-0.177:0.584 |
| TLE Side: Right | Estimate=0.003, p-value=0.988, 95CI=-0.366:0.371 |
| Gender | Estimate=-0.113, p-value=0.551, 95CI=-0.487:0.260 |
| Handedness_2.0 | Estimate=-0.078, p-value=0.766, 95CI=-0.590:0.435 |
| Handedness_3.0 | Estimate=-0.477, p-value=0.499, 95CI=-1.865:0.911 |
| Age of Onset | Estimate=-0.013, p-value=0.140, 95CI=-0.031:0.004 |
| ml_stage | Estimate=-0.009, p-value=0.650, 95CI=-0.049:0.030 |
| Age | Estimate=-0.009, p-value=0.313, 95CI=-0.026:0.008 |
| Preoperative Score | Estimate=-0.270, p-value=0.022, 95CI=-0.502:-0.039 |
| Full Model | F(9,331)=1.411, p=0.182, R2=0.037, Breusch-Pagan=0.684, Durbin-Watson=1.945, Cook's Distance=0.536 |

1.3.2.10.2. Subtype 2

| Variable | Statistic |
| --- | --- |
| ml_subtype | Estimate=-0.128, p-value=0.550, 95CI=-0.548:0.292 |
| TLE Side: Right | Estimate=-0.007, p-value=0.971, 95CI=-0.378:0.364 |
| Gender | Estimate=-0.081, p-value=0.666, 95CI=-0.448:0.286 |
| Handedness_2.0 | Estimate=-0.081, p-value=0.757, 95CI=-0.594:0.433 |
| Handedness_3.0 | Estimate=-0.488, p-value=0.490, 95CI=-1.878:0.902 |
| Age of Onset | Estimate=-0.014, p-value=0.138, 95CI=-0.032:0.004 |
| ml_stage | Estimate=-0.009, p-value=0.664, 95CI=-0.049:0.031 |
| Age | Estimate=-0.009, p-value=0.293, 95CI=-0.027:0.008 |
| Preoperative Score | Estimate=-0.248, p-value=0.033, 95CI=-0.475:-0.021 |
| Full Model | F(9,331)=1.325, p=0.223, R2=0.035, Breusch-Pagan=0.776, Durbin-Watson=1.946, Cook's Distance=0.473 |

1.3.2.10.3. Subtype 3

| Variable | Statistic |
| --- | --- |
| ml_subtype | Estimate=-0.170, p-value=0.505, 95CI=-0.670:0.330 |
| TLE Side: Right | Estimate=0.018, p-value=0.925, 95CI=-0.353:0.389 |
| Gender | Estimate=-0.097, p-value=0.608, 95CI=-0.470:0.276 |
| Handedness_2.0 | Estimate=-0.074, p-value=0.778, 95CI=-0.587:0.440 |
| Handedness_3.0 | Estimate=-0.460, p-value=0.516, 95CI=-1.850:0.930 |
| Age of Onset | Estimate=-0.013, p-value=0.168, 95CI=-0.031:0.005 |
| ml_stage | Estimate=-0.006, p-value=0.763, 95CI=-0.045:0.033 |
| Age | Estimate=-0.010, p-value=0.276, 95CI=-0.027:0.008 |
| Preoperative Score | Estimate=-0.262, p-value=0.027, 95CI=-0.494:-0.029 |
| Full Model | F(9,331)=1.335, p=0.217, R2=0.035, Breusch-Pagan=0.772, Durbin-Watson=1.954, Cook's Distance=0.471 |

#### 1.4. Postoperative Seziure Outcome

| Variable | Statistic |
| --- | --- |
| Age of Onset | coef=-0.01, exp(coef)=0.99, se(coef)=0.01, coef lower 95%=-0.02, coef upper 95%=-0.00, exp(coef) lower 95%=0.98, exp(coef) upper 95%=1.00, cmp to=0.00, z=-2.36, p=0.02, -log2(p)=5.76 |
| ml_stage | coef=-0.01, exp(coef)=0.99, se(coef)=0.01, coef lower 95%=-0.03, coef upper 95%=0.01, exp(coef) lower 95%=0.97, exp(coef) upper 95%=1.01, cmp to=0.00, z=-0.90, p=0.37, -log2(p)=1.45 |
| Age | coef=-0.00, exp(coef)=1.00, se(coef)=0.01, coef lower 95%=-0.01, coef upper 95%=0.01, exp(coef) lower 95%=0.99, exp(coef) upper 95%=1.01, cmp to=0.00, z=-0.02, p=0.98, -log2(p)=0.03 |
| ml_subtype_1 | coef=0.03, exp(coef)=1.03, se(coef)=0.18, coef lower 95%=-0.32, coef upper 95%=0.38, exp(coef) lower 95%=0.73, exp(coef) upper 95%=1.47, cmp to=0.00, z=0.18, p=0.86, -log2(p)=0.22 |
| ml_subtype_2 | coef=0.30, exp(coef)=1.35, se(coef)=0.19, coef lower 95%=-0.08, coef upper 95%=0.68, exp(coef) lower 95%=0.93, exp(coef) upper 95%=1.98, cmp to=0.00, z=1.56, p=0.12, -log2(p)=3.08 |
| ml_subtype_3 | coef=0.24, exp(coef)=1.27, se(coef)=0.22, coef lower 95%=-0.20, coef upper 95%=0.68, exp(coef) lower 95%=0.82, exp(coef) upper 95%=1.98, cmp to=0.00, z=1.08, p=0.28, -log2(p)=1.82 |
| TLE side: Right | coef=-0.01, exp(coef)=0.99, se(coef)=0.11, coef lower 95%=-0.23, coef upper 95%=0.20, exp(coef) lower 95%=0.79, exp(coef) upper 95%=1.23, cmp to=0.00, z=-0.12, p=0.90, -log2(p)=0.15 |
| Gender: 1 | coef=0.04, exp(coef)=1.04, se(coef)=0.11, coef lower 95%=-0.18, coef upper 95%=0.26, exp(coef) lower 95%=0.84, exp(coef) upper 95%=1.29, cmp to=0.00, z=0.36, p=0.72, -log2(p)=0.48 |
| Handedness_2 | coef=-0.12, exp(coef)=0.89, se(coef)=0.16, coef lower 95%=-0.43, coef upper 95%=0.19, exp(coef) lower 95%=0.65, exp(coef) upper 95%=1.21, cmp to=0.00, z=-0.73, p=0.46, -log2(p)=1.11 |
| Handedness_3 | coef=-0.08, exp(coef)=0.92, se(coef)=0.39, coef lower 95%=-0.84, coef upper 95%=0.67, exp(coef) lower 95%=0.43, exp(coef) upper 95%=1.96, cmp to=0.00, z=-0.22, p=0.83, -log2(p)=0.27 |
| Full Model | Concordance=0.56, Partial AIC=3818.37, log-likelihood ratio test=12.53 on 10 df, -log2(p) of ll-ratio test=1.99 |

### 2. Unadjusted Tables

#### 2.1. Clinical/Demographic

|  | **Subtype 0** | **Verbal Memory Subtype** | **Naming Subtype** | **Visual Memory Subtype** |
| --- | --- | --- | --- | --- |
| Age | 36.28 (10.33) | 35.82 (9.99) | 36.86 (9.82) | 35.89 (12.20) |
| Onset Age | 15.37 (11.02) | 13.01 (10.58) | 11.62 (9.33) | 13.62 (10.75) |
| List learning | 0.10 (0.84) | -2.02 (1.00) | -0.71 (1.10) | -1.46 (1.24) |
| List recall | 0.16 (0.74) | -1.95 (0.97) | -0.68 (0.98) | -1.35 (1.28) |
| Design learning | 0.45 (1.19) | -0.25 (0.93) | -0.06 (0.94) | -1.98 (1.11) |
| Design recall | 0.57 (2.81) | -0.12 (1.77) | -0.08 (0.77) | -2.55 (1.39) |
| Story recall (immediate) | 0.29 (0.99) | -1.47 (0.91) | -0.90 (0.98) | -1.01 (1.27) |
| Story recall (delayed) | 0.13 (0.95) | -1.66 (0.94) | -1.01 (1.02) | -1.09 (1.32) |
| Graded Naming Test | -0.04 (0.83) | -1.59 (1.55) | -2.98 (1.13) | -1.72 (1.06) |
| Category fluency | 0.48 (1.22) | -0.68 (1.07) | -0.99 (1.13) | -0.84 (1.35) |
| Letter fluency | 0.58 (1.21) | -0.09 (1.19) | -0.70 (1.24) | -0.42 (1.25) |
| Digit span | -0.02 (0.91) | -0.82 (0.95) | -0.95 (0.94) | -1.51 (0.77) |

#### 2.2. Postoperative Outcome

|  | **Subtype 0** | **Verbal Memory Subtype** | **Naming Subtype** | **Visual Memory Subtype** |
| --- | --- | --- | --- | --- |
| List learning 12 months change | -0.37 (1.19) | 0.16 (1.13) | -0.69 (1.30) | -0.13 (1.27) |
| List recall 12 months change | -0.05 (3.49) | 0.17 (1.21) | -0.42 (1.22) | 0.01 (1.39) |
| Design learning 12 months change | -0.27 (1.00) | -0.19 (1.15) | -0.32 (1.20) | 0.43 (1.24) |
| Design recall 12 months change | -0.20 (1.06) | -0.28 (2.29) | -0.25 (1.24) | 0.75 (1.33) |
| Story recall (immediate) 12 months change | -0.19 (1.10) | 0.15 (0.91) | -0.21 (1.08) | -0.11 (1.14) |
| Story recall (delayed) 12 months change | -0.05 (0.95) | 0.23 (0.89) | -0.17 (1.13) | -0.07 (1.24) |
| Picture Naming 12 months change | -0.15 (0.90) | -0.13 (0.92) | 0.34 (0.96) | -0.05 (1.07) |
| Category fluency 12 months change | 0.07 (1.28) | 0.17 (1.16) | 0.26 (1.03) | 0.32 (1.19) |
| Letter fluency 12 months change | -0.00 (1.12) | 0.16 (1.22) | 0.43 (1.12) | 0.18 (1.13) |
| Digit span 12 Months change | 0.06 (0.98) | 0.23 (2.09) | 0.14 (0.80) | 0.22 (0.86) |

### 3. In-sample clinical vs. SuStaIn Phenotype models

To evaluate the incremental predictive value of the SuStaIn phenotypes, we compared nested linear regression models using an analysis of variance (ANOVA). The baseline models included standard clinical covariates (sex, age at onset, age at surgery, TLE side, handedness, and preoperative cognitive scores). These were compared against full models that additionally incorporated the SuStaIn disease stage and subtype. As illustrated in Figure 2, the inclusion of SuStaIn phenotypes significantly improved the variance explained across all measured postoperative cognitive scores. The most substantial improvements in predictive accuracy were observed in the DesignA6 and ListA15 tasks. All full models demonstrated a statistically significant improvement over their respective baseline models (all *p* < 0.01), underscoring the added prognostic value of SuStaIn subtyping and staging in the preoperative assessment of TLE.

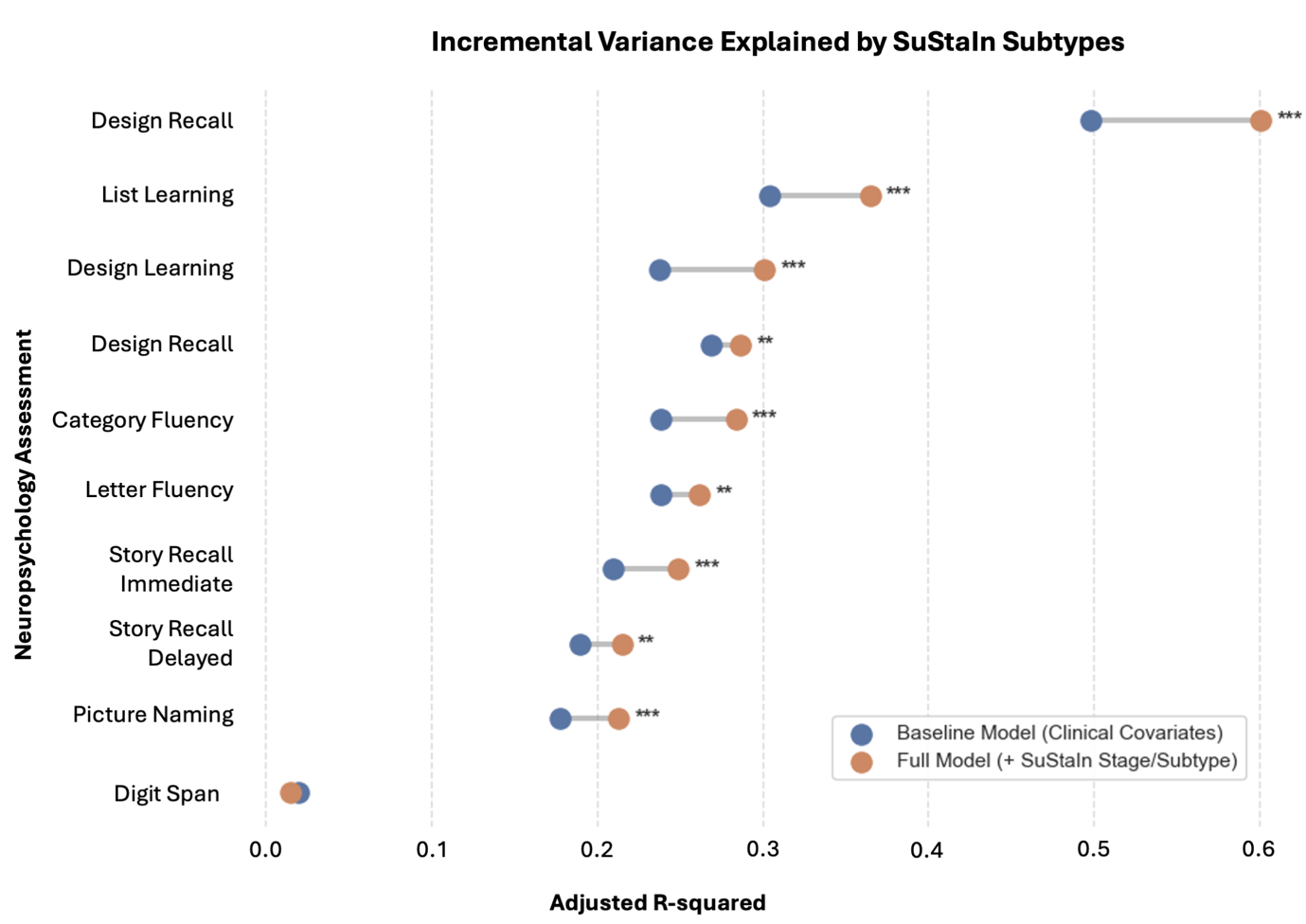

**Supplementary Figure 1. Incremental in-sample variance explained by SuStaIn phenotypes in postoperative cognitive outcomes.** The dumbbell plot illustrates the adjusted R-squared values for nested linear regression models predicting postoperative neuropsychological scores at 12 months. Blue circles represent the baseline models, which include standard clinical covariates (sex, age at onset, age at surgery, TLE side, handedness, and preoperative cognitive scores). Orange circles represent the full models, which additionally incorporate SuStaIn disease stage and subtype. The grey connecting line visually represents the incremental variance gained by including the SuStaIn phenotypes. Significance levels for the model comparisons (ANOVA) are denoted by asterisks: p < 0.01, *** p < 0.001.

### 4. Floor effects

To empirically evaluate whether the predictive capacity of SuStaIn disease stage on 12-month postoperative cognitive change was confounded by preoperative floor effects (limited capacity to decline), we performed iterative sensitivity analyses across nine neuropsychological tests. Preoperative raw score exclusion thresholds were raised incrementally from 0 (all available cases included) to 8 (excluding all patients scoring ≤8 preoperatively), and full linear regression models were re-estimated at each step.

Across the majority of cognitive domains, SuStaIn disease stage demonstrated robust predictive capacity that remained statistically significant (*p*<0.05) regardless of the applied raw score exclusion threshold (Supplementary Figure 2). However, a threshold-sensitive pattern was observed for ListA6, where SuStaIn stage served as a significant predictor of 12-month decline from baseline through an exclusion threshold of ≤5 (N=346, *p* = 0.047). Statistical significance temporarily attenuated at exclusion thresholds of 6 (N = 307, p = 0.076) and 7 (N= 268, p = 0.084), before returning below 0.05 at threshold 8 (N=223, p = 0.028). This trajectory indicates that while baseline impairment level contributes to the association, power reduction resulting from sample truncation also influences *p*-value fluctuations at higher exclusion levels. Conversely, for Story Delayed, SuStaIn stage was not a statistically significant independent predictor in the baseline model and subsequent exclusions of lower-scoring cases led to expected increases in p-values as statistical power diminished.

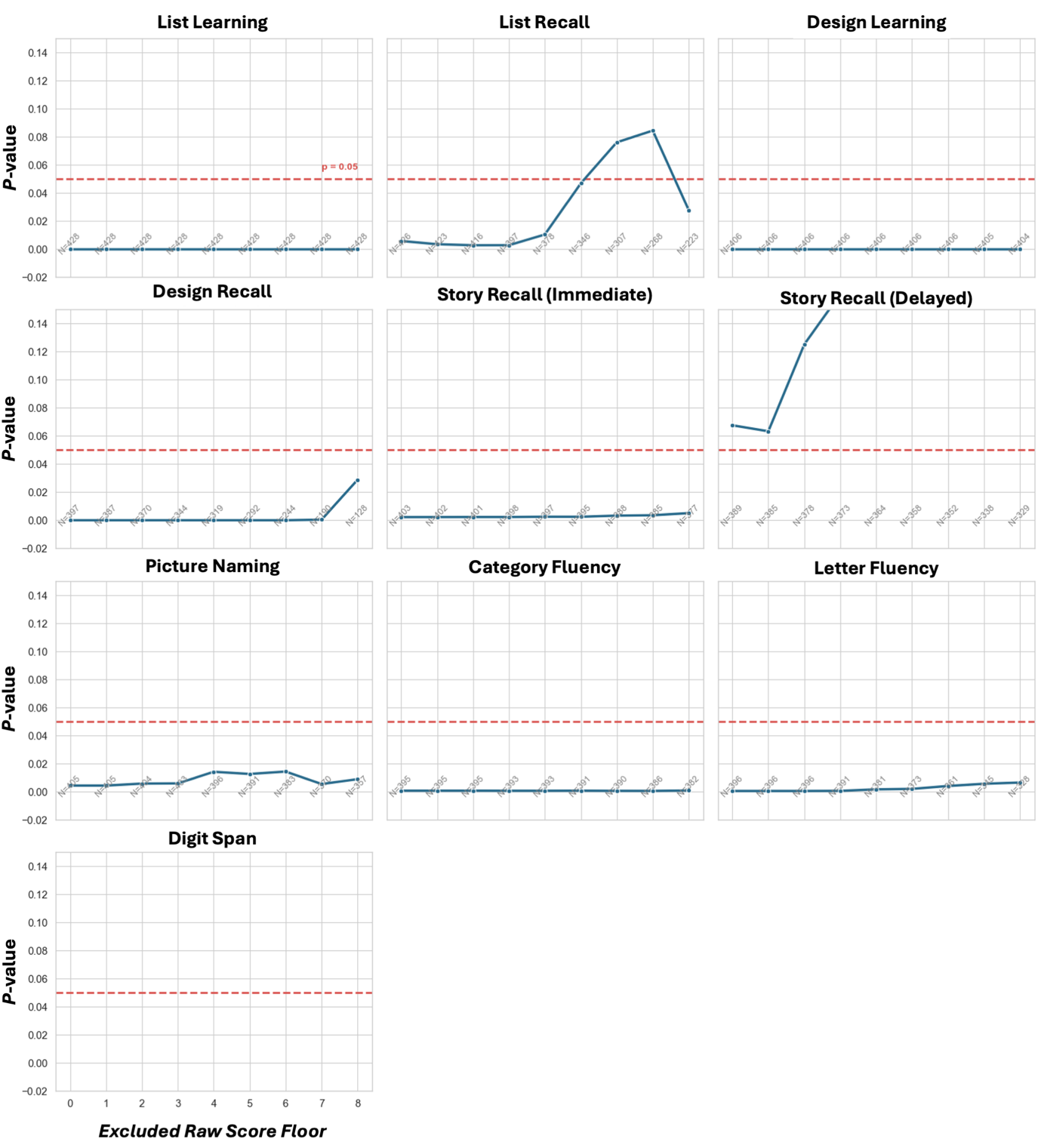

**Supplementary Figure 2. Sensitivity analysis of SuStaIn disease stage predicting postoperative cognitive decline.** The multiples line plot displays the trajectory of the $p$-value for the SuStaIn stage variable across incrementally stricter raw score exclusion thresholds. Patients scoring at or below the indicated raw score threshold on the x-axis were excluded to account for limited psychometric capacity to decline. The horizontal dashed red line indicates the threshold for statistical significance ($p=0.05$). Annotated values at the bottom of each panel indicate the remaining sample size ($N$) of the cohort at each respective exclusion threshold.

Subtype-outcome associations demonstrated strong domain-specific stability across the majority of tests (Supplementary Figure 3). However Domain-specific capacity limits were observed primarily on single-trial recall tasks. On ListA6, the Verbal Memory Subtype remained a significant predictor of 12-month decline from baseline through an exclusion threshold of ≤3 (N=397, *p* = 0.046), losing statistical significance at thresholds of 4 and above as sample size truncated (N≤378). On DesignA6, the Visual Memory Subtype was a significant predictor of postoperative change through threshold 3 (N=344, *p* = 0.026), but exhibited a sharp attenuation of significance at threshold 4 (N=319, *p* = 0.264). This shift reflects the fact that patients assigned to the Visual Memory Subtype exhibit marked baseline visual recall impairment, leaving minimal measurable range to demonstrate further post-surgical drop once lower baseline scores are removed.

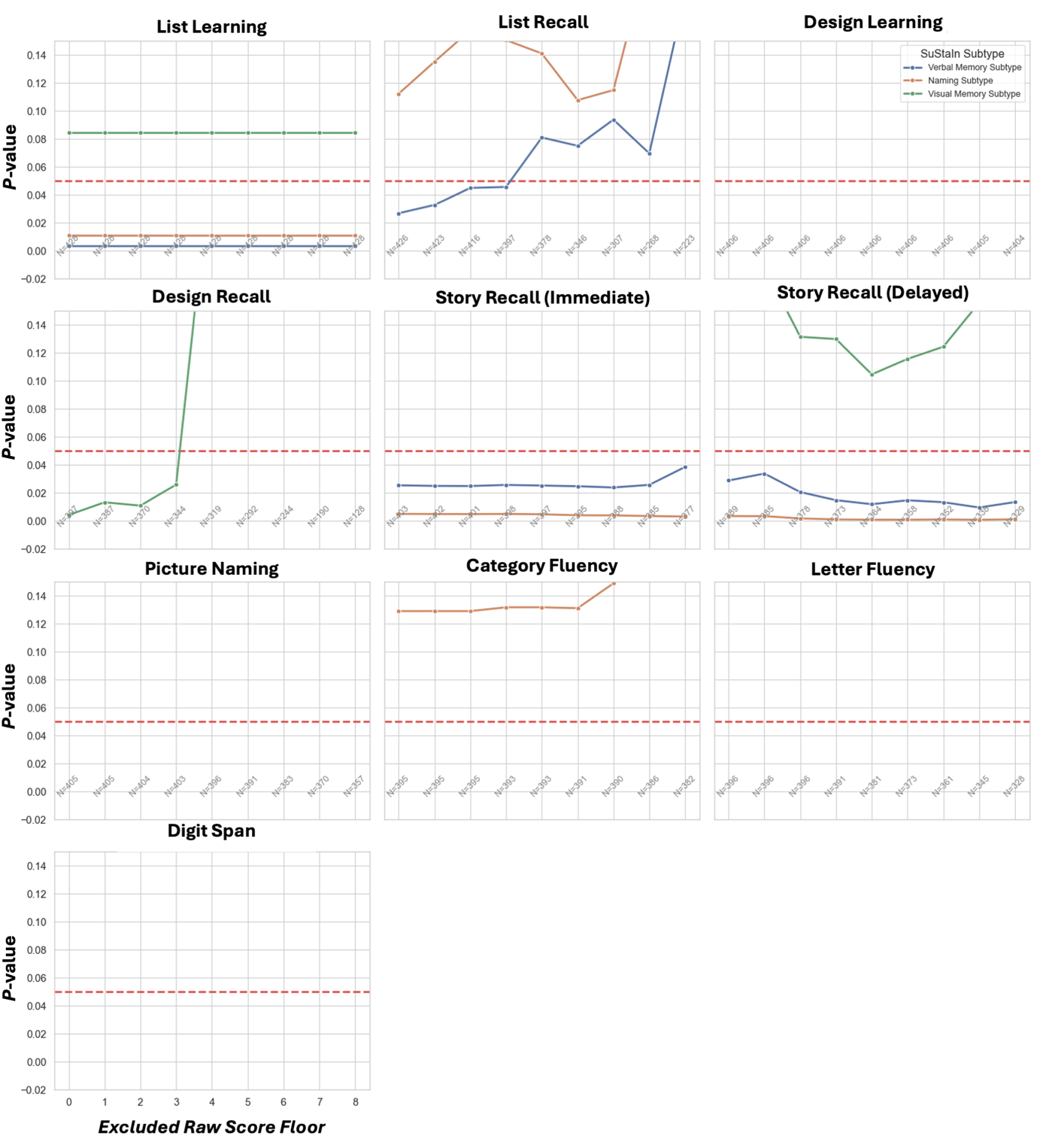

**Supplementary Figure 3. Sensitivity analysis of SuStaIn disease subtypes predicting postoperative cognitive outcome across varying preoperative score floor thresholds.** The multiples line plot displays the p-value trajectories for the three SuStaIn subtypes, Verbal Memory (blue line), Naming (orange line), and Visual Memory (green line), across nine neuropsychological parameters. Patients scoring at or below the raw score floor threshold indicated on the x-axis were iteratively excluded pre-regression to control for limited psychometric capacity to decline. The horizontal red dashed line marks the threshold for statistical significance (p=0.05). Sample size annotations at the base of each panel indicate the remaining cohort size (N) at each exclusion threshold.

### 5. Neuroimaging Meta-ROI

We employed meta-regions of interest (meta-ROIs) to reduce the number of multiple comparisons. These target regions were derived from the Desikan-Killiany atlas using FreeSurfer (v8.0). The composition of these meta-ROI can be seen in Supplementary Table 1.

*Supplementary Table 1. Constituent cortical regions comprising the meta-ROIs, defined according to the Desikan-Killiany atlas.*

| Meta-ROI | Regions Included |
| --- | --- |
| Left Frontal | Left caudal middle frontal  Left frontal pole  Left pars opercularis  Left pars orbitalis  Left pars triangularis  Left precentral  Left rostral middle frontal  Left superior frontal  Left lateral orbitofrontal  Left medial orbitofrontal |
| Right Frontal | Right caudal middle frontal  Right frontal pole  Right pars opercularis  Right pars orbitalis  Right pars triangularis  Right precentral  Right rostral middle frontal  Right superior frontal  Right lateral orbitofrontal  Right medial orbitofrontal |
| Left Parietal | Left inferior parietal  Left paracentral  Left postcentral  Left precuneus  Left superior parietal  Left supramarginal |
| Right Parietal | Right inferior parietal  Right postcentral  Right precuneus  Right superior parietal  Right supramarginal  Right paracentral |
| Left Occipital | Left cuneus  Left lateral occipital  Left lingual  Left pericalcarine |
| Right Occipital | Right cuneus  Right lateral occipital  Right lingual  Right pericalcarine |
| Left Medial Temporal Lobe | Left fusiform  Left parahippocampal  Left entorhinal |
| Right Medial Temporal Lobe | Right fusiform  Right parahippocampal  Right entorhinal |
| Left Extra-Temporal Lobe | Left bankssts  Left inferior temporal  Left middle temporal  Left superior temporal  Left transverse temporal  Left temporal pole |
| Right Extra-Temporal Lobe | Right bankssts  Right inferior temporal  Right middle temporal  Right superior temporal  Right transverse temporal  Right temporal pole |
| Left Hippocampus/Amygdala | Left-Hippocampus  Left-Amygdala |
| Right Hippocampus/Amygdala | Right-Hippocampus  Right-Amygdala |
| Left Subcortical | Left-Thalamus  Left-Caudate  Left-Putamen  Left-Pallidum |
| Right Subcortical | Right-Thalamus  Right-Caudate  Right-Putamen  Right-Pallidum |

### 6. Other Abnormality Thresholds

To ensure our findings were not dependent on the specific abnormality threshold used, we evaluated alternative thresholds of 0.5, 1.5, and 2.5. The same cognitive impairment subtypes consistently emerged across these variations, demonstrating the robustness of our classification (Supplementary Figure 4).

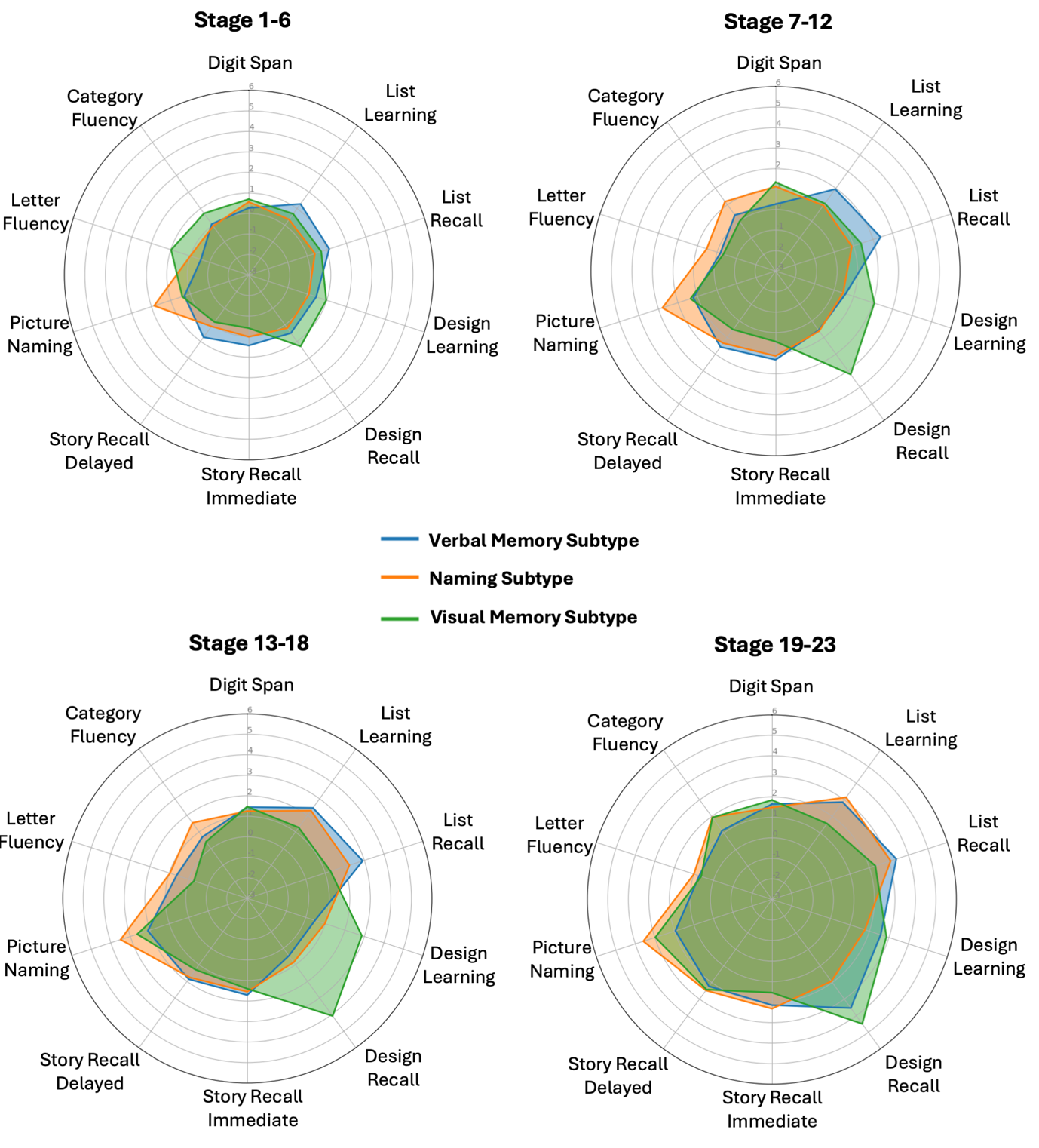

*Supplementary Figure 4.* ***Evolution of neuropsychological profiles across subtype stage and inference (SuStaIn) inferred stages and subtype at alterative abnormality thresholds.*** *Radar plots illustrate the mean cognitive performance of three distinct patient subtypes: Verbal Memory Subtype (blue), Naming Subtype (orange), and Visual Memory Subtype (green), across grouped stages of SuStaIn inferred disease progression. Radial axes denote mean standardised scores across ten neuropsychological assessments with greater percipheral points representing greater deficits. This covers the domains of working memory (Digit Span), verbal memory (ListA15, ListA6), verbal memory and attention (Story Immediate, Story Delayed), visuospatial memory (DesignA15, DesignA6), and language/fluency (GNT, FluS, FluAN). Solid lines represent the group mean, with shaded regions denoting the encompassing profile area. The cross-sectional trajectories demonstrate that while subtype-specific cognitive patterns are initially overlapping in early disease stages (Stage 1-6), they progressively uncouple. By advanced stages (Stage 19-23), the cohorts exhibit highly distinct, domain-specific multidimensional impairment signatures.*

### 7. Model Selection

To determine the optimal number of latent cognitive impairment subtypes, we trained and evaluated models ranging from one to five subtypes. Model selection was guided by two primary metrics: the Cross-Validation Information Criterion (CVIC) to evaluate overall model fit balanced against complexity, and out-of-sample log-likelihood across cross-validation folds to assess generalisability.

The CVIC values dropped sharply from the 1-subtype model to the 3-subtype model, after which the curve exhibited a clear "elbow" and plateaued through to the 5-subtype model (Supplementary Figure 5). Consistently, out-of-sample test set log-likelihood performance across the cross-validation folds improved (became less negative) up to the 3-subtype configuration (Supplementary Figure 6). Beyond three subtypes, additional clusters yielded marginal gains in log-likelihood and introduced higher variance across folds, suggesting diminishing returns and potential overfitting. Based on these convergent metrics of parsimony, model fit, and reproducibility, the 3-subtype model was selected as the optimal architecture for all subsequent analyses.

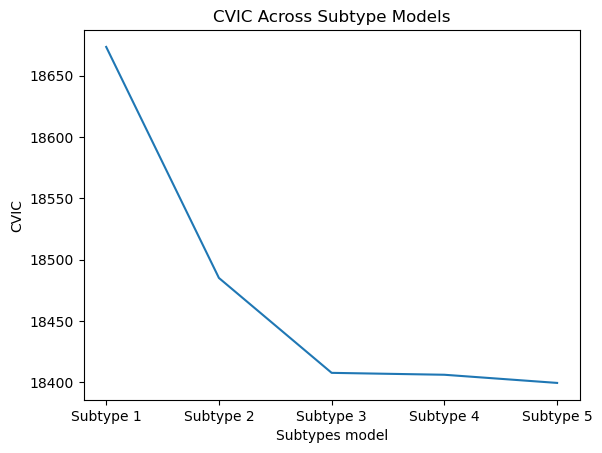

**Supplementary Fig. 5. Model fit evaluation across latent subtype configurations using CVIC.** The line plot displays the Cross-Validation Information Criterion (CVIC) evaluated for models ranging from one to five subtypes. A distinct elbow is observed at the 3-subtype model, indicating the point of optimal trade-off between model complexity and statistical fit.

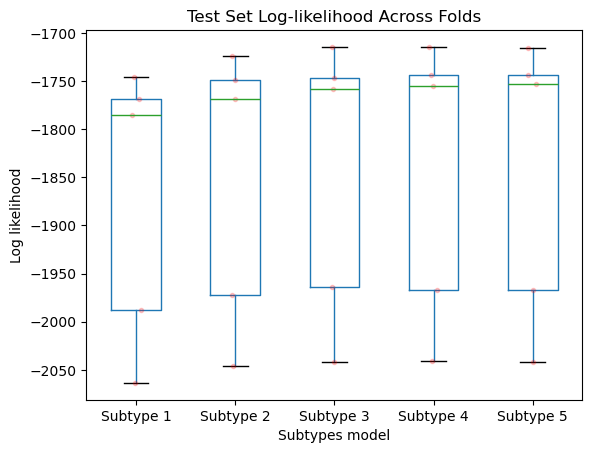

**Supplementary Fig. 6. Out-of-sample test set log-likelihood across cross-validation folds.** Boxplots illustrate the distribution of log-likelihood values calculated on the independent test sets across cross-validation folds for each subtype model configuration (1–5 subtypes). Individual data points (red dots) represent individual cross-validation folds. The green horizontal lines denote the median log-likelihood, which plateaus at the 3-subtype model, demonstrating stable generalisability.
